# Modeling the Cost-Effectiveness of the Next-Generation COVID-19 mRNA-1283 vaccine in the United States

**DOI:** 10.64898/2025.12.01.25341380

**Authors:** Kelly Fust, Michele Kohli, Keya Joshi, Shannon Cartier, Amy Lee, Nicolas Van de Velde, Milton Weinstein, Ekkehard Beck

## Abstract

**Aims:** COVID-19 disease burden in United States (US) adults ≥65 years and persons with underlying medical conditions remains high. This modeling study estimates the cost-effectiveness of the next-generation COVID-19 mRNA-1283 vaccine in those ages 12-64 at high-risk of severe COVID-19 outcomes and all adults ≥65 years.

**Methods:** mRNA-1283 was compared to no annual vaccination and originally licensed mRNA vaccines mRNA-1273 and BNT162b2. Analyses were conducted using a static decision-analytic model (1-year horizon). Vaccine effectiveness (VE) against infection and hospitalization for mRNA-1283 versus no vaccination was based on relative VE (rVE) from the Phase 3 pivotal randomized controlled trial comparing mRNA-1283 against mRNA-1273 and mRNA-1273 real-world data. rVE estimates for mRNA-1283 versus BNT162b2 were based on an indirect treatment comparison. The societal incremental cost per quality-adjusted life-year (QALY) gained and the benefit cost ratio (BCR) were calculated.

**Results:** During the 2025/2026 season, a single dose of mRNA-1283 was estimated to yield an incremental cost per QALY gained of $16,241 compared to no vaccine. The BCR for the base case strategy ranged from 2.16-9.74 USD returned for one dollar spent for mRNA-1283. mRNA-1283 was shown to dominate originally licensed COVID-19 vaccines in analyses of the target population. Results are sensitive to COVID-19 incidence, hospitalization rates, post-discharge mortality rates, and VE.

**Limitations:** The real-world effectiveness and safety of mRNA-1283 have not yet been established and relative VE estimates should be validated with real-world data. 2025/2026 COVID-19 incidence and vaccine uptake in the US is uncertain.

**Conclusions:** Study results suggest mRNA-1283 represents a highly cost-effective strategy (considering a $100,000-150,000 per QALY willingness-to-pay threshold) to reduce burden of COVID-19 among the target population. Given the finding of mRNA-1283 dominance in this population compared to originally approved mRNA vaccines, mRNA-1283 provides a valuable option to optimize US COVID-19 immunization programs and protect those most vulnerable.

## Introduction

Although the pandemic has ended, COVID-19 continues to place a significant burden on the United States (US) healthcare system. The Centers for Disease Control and Prevention (CDC) estimates that there were between 13.8 to 20.3 million COVID-19 illnesses, 380,000–540,000 COVID-19 hospitalizations and 44,000–63,000 COVID-19 deaths, between September 2024 and September 2025^1^. Increasing age continues to be a risk factor for severe COVID-19 disease.

During this timeframe, COVID-19 hospitalizations were primarily in older adults, with cumulative hospitalization rates (per 100,000) of 400.3 in those aged 65 years and over, compared with 78.1 in those aged 50–64, and 28.6 in those aged 18–49 years. In those under 65 years of age, patients with underlying medical conditions are at increased risk of severe COVID-19. Based on US CDC data presented at the Advisory Committee on Immunization Practices (ACIP) meetings in April and September 2025, 18 to 49 year old patients with diabetes or coronary artery disease were observed to have a 9 and 8 times higher hospitalization rate, respectively, than the overall population of community dwelling adults of same age in the US.^2^ Further, the risk of severe COVID-19 has been shown to increase with the number of underlying conditions, with data from ACIP showing that the adjusted relative risk (aRR) of hospitalization from COVID-19 for patients with ≥3 comorbid conditions was 5.9, compared to aRR=2.1 for patients with 1 underlying condition.^3^ Despite these well-documented risks, uptake of the updated 2024/25 COVID-19 vaccine only reached 44.4% in adults over 65 years of age by April 5, 2025^4^, which was lower than the 2024/25 influenza vaccine uptake in the same age group.^5^ mRNA-1283 (mNEXSPIKE, Moderna) is a FDA licensed^6^ and US ACIP recommended next generation COVID-19 vaccine^7^ with an innovative design, developed to focus the immune responses to increase protection against COVID-19 and to increase product stability compared with the originally licensed mRNA COVID-19 vaccines such as mRNA-1273^8^ (Spikevax, Moderna) or BNT162b2 (Comirnaty, Pfizer-BioNTech). In comparison to mRNA-1273 and BNT162b2, which encode the full-length SARS-CoV-2 spike protein, the next-generation COVID-19 vaccine mRNA-1283 uses an innovative design which includes only the receptor binding domain (RBD) and N-terminal domain (NTD) of the SARS-CoV-2 spike protein. Both of these domains contain immunodominant epitopes for neutralizing antibodies against SARS-CoV-2 and allowing enhanced immune response at a lower mRNA dose of 10 µg (e.g., 1/5th of dose of mRNA-1273)^8^. Additionally, shorter mRNA sequences are more stable^9^ and therefore improve refrigeration stability^10^. With its innovative design, mRNA-1283 also allows for inclusion in respiratory combination vaccines.

The pivotal NextCOVE randomized observer-blind active controlled phase 3 trial demonstrated the efficacy of mRNA-1283 compared to mRNA-1273^8^. Individuals aged 12 years and over were randomized to receive a bivalent original and Omicron-containing BA.4/5 formulation of mRNA-1283 or mRNA-1273 between March 23 and August 23, 2023. Higher immunogenicity of mRNA-1283 over mRNA-1273 was observed with a geometric mean titer ratio (GMR) against BA.4/5 of 1.3 (95% CI 1.2-1.5) among the overall population, which was even higher in among the vulnerable group of 65 years and older (GMR of 1.8; (95% CI 1.4-2.2). The relative vaccine efficacy (rVE) of mRNA-1283 versus mRNA-1273 was 9.3% (99.4 % CI- −6.6–22.8%) for COVID-19 symptomatic infection in the overall population, with higher rVE against infection observed in those aged 65 years and over (13.5%, 95% CI −7.7–30.6%)^8^ and in those with underlying medical conditions putting them at higher risk of severe COVID-19 (≥12 years 16.3%, 95% CI 1.8-28.7%; ≥65 years 28.1%, 95% CI 4.4-45.9%)^8^. In a post-hoc analysis, the rVE of mRNA-1283 vs mRNA-1273 against FDA defined severe COVID-19 was estimated at 38.1% (95% CI −6.7–64.1%)^8^.

A substantial body of real-world evidence suggests that mRNA-1273 is more protective than BNT162b2. This evidence includes head-to-head real-world effectiveness studies and GRADE meta-analyses of pairwise studies which capture different vaccine versions up until bivalent vaccines containing the original strain and Omicron BA.4/5. For example, in a large real-world study comparing the effectiveness of mRNA-1273 and BNT162b2 bivalent original/BA.4/5 COVID-19 vaccines in adults during the 2022/23 season, the relative VE (rVE) against outpatient visits and hospitalization of mRNA-1273 was higher by 5.1% (95% CI 3.2–6.9%) and 9.8% (95% CI 2.6–16.4%), respectively, compared with BNT162b2^11^. This enhanced protective effect was also seen in older adults and adults with underlying medical conditions. A systematic literature review (SLR) and meta-analysis (MA) reported a lower risk of symptomatic infections (risk ratio [RR] 0.74, 95% CI 0.56–0.97) and hospitalizations (RR 0.69, 95% CI 0.53–0.89) with mRNA-1273 compared with BNT162b2 in those 65 years and over^12^. Another SLR and MA focused on adults with medical conditions also found mRNA-1273 was more protective against infections (RR 0.85, 95% CI 0.79–0.92) and hospitalizations (RR 0.88, 95% CI 0.82–0.94) compared with BNT162b2, in adults with at least one underlying medical condition^13^. Based on these real-world effectiveness (RWE) data, mRNA-1273 likely delivers greater protection against infection and hospitalizations than BNT162b2. Phase 3 RCT data suggest that mRNA-1283 could provide greater protection compared with mRNA-1273 and therefore it is expected that mRNA-1283 could provide greater protection than BNT162b2. An indirect treatment comparison (ITC) using data from the NextCOVE trial^8^ and a large real-world study comparing mRNA-1273 vs BNT162b2 in adults aged 18 and over suggested that mRNA-1283 will be more effective than BNT162b2.^11,14^ The rVE against symptomatic COVID-19 infection of mRNA-1283 compared with BNT162b2 was 15.3% (95% CI 4.7-24.8%) among all adults and even more pronounced in those at highest unmet need such as adults 65 years and older with an rVE of 22.8% (95% CI 4.7–24.8%)^11^.

In May 2025, the Food and Drug Administration (FDA) authorized the use of mRNA-1283 for those 65 years of age and older, and 12 years through 64 years of age with at least one underlying condition that puts them at high risk for severe outcomes from COVID-19^13^. In September 2025, ACIP recommended^15^ that vaccination for COVID-19 be determined by shared clinical decision-making (SCDM) for those ages 6 months – 64 years and ≥65 years; it was recommended that all persons 12 years and older receive a COVID-19 vaccine, including mRNA-1283 based on SCDM. As the COVID-19 disease burden in the US remains high, especially for those at high risk for severe COVID-19 disease, decision makers are faced with the challenges both to increase vaccination coverage rates and to optimize COVID-19 vaccination policy with the available vaccine options. This modeling study was conducted to provide estimates of the public health impact and cost-effectiveness of the next-generation mRNA-1283 vaccine during season 2025/2026. The analysis compares mRNA-1283 to no vaccination and to originally licensed mRNA vaccines (mRNA-1273 and BNT162b2) within the licensed target population (ages 12-64 at high-risk of severe outcomes from COVID-19, and all adults aged ≥65 years) for mRNA-1283 in the US.

## Methods

### Overview

A Markov model with monthly cycles, described in detail by Fust et al. (2025)^16^, was used to project the potential number of symptomatic COVID-19 infections and clinical outcomes (hospitalizations, deaths and cases of long COVID) and associated costs from the societal perspective and quality-adjusted life-year (QALY) losses expected in the target population in the US. Analyses were performed over a one-year time horizon from September 2025 to August 2026 with a single dose of mRNA-1283 (annual vaccination) compared to no vaccination,^3,17,18^ consistent with the CDC cost-effectiveness analyses and Prosser et al. (2025). Cost-effectiveness, in terms of incremental cost per QALY gained, was assessed from the societal cost perspective. A scenario analysis was conducted in the target population where those 65 years and older were eligible to receive a second dose a minimum of two months after the first dose (semi-annual vaccination) considering current ACIP recommendations^19^ for 2 or more doses in 65 years and older. In addition, vaccination with mRNA-1283 was compared to vaccination with mRNA-1273 and BNT162b2.^20^ As the value of COVID-19 vaccination in post-pandemic settings is overlooked, especially from stakeholders outside the healthcare sector, the benefit cost ratio (BCR) was calculated for all scenarios to inform debate about the value of COVID-19 prevention holistically. This was defined as the benefit of vaccination (cost savings associated with prevention of disease and the monetized health gains due to averted COVID-19 outcomes) divided by the costs of vaccination (costs and monetized health lost due to vaccine adverse events (AEs)). Health was monetized using various assumptions: a QALY gained was valued at the standard US willingness to pay for QALYs gained of $100,000 and $150,000;^21^ value per statistical life-year (VSLY) $604,000^22^ and value per QALY (VQALY) $717,000.^22^ A completed CHEERS checklist can be found in the Technical Appendix.

### Model Structure

The cohort begins the model in the Well health state (Figure 1) in which each person is susceptible to symptomatic COVID-19 infection irrespective of their previous history of COVID-19 infections and vaccinations. In each month moving forward, a portion of those in the Well state may receive a vaccine and are then subject to vaccine-related AEs (AE rates, costs and QALY losses are described in the Technical Appendix Section 8). Each month, individuals in the Well health state are at risk of symptomatic COVID-19 infection. If they develop an infection, they move through the COVID-19 consequences decision tree (Figure 2) which estimates the subsequent clinical outcomes of infection and related costs and QALY losses. Those who die following their COVID-19 infection move into the Dead health state while all others return to the Well health state. Those who did not develop an infection remain in the Well health state.

**Figure 1.**
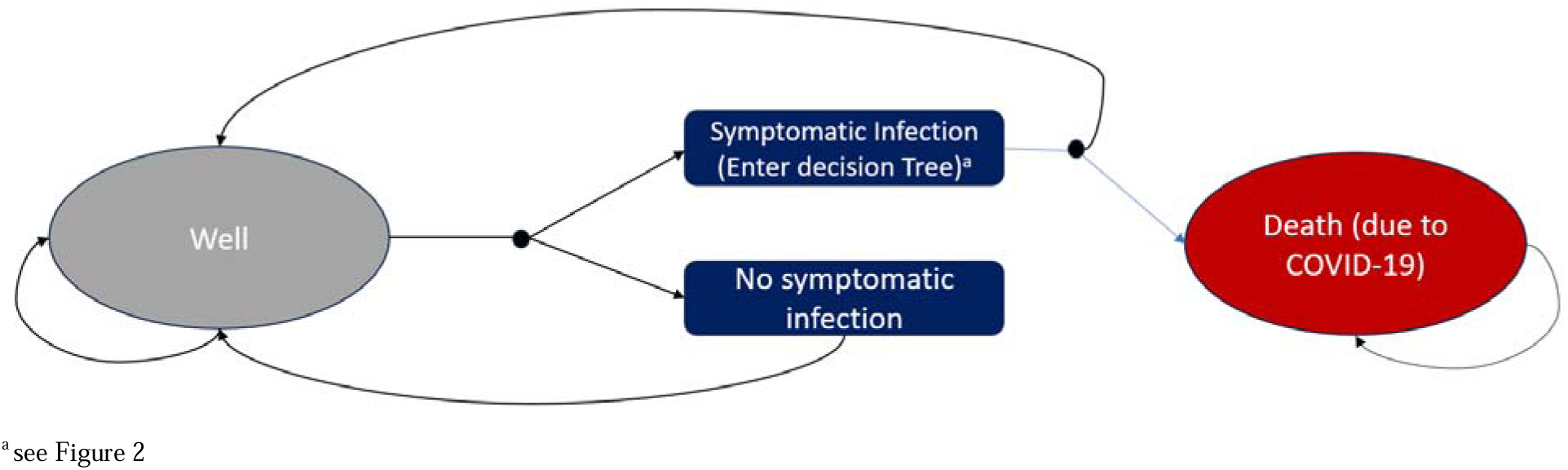
Illustration of the Markov model

**Figure 2:**
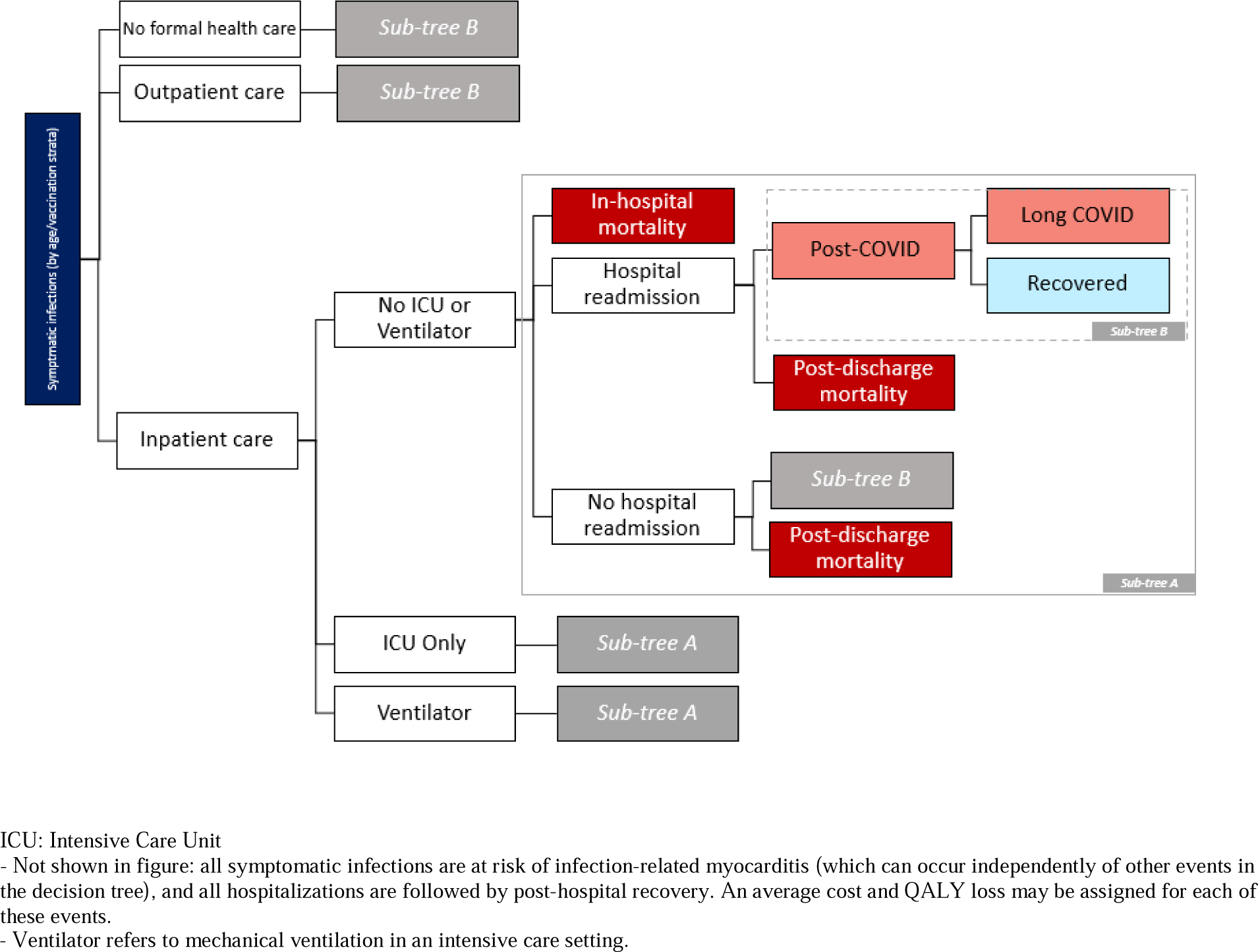
Illustration of the consequences of COVID-19 decision tree structure

The risk of infection and the risk of hospitalization once an infection has developed is reduced in those who are vaccinated. Starting from an initial VE, VE declines linearly on a monthly basis within the model. As a portion of the cohort can receive a new vaccine each month, the effectiveness calculation is a function of the fraction of the age group vaccinated each month and VE. The hospitalization VE values are adjusted so that the incremental protection against hospitalization above the protection against infection is applied to the probability of hospitalization. (See Technical Appendix Section 2).

### Target Population

The size of the cohort was based on the US population^23^ and FDA licensure for mRNA-1283.^6^ For ages 12-64, the cohort size eligible for vaccination is limited to high-risk individuals, estimated based on data from Kompanieyts et al. (2021)^24^ for adolescents and Panagiotakopoulos (2025)^25^ for adults 18-64 (See Appendix). A total of 214,034,195 people, including persons 65 and older in addition to eligible 12-64 year olds were modelled.

### Infection Incidence

The model required the incidence of symptomatic COVID-19 infection in the target population when no one receives the vaccine. As these data are not collected by the CDC, this input was approximated based on estimates of observed hospitalization rates per 100,000 by age group for the 2023-2024 and 2024-2025 seasons from COVID NET^26^, using data on vaccine coverage and effectiveness and the conditional probability of hospitalization given infection in a modified version of the model, as described in the Technical Appendix. Given the uncertainty in COVID-19 incidence in the post-pandemic setting, similar to approaches for seasonal influenza,^27^ an average of the estimated incidence rates for September 2023 to August 2024 and September 2024 to August 2025 were used to project the COVID-19 incidence for season 2025/2026 for the base case in this analysis. Use of the values from the individual seasons was tested in sensitivity analyses. For high-risk patients, the incidence of symptomatic COVID-19 infection was assumed to be equivalent to the incidence of COVID-19 infection for the general population, as it is assumed that high-risk patients are more likely to experience more severe COVID-19 outcomes but are not at increased risk of developing COVID-19 infection.^3^

### Vaccine Coverage

The vaccine coverage rate (VCR) of the first seasonal dose of the vaccines (annual vaccination) for season 2025/2026 was assumed based on data from COVIDVaxView.^28,29^ Data from September 2024 to July 2025 were available, with no further uptake assumed in August 2025 (See figure in the Technical Appendix Section 4). Data for those 12-17 years was taken from the pediatric database, while data for the other age groups from the adult database. For the scenario where those aged 65 years and over received a second seasonal dose (semi-annual vaccination), in the absence of CDC-reported data on second dose coverage for the 2024/2025 season, the same coverage as observed during 2023/2024 season was used as an approximation of the 2024/2025 estimates. The use of the 2023/2024 data likely reflects a conservative estimate of second dose coverage, as data from Los Angeles county suggest second dose coverage rates of approximately 22% in those ≥65 years who already had received a first those.^30^ In absence of CDC data, the VCR for the high-risk population was conservatively assumed to be equivalent to the general population.

### Vaccine Effectiveness

The VE values used for the model analytic time horizon were derived based on the age-specific rVEs of mRNA-1283 compared to the mRNA-1273 vaccine from the pivotal phase 3 randomized clinical trial (RCT) NextCOVE^8,31^ and the 2024-2025 US real-world VEs of mRNA-1273.712 targeting KP.2.^32^ Estimates for high-risk populations in RCT and real-world effectiveness (RWE) studies were used for those aged 18-64 years with high-risk in the model. mRNA-1273 VEs were based on a large, US nationwide RWE study providing interim VE estimates of mRNA-1273. For those aged 12 to 17 years, the 2024-2025 VEs for mRNA-1273 against infection and hospitalization were assumed to be the same and were based on MacNeil’s estimate of the real-world VE of 2024-2025 COVID-19 vaccines against emergency department and urgent care visits in children aged 5-17 years, as these were the only available US VE estimates applicable to mRNA-1273 for this age range.^33^ rVE values for mRNA-1283 compared to mRNA-1273 were applied to the mRNA-1273 VEs to calculate the initial VEs for mRNA-1283 as displayed in Table 1 (details provided in the Technical Appendix). The 95% confidence intervals (CIs) for the VE of mRNA-1283 against infection and hospitalization were derived based on Monte-Carlo simulations considering the 95% CIs of the VE of mRNA-1273 and of the rVE estimates of mRNA-1283 vs mRNA-1273.

**Table 1.**
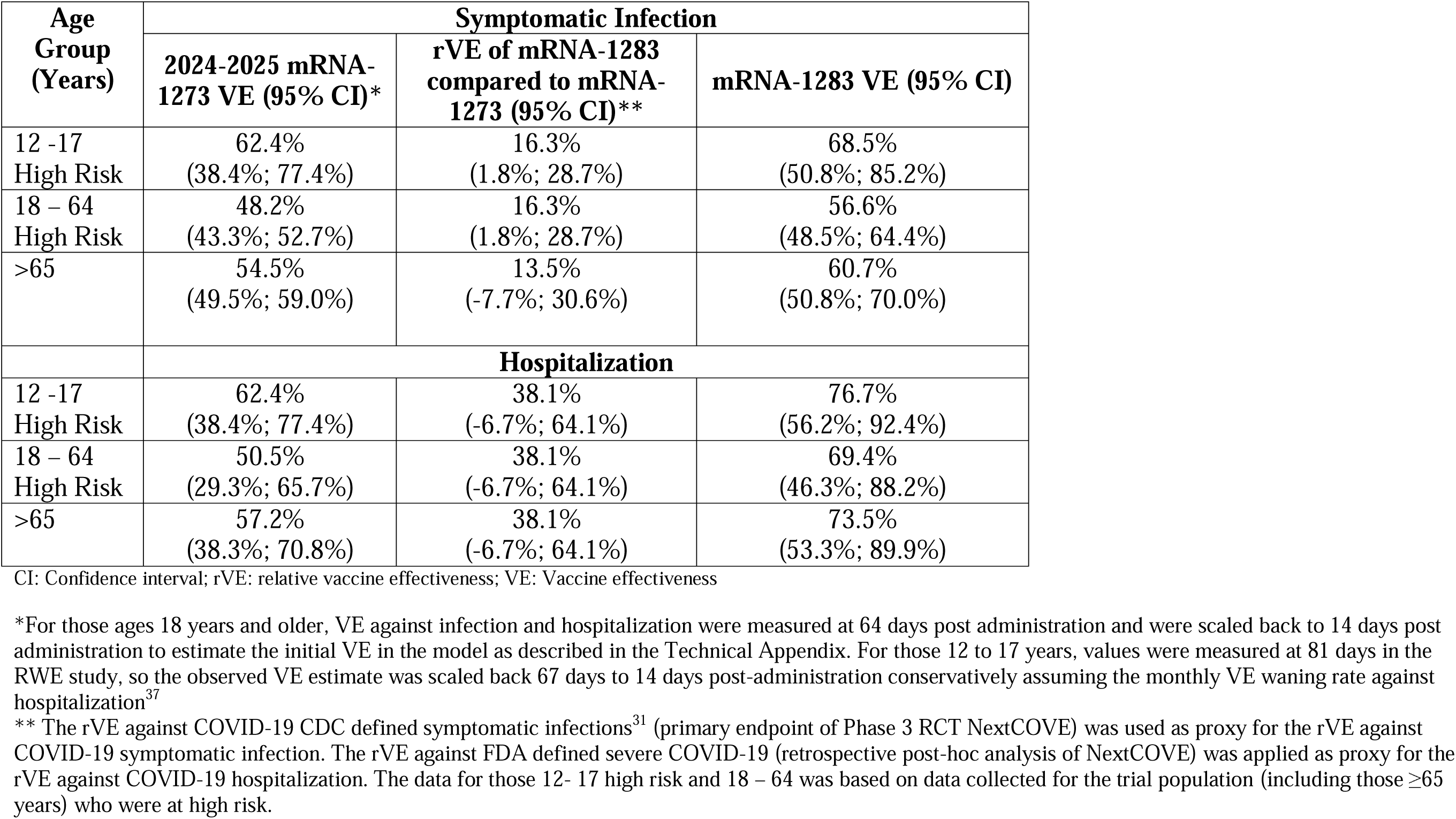
mRNA-1283 COVID-19 Initial Vaccine Effectiveness.

| Age Group (Years) | Symptomatic Infection |  |  |
| --- | --- | --- | --- |
|  | 2024-2025 mRNA-1273 VE (95% CI)* | rVE of mRNA-1283 compared to mRNA-1273 (95% CI)** | mRNA-1283 VE (95% CI) |
| 12 -17<br>High Risk | 62.4%<br>(38.4%; 77.4%) | 16.3%<br>(1.8%; 28.7%) | 68.5%<br>(50.8%; 85.2%) |
| 18 – 64<br>High Risk | 48.2%<br>(43.3%; 52.7%) | 16.3%<br>(1.8%; 28.7%) | 56.6%<br>(48.5%; 64.4%) |
| >65 | 54.5%<br>(49.5%; 59.0%) | 13.5%<br>(-7.7%; 30.6%) | 60.7%<br>(50.8%; 70.0%) |
| Hospitalization |  |  |  |
| 12 -17<br>High Risk | 62.4%<br>(38.4%; 77.4%) | 38.1%<br>(-6.7%; 64.1%) | 76.7%<br>(56.2%; 92.4%) |
| 18 – 64<br>High Risk | 50.5%<br>(29.3%; 65.7%) | 38.1%<br>(-6.7%; 64.1%) | 69.4%<br>(46.3%; 88.2%) |
| >65 | 57.2%<br>(38.3%; 70.8%) | 38.1%<br>(-6.7%; 64.1%) | 73.5%<br>(53.3%; 89.9%) |

CI: Confidence interval; rVE: relative vaccine effectiveness; VE: Vaccine effectiveness
\*For those ages 18 years and older, VE against infection and hospitalization were measured at 64 days post administration and were scaled back to 14 days post administration to estimate the initial VE in the model as described in the Technical Appendix. For those 12 to 17 years, values were measured at 81 days in the RWE study, so the observed VE estimate was scaled back 67 days to 14 days post-administration conservatively assuming the monthly VE waning rate against hospitalization<sup>37</sup>
\*\* The rVE against COVID-19 CDC defined symptomatic infections<sup>31</sup> (primary endpoint of Phase 3 RCT NextCOVE) was used as proxy for the rVE against COVID-19 symptomatic infection. The rVE against FDA defined severe COVID-19 (retrospective post-hoc analysis of NextCOVE) was applied as proxy for the rVE against COVID-19 hospitalization. The data for those 12- 17 high risk and 18 – 64 was based on data collected for the trial population (including those ≥65 years) who were at high risk.

For the scenarios, where mRNA-1283 was compared to mRNA-1273 or BNT-162b2, the initial VE estimates shown in Table 2 were used. To estimate the VE of BNT162b2, the rVE estimates derived in an indirect treatment comparison (ITC)^14^ were applied to the VE of mRNA-1283. For the 65 plus and the high-risk 18 to 64 years of age populations the rVE estimate of mRNA-1283 vs BNT162b2 against symptomatic COVID-19 infection of 22.8% (95% CI: 3.7%; 38.1%) and 19.0% (95% CI: 4.9; 31.0%) were assumed. For the rVE against hospitalization, the overall population estimate of 44.1% (95% CI: 3.2; 67.7%) was assumed; this number was derived applying the same methods as outlined in Beck et al. but using the NextCOVE estimate of the rVE of mRNA-1283 vs mRNA-1273 (38.1%, 95% CI: −6.7; 64.1%) against FDA defined severe COVID and the Kopel et al.^34^ estimate of the rVE of mRNA-1273 vs BNT162b2 (9.8%, 95% CI: 2.6; 16.4%) against COVID-19 hospitalization.^35^ For ages 12 to 17, BNT162b2 VEs against infection and hospitalization were assumed to be the same as mRNA-1273 as the values were based on a mixed population that received either of the two vaccines.

**Table 2.** Initial Vaccine Effectiveness for the Comparison of mRNA-1283 to mRNA-1273 and BNT-162b2.

| Age Group (Years) | Comparison of mRNA-1273 & mRNA-1283 |  |  | Comparison of mRNA-1283 & BNT-162b2 |  |  |
| --- | --- | --- | --- | --- | --- | --- |
|  | Symptomatic Infection |  |  |  |  |  |
|  | 2024-2025 mRNA-1273 VE | rVE of mRNA-1283 compared to mRNA-1273 (95% CI) | mRNA-1283 VE (Range) | mRNA-1283 VE | rVE of mRNA-1283 compared to BNT-162b2 (95% CI) | BNT-162b2VE (Range) |
| 12 -17 High Risk | 62.4% | 16.3% (1.8%; 28.7%) | 68.5% (63.1%; 73.2%) | 68.5% | n/a | 62.4% |
| 18 – 64 High Risk | 48.2% | 16.3% (1.8%; 28.7%) | 56.6% (49.1; 63.0%) | 56.6% | 19.0% (4.9%; 31.0%) | 46.5% (37.2%; 54.4%) |
| >65 | 54.5% | 13.5% (0%; 30.6%)* | 60.7% (54.5%; 68.4%) | 60.7% | 22.8% (3.7%; 38.1%) | 49.0% (36.5; 59.2%) |
|  | Hospitalization<sup>†</sup> |  |  |  |  |  |
| 12 -17 High Risk | 62.4% | 38.1% (0%; 64.1%)** | 76.7% (62.4%; 86.5% ) | 76.7% | n/a | 62.4% |
| 18 – 64 High Risk | 50.5% | 38.1% (0%; 64.1%)** | 69.4% (50.5%; 82.2%) | 69.4% | 44.1% (3.2%; 67.7%) | 45.2% (5.1%; 68.3%) |
| >65 | 57.2% | 38.1% (0%; 64.1%)** | 73.5% (57.2%; 84.6%) | 73.5% | 44.1% (3.2%; 67.7%) | 52.6% (17.9%; 72.6%) |

\* Lower range truncated from -7.7% to 0% for this comparison
\*\* Lower range truncated from -6.7% to 0% for this comparison
<sup>†</sup> If Hospitalization is lower than infection VE, the incremental protection against the probability of hospitalization given infection is set to 0.
CI: Confidence interval; rVE: relative vaccine effectiveness; VE: Vaccine effectiveness

The monthly VE waning of mRNA vaccines against infection for Omicron variants relative to those who had never been vaccinated was estimated in a meta-analysis by Higdon et al., (2022)^36^ to be 4.75% (95% CI 3.05%; 6.75%). We considered this to be an appropriate source for VE waning against infection because there is minimal to no protection against COVID-19 infection a year after receiving vaccination, which implies that waning of annual vaccination VE would be similar to the waning of VE of those who received initial doses of COVID-19 vaccine. However, as protection against hospitalization is longer lasting, a study by Andersson et al., (2024)^37^ which examined the effectiveness of XBB.1.5 containing COVID-19 mRNA vaccines was used to estimate the base case rate of waning of VE against hospitalization of 2.46% per month, also considering these findings to be consistent with findings for JN.1 and KP.2 updated vaccines.^3,38^ In sensitivity analyses, the estimate from Higdon et al, against hospitalization (1.37%), as well as an alternate source which examine the durability of the mRNA-1273 XBB.1.5 vaccine (3.87%)^39^ were tested.

### Probabilities for the COVID-19 Consequences Tree

Key probability inputs are summarized in Table 3 and the text. Further details are provided in the Technical Appendix. Patients with symptomatic COVID-19 infections are divided by their highest level of medical care received: 1) No formal health care (not hospitalized nor was outpatient care received); 2) Outpatient care (not hospitalized but did receive outpatient physician visits or emergency department visits without hospital admission); and 3) Inpatient care. These age-specific probabilities were estimated based on database analyses and adjusted to represent the high-risk population for ages 12 to 64 years as described in the Technical Appendix (Supplemental Table 10). Hospitalized patients are further stratified by location of care, including general ward (no intensive care unit [ICU] or ventilation required), ICU (excluding extracorpeal membrane oxygenation or invasive ventilation), or ICU with mechanical ventilation based on data from the COVID-Net surveillance system.^26^ In absence of data, the proportions for the high-risk populations were assumed to be equivalent to the general population.

**Table 3.**
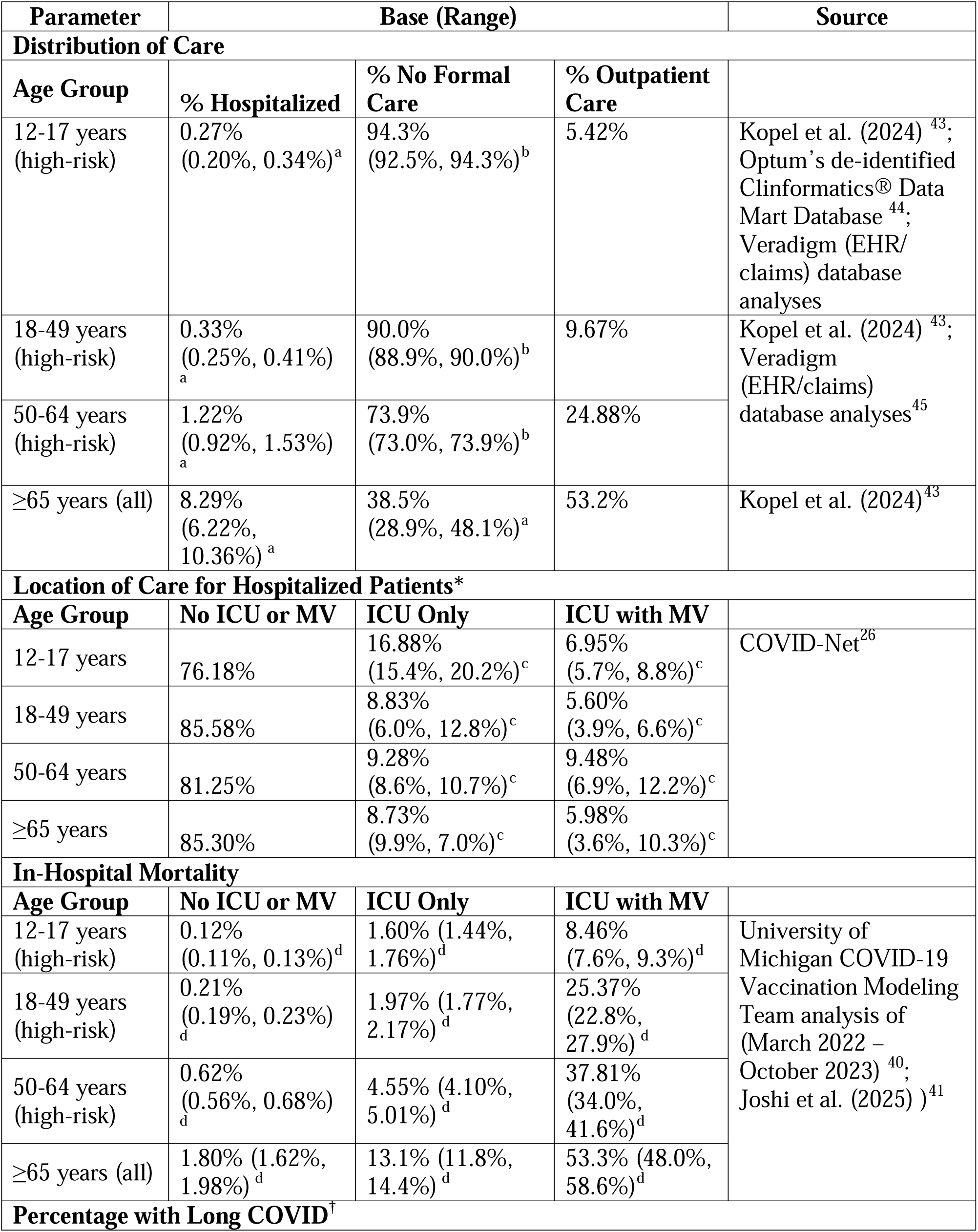
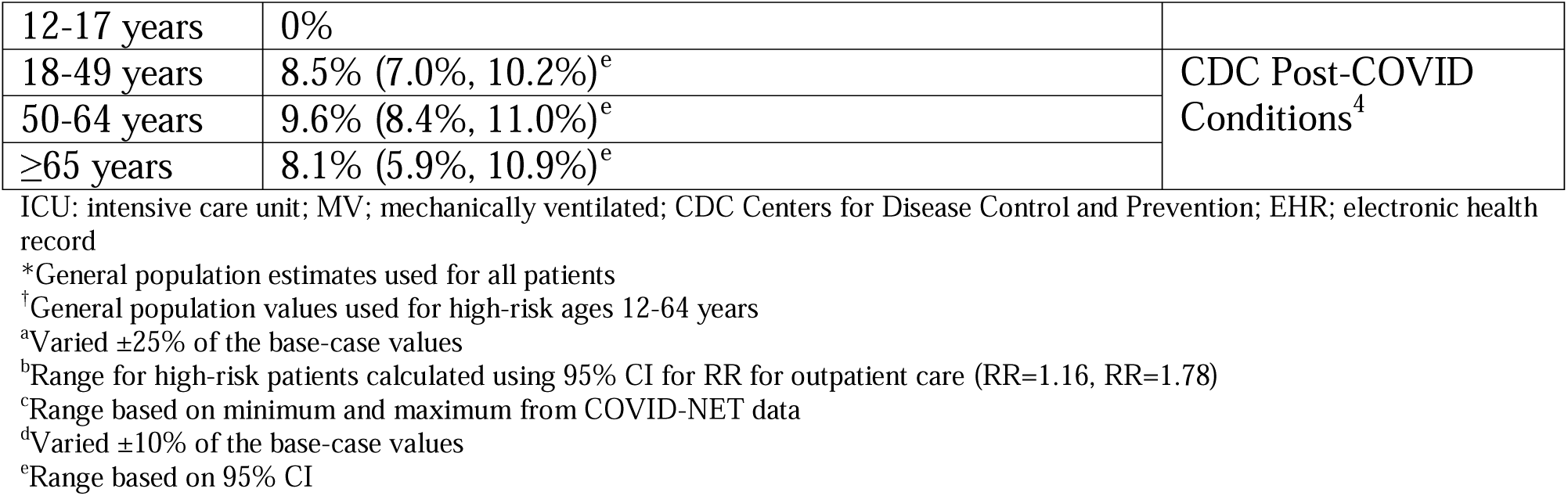
Key decision tree probabilities, base case and range for deterministic sensitivity analyses.

Hospitalized patients may die, be discharged and then readmitted, or simply discharged. Estimates of in-hospital mortality for the general population^40^, stratified by age and in-hospital location of care, were based on COVID-NET data from March 2022 – October 2023 as analyzed by the University of Michigan COVID-19 Vaccination Modeling Team.^40^ Based on Joshi et al. (2025) ^41^, it was conservatively assumed that the relative risk for in-hospital mortality for high-risk patients relative to non-high-risk patients was 1.23, considering an estimate derived for patients with diabetes. The COVID-NET surveillance system tracks only in-hospital deaths and not post-discharge mortality. The 30-day readmission and post-discharge mortality probabilities were estimated from a meta-analysis for the general population.^42^ The readmission rate for the high-risk populations was calculated using the RR for patients with diabetes (RR=1.27) from Joshi et al. (2025)^41^ and a weighted^25^ average of high-risk and general population of 9.35% (range: 4.1%, 9.28%) was used. A post-discharge mortality of 7.87% (95% CI: 2.8% to 13.0%) was assumed to be the same for both the high-risk and general populations and applied to all age groups.

The probability of long COVID and infection-related myocarditis is described in the Technical Appendix Sections 7 and 9.

### Quality of Life

QALY losses due to COVID-19 related outcomes and AEs were assumed to be the same for the general population and high-risk patients. They are summarized in Tables 25 and 26 of the Appendix.

QALY losses associated during the acute infection phase were estimated for consistency with data presented to ACIP by the University of Michigan COVID-19 Vaccination Modeling Team.^40^ Caregiver QALY losses were included for patients <18 years based on caregiver spillover data reported by Mercon et al. (2025) ^46^; caregiver QALY losses were not included for adult patients. Patients receiving no formal care were assumed to experience QALY losses equal to those seeking outpatient care, as the health states described in Mercon et al. (2025) ^46^ do not include requirements for medical attendance.

Post-COVID QALY losses were estimated based on data from Sandmann et al. (2022) ^47^ and the “Post-HOSPitalization COVID-19 study” and applied to patients who sought outpatient care and patients who were hospitalized, respectively; patients who did not seek formal health care were assumed to have no post-infection QALY loss (post-COVID QALY losses were only applied to those that survived a medically attended acute phase of illness). The post-COVID QALY losses applied to those who received outpatient care are assumed to reflect the 6-month period post-infection, while the estimates for hospitalized patients reflect the 1-year period post-discharge; these estimates exclude the acute phase of care. It is assumed that the post-COVID QALY losses capture long-term health impacts associated with not only long COVID, but also exacerbations of underlying medical conditions and any other long-term impacts of COVID-19 infection. No additional QALY losses specific to long COVID are applied.

The loss of QALYs due to infection-related mortality includes losses over the patient’s entire life span, not just the months lost within the one-year time horizon. QALYs lost were calculated as the discounted present values (at a rate of 3% per year)^48^ of the lost utility over the remaining expected lifetime, based on age-specific life expectancy at the time of the event ^49^ and standard baseline utility values.^50^

### Healthcare Costs

All cost estimates are presented in 2025 US dollars and are summarized in the Appendix with key inputs described below.

Hospitalization costs, stratified by location of care within the hospital setting, for the general population were estimated from Yehoshua et al. ^51^, a database analysis designed to examine healthcare costs and resource utilization for patients with a hospital admission with an ICD-10 code for COVID-19 (U07.1) between February 4, 2023 and February 29, 2024 (XBB and JN.1 Omicron lineages). The cost of inpatient care for high-risk patients was estimated using a similar approach to the probability of hospitalization given symptomatic infection and inpatient mortality. It was assumed that the RR for increased costs for high-risk patients was 1.07 based on Kapinos et al.^52^ Weighted averages of the high-risk and general population estimates of hospitalization costs were calculated as shown in the Technical Appendix Table 21 to be $51,415 for patients who received ventilator support, $22,449 for those treated in the ICU without ventilation and $51,415 for all other patients. It was assumed that patients experiencing hospital readmission would be readmitted to their original location of care, and readmission costs were assumed to be equivalent to the initial hospitalization costs.

A post-COVID cost toll, estimated separately for outpatients and hospitalized patients, is applied to those who survive the acute phase of COVID-19 infection. This toll is designed to capture medical costs related to long-term health effects of COVID-19 infection in the year following COVID-19 diagnosis, including, but not limited to, long COVID. Post-COVID costs for those ages ≥65 years were based on Chambers et al. (2023)^53^, and high-risk estimates were based on Scott et al. (2024) ^54,55^; the model uses a weighted average of the high-risk and general population in all analyses. To avoid double-counting with the post-COVID estimates, long COVID is associated with productivity losses in model analyses only, and no additional cost impact specific to long COVID is applied. 2025/2026 season private sector list prices were assumed for the unit cost of the vaccines: - $177.12 for mRNA-1283, $141.80 for mRNA-1273 and $147.69 for BNT162b2.^56^ An administration fee of $20.05 was included in base case.^57^ A scenario analysis examining a vaccine administration fee of $40 based on the Centers for Medicare & Medicaid Services (CMS) national payment allowance in 2024-2025 for COVID-19 vaccine administration.^58^

### Lost Productivity

Non-health care associated costs for COVID-19 patients included lost productivity from time missed from work due to receiving a COVID-19 vaccine and AEs, COVID-19 infection and long COVID, and infection-induced myocarditis. Caregiver productivity loss was also included to account for time loss from work to care for patients with COVID-19. It was assumed that all patients under 18 years of age required caregiver time for vaccination, AEs associated with vaccination, and during the acute infection period; it was further assumed that adults would not require a caregiver for vaccination, AEs, or during the acute symptomatic infection period. It was assumed that all patients with long COVID (both <18 and ≥18 years of age) would require a caregiver based on data from Kwon et al. (2024)^59^. Lost productivity estimates are provided in the Technical Appendix; general population estimates were used for all model age groups, with the exception of the percentage in the labor force, which was adjusted for the high-risk population.

In the base-case analysis, the productivity losses due to COVID-19 premature mortality were included, using a human capital approach based on age-stratified market productivity loss estimates from Grosse et al. (2019).^60^ The inclusion of productivity loss due to premature COVID-19 mortality is consistent with recent health economics analyses performed by ACIP and the CDC for RSV and pneumococcal vaccines.^61–63^ A scenario analysis where non-market productivity losses were also included was performed; additionally, a scenario analysis excluding all premature mortality-related productivity losses (both market and non-market) was performed. *Sensitivity Analyses*

Deterministic sensitivity analyses (DSAs) performed to assess the impact of key variables on estimated symptomatic infections, COVID-19 hospitalizations, deaths are presented in the technical appendix. Deterministic sensitivity analyses were also designed to assess the robustness of the cost-effectiveness results. Several of these have been described above in the text and include parameters related to incidence of COVID-19 infection, probabilities of infection consequences, VE and vaccine coverage. In addition, COVID-19 related costs and QALY losses were also varied. Where available, estimates were varied according to 95% confidence intervals (CIs) or reported ranges. For all other DSAs, an alternative data source was used or parameters were varied by ±10% or ±25% of their base-case value.

### Scenario Analyses

Several scenario analyses described below were conducted (further detail on model inputs are presented in the Technical appendix). Two scenario analyses were conducted for VE: 1) rVE against hospitalization for mRNA-1283 versus mRNA-1273 were set to be equal to the rVE against infections; and 2) VE against symptomatic COVID-19 infection for adults ages 18 years and older from ICATT study.^64^ The model cannot estimate any benefit associated with changes in transmission, which may include the reduction in secondary transmission when a vaccinated person develops an infection. This effect was demonstrated by the CDC RIGHT study^33^, which estimated that if infected people had been vaccinated within the prior 6 months, their risk of transmitting to household contacts was reduced compared to the who had not been vaccinated (adjusted RR 0.55; VE: 45%). These results were therefore used to create an indirect benefit scenario.

Vaccine coverage rates in September were increased either by 5% or 10%, for all ages and for the ≥65 years subgroup. The percentage of patients seeking no formal care for COVID-19 related illness was assumed to be equal to CDC influenza rates from the 2023-2024 season. Several scenario analyses were also performed specific to high-risk individuals, including assuming that 50% of 12-17 year-olds are considered high-risk, that the likelihood of receiving outpatient care is equivalent to the general population, that the RR of inpatient mortality is 1.62 based on cardiovascular disease^41^, and using the percentage hospitalized based on the RR derived from Wilson et al. (2025) (RR=1.48). Scenario analysis considering the healthcare perspective as well as the societal perspective including also non-market productivity losses associated with premature mortality, excluding all productivity losses (market and non-market) associated with premature mortality, and excluding caregiver productivity losses and by considering opportunity costs (Supplemental Section 12).

The number of antibiotic prescriptions that can be avoided through vaccination with mRNA-1283 was investigated (further details are provided in the Technical appendix section 18).

For the analyses comparing mRNA-1283 to mRNA-1273 and BNT-162b2, a range of rVE values were used to calculate the ranges used for sensitivity analyses as shown in Table 2; scenario analyses examining COVID-19 incidence and vaccine coverage were also performed for these comparisons.

## Results

### Base case analysis (compared to no vaccine)

As shown in Table 4, use of an annual dose of mRNA-1283 is estimated to reduce the number of symptomatic infections, hospitalizations, and deaths COVID in the target population by 2.2 million, 137,391, and 17,960 compared to no vaccine, respectively (additional clinical outcomes are presented in Table 4). The corresponding numbers needed to vaccinate (NNV) to avert one COVID-19 symptomatic infection, outpatient episode, hospitalization, death and long COVID were estimated at 27, 90, 432, 3,301, and 908 respectively.

**Table 4:** Health outcomes and health outcomes averted.

| Strategy | Health outcomes |  |  |  |  |
| --- | --- | --- | --- | --- | --- |
|  | Cases | Outpatient | Long COVID | Hosp. | Deaths |
| **Base-Case: Annual dosing for 12-64 High-Risk, ≥65 All** |  |  |  |  |  |
| **No vaccine** | 29,096,031 | 6,249,345 | 577,355 | 604,259 | 77,099 |
| **mRNA-1283** | 26,879,616 | 5,587,186 | 512,096 | 466,869 | 59,140 |
| **Outcomes Averted by mRNA-1283 (%)** | 2,216,414 (7.6%) | 662,158 (10.6%) | 65,259 (11.3%) | 137,391 (22.7%) | 17,960 (23.3%) |
| **Scenario Analysis: Annual dosing for 12-64 High-Risk, Semi-annual dosing for ≥65 All** |  |  |  |  |  |
| **No vaccine** | 29,096,031 | 6,249,345 | 577,355 | 604,259 | 77,099 |
| **mRNA-1283** | 26,841,678 | 5,565,488 | 510,231 | 465,237 | 58,923 |
| **Outcomes Averted by mRNA-1283 (%)** | 2,254,353 (7.7%) | 683,857 (10.9%) | 67,124 (11.6%) | 139,023 (23.0%) | 18,176 (23.6%) |

Hosp.: Hospitalizations

**Table 5:** Base-case cost-effectiveness results.

| Vaccination strategy | Total Costs | Total QALYs Lost | $\Delta$ Costs | $\Delta$ QALYs Gained* | ICER ($\Delta$ Cost per QALY Gained) |
| --- | --- | --- | --- | --- | --- |
| **Base-Case: Annual dosing for 12-64 High-Risk, $\geq 65$ All** |  |  |  |  |  |
| No vaccine | \$64,579,484,159 | 1,017,879 | -- | -- | Reference |
| mRNA-1283 | \$67,360,095,079 | 846,729 | \$2,780,610,919 | 171,150 | \$16,247 |
| **Scenario Analysis: Annual dosing for 12-64 High-Risk, Semi-annual dosing for $\geq 65$ All** |  |  |  |  |  |
| No vaccine | \$64,579,484,159 | 1,017,879 | -- | -- | Reference |
| mRNA-1283 | \$68,295,993,219 | 844,691 | \$3,716,509,060 | 173,188 | \$21,459 |

Abbreviations: $\Delta$ , difference; ICER, incremental cost-effectiveness ratio; QALY, quality-adjusted life-year
\*Difference in QALYs gained is equivalent to negative one times the difference in total QALYs lost.

The majority of severe COVID-19 cases, defined as hospitalizations and deaths, were prevented among adults aged 65 years and older, accounting for 123,324 hospitalizations and 16,367 deaths. These figures represent 89.8% of all hospitalizations and 91.1% of all deaths, respectively. Second dose coverage has been low, but use of semi-annual dosing in those 65 years and older is estimated to increase the number averted by an additional 37,939 symptomatic infections, 1,632 hospitalizations, and 217 deaths in the overall target population of 12 to 64 high-risk and 65 years and older compared to the base-case strategy of annual vaccination for all ages.

For the broader public health impact, mRNA-1283 compared to no vaccine was estimated to prevent 252,892 hospital bed days during crowded conditions and the number of antibiotic prescriptions avoided in the outpatient setting with use of mRNA-1283 compared to no vaccine is 167,896, with 83% of the antibiotic prescriptions avoided being among those 65 years and older.

Given this clinical impact, annual dosing with mRNA-1283 in the target population (base-case analysis) is expected to lead to a gain of 171,150 QALYs compared to no vaccine. Although vaccination costs $12 billion, mRNA-1283 prevents $9.3 billion in costs due to reductions in COVID-19 infection-related costs and lost productivity savings compared to no vaccine. The incremental cost per QALY gained for mRNA-1283 compared to no vaccine is therefore $16,247 (Table 6). In the subgroup of adults aged ≥65 years, mRNA-1283 is associated with 132,619 fewer QALYs lost and $91.1 million in cost savings compared to no vaccine, resulting in mRNA-1283 dominating no vaccine in this population, while the incremental cost per QALY gained for high risk 12 – 64 years olds was $74,530 (See Supplement Table 34). In the scenario of the target population including semi-annual dosing for those aged ≥65 years, vaccine costs increase to $13.2 billion; mRNA-1283 is associated with $3.7 billion in additional costs and 173,188 QALYs gained for an ICER of $21,459 per QALY gained relative to no vaccine.

**Table 6.** Scenario analysis results (mRNA-1283 compared to no vaccine in the target population)

| Model Parameter | Range/Data Selections | ICER (Δ Cost per QALY Gained)) | % Change from Base-Case |
| --- | --- | --- | --- |
| Base-Case | -- | \$16,247 | Reference |
| Premature mortality productivity losses | Includes both market and non-market productivity losses | mRNA-1283 Dominates | -- |
| Perspective | Healthcare | \$30,886 | 90.1% |
| Premature mortality productivity losses | Excludes both market and non-market productivity losses | \$29,171 | 79.6% |
| VE | rVE against hospitalization = rVE against infection | \$26,018 | 60.1% |
| Vaccine coverage rates | Increase coverage by 10% in September for all ages | \$18,046 | 11.1% |
| Vaccine coverage rates | Increase coverage by 5% in September for all ages | \$17,267 | 6.3% |
| Percentage hospitalized (high-risk) | Increased RR of hospitalization (RR=1.45) for high-risk | \$16,566 | 2.0% |
| Inpatient mortality (high-risk) | Increased mortality (RR=1.62) for high-risk | \$16,491 | 1.5% |
| Percentage with outpatient care (high-risk) | Assume equal to general population | \$16,454 | 1.3% |
| Caregiver productivity losses | Excluded | \$16,274 | 0.2% |
| VE | VE against symptomatic COVID-19 infection for adults ages 18 years and older from ICATT study | \$14,154 | -12.9% |
| Vaccine coverage rates | Increase coverage by 5% in September for ages $\geq 65$ only | \$14,085 | -13.3% |
| Opportunity costs | Include opportunity costs | \$14,059 | -13.5% |
| Vaccine coverage rates | Increase coverage by 10% in September for ages $\geq 65$ only | \$12,256 | -24.6% |
| Indirect benefit | Based on CDC Right Study | \$9,653 | -40.6% |
| Percentage with no formal care | Based on influenza (2023 - 2024) | \$7,094 | -56.3% |

Abbreviations: $\Delta$ , difference; ICER, incremental cost-effectiveness ratio; QALY, quality-adjusted life-year; RR: relative risk; VE: vaccine effectiveness

Vaccinating with mRNA-1283 generates at least a return on investment of $2.16 per one dollar spent. The BCR increases to $9.74 per dollar spent for mRNA-1283 COVID-19 vaccination as the method of monetizing health benefit is varied. (See Supplement Table 50 for all BCRs for base case and the scenario analyses with semi-annual dosing.)

### Deterministic Sensitivity Analyses (mRNA-1283 compared to no vaccine)

The impact of deterministic sensitivity analyses (DSAs) on the number of symptomatic infections, hospitalizations, and deaths prevented is summarized in the Technical Appendix (Supplemental Table 37); the impact of key DSAs on cost-effectiveness results is presented in Figure 3 (details in Supplemental Table 38). Varying the COVID-19 incidence has the greatest impact on the ICER; varying COVID-19 incidence to the lower bound estimates based on the 2024-2025 season yields an ICER of $51,113 per QALY gained relative to no vaccine (214.7% change from the base-case ICER), while varying to the upper bound estimates based on the 2023-2024 season leads to mRNA-1283 dominating no vaccine. Varying the proportion of patients with symptomatic infection who are hospitalized also has a large impact on the ICER; varying to −25% and +25% of the base-case values yields ICERs of $31,826 (95.9%) and $6,100 (−62.5%) per QALY gained, respectively. Varying post-discharge mortality to the 95% CI upper bound decreases the ICER to $8,784 (45.9% decrease), while the 95% CI lower bound increases the ICER to $29,794 (83.4%). Varying the mRNA-1283 initial VE against hospitalization based on the 95% CI lower and upper bounds led to ICERs of $28,493 (75.4% increase) and $8,301 (48.9% decrease), respectively. Other key variables impacting the cost-effectiveness results by >20% include the vaccine administration cost, the mRNA-1283 waning rates against infection and hospitalization, post-discharge costs, mRNA-1283 initial VE against infection, and hospitalization costs. Vaccine coverage also impacts cost-effectiveness results by 20%, as VCRs impact clinical outcomes due to the heterogeneity in model age groups (i.e., changing coverage rates in model age groups will affect clinical outcomes).

**Figure 3:**
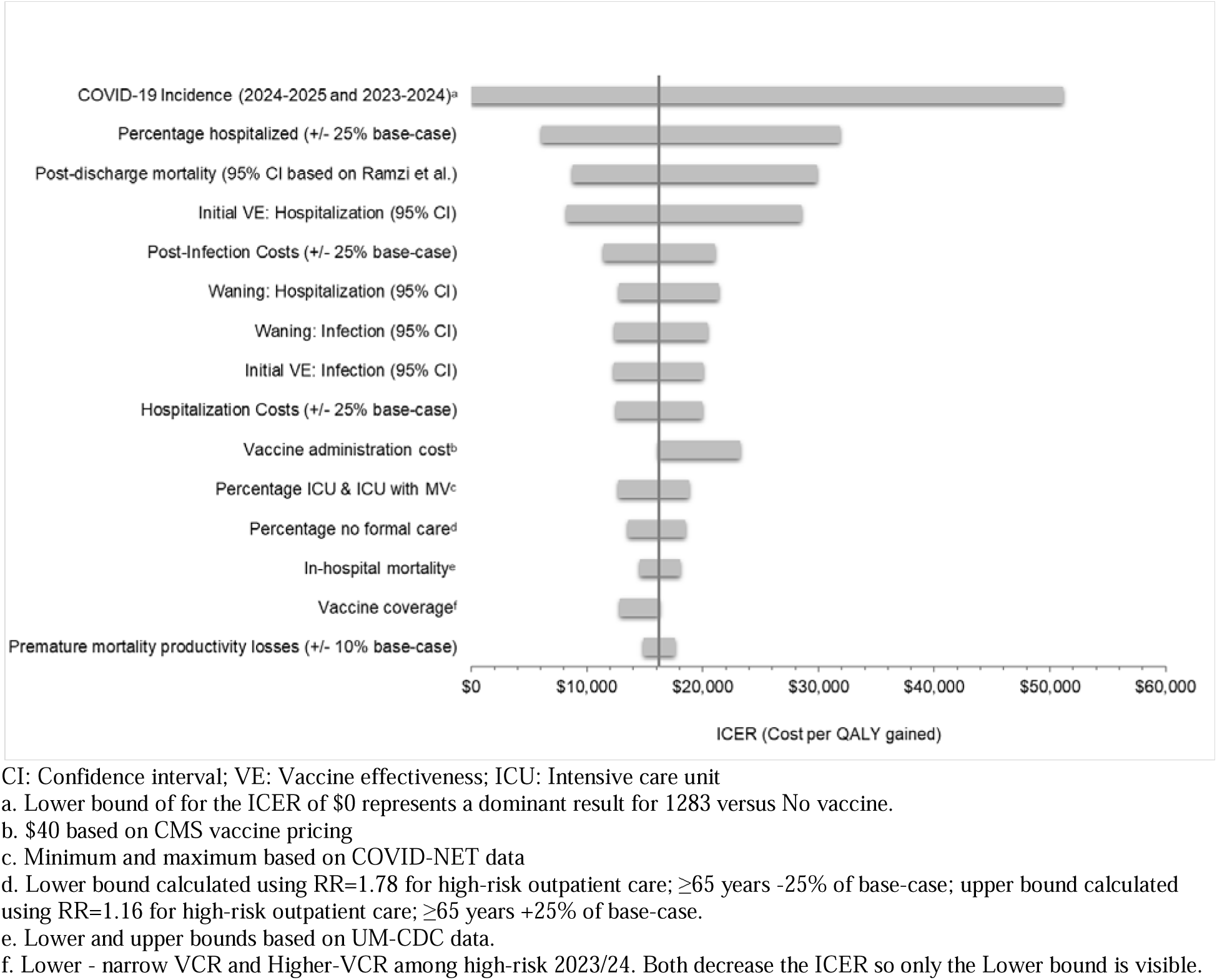
Tornados: Impact of deterministic sensitivity analyses on the base-case cost-effectiveness results (mRNA-1283 relative to no vaccine)

### Scenario Analyses (mRNA-1283 compared to no vaccine)

The results of the impact of scenario analyses on infections, hospitalizations, and deaths in the base-case target population are shown in the Technical Appendix; the economic impact in the comparison of mRNA-1283 to no vaccine is depicted in the table below (Table 6). Premature mortality productivity losses were shown to have a large impact on the ICER; including both non-market and market productivity losses leads to mRNA-1283 dominating no vaccine, while excluding both market and non-market premature mortality productivity losses leads to a 79.5% increase in the ICER (compared to the base-case) to $29,171 per QALY gained. An analysis performed from the healthcare perspective (excluding both patient and caregiver lost productivity) yielded an ICER of $30,886 per QALY gained, representing a 90% increase from the base-case. Assuming the rVE for hospitalization is equivalent to the rVE against infection (i.e., there is no additional benefit against hospitalization) increases the ICER by 60% to $26,018 per QALY gained. Basing the proportion of patients with no formal care for COVID-19 related-illness on CDC burden of illness influenza data from the 2023-2024 season leads to a 56% reduction in the ICER; this is largely due to the percentage with no formal care being lower in the influenza data compared to the base-case analysis. In the scenario analysis examining indirect benefit, the ICER is reduced to $9,653 (40.6% decrease).

### Comparison against originally licensed mRNA COVID-19 vaccines mRNA-1273 and BNT162b2

Clinical outcomes for the comparisons between mRNA-1283 and mRNA-1273 and mRNA-1283 and BNT162b2 are presented in the Technical Appendix (Supplemental Tables 32 and 33).

In the target population, mRNA-1283 is expected to avert an additional 427,337 symptomatic COVID-19 infections, 36,263 COVID-19 related hospitalizations and 4,725 deaths compared to mRNA-1273. This impact on clinical outcomes leads to reduced total costs for the mRNA-1283 vaccination strategy compared to mRNA-1273, and fewer QALYs lost, leading to mRNA-1283 dominating mRNA-1273 in the base-case analysis for both the target population and the ≥65 years subgroup (Table 6; Supplemental Table 35). In the 12-64 high risk group the incremental cost per QALY gained was $14,206. Compared to BNT162b2, mRNA-1283 is expected to avert 606,942 symptomatic infections, 46,176 hospitalizations, and 6,023 deaths, leading to reduced total costs and fewer QALYs lost for the mRNA-1283 vaccination strategy compared to BNT162b2. As a result, mRNA-1283 dominates BNT162b2 in the base-case analysis for both the target population and the ≥65 years and 12-64 high risk subgroups (Table 6; Supplemental Table 43). The BCR for the base case target population for mRNA-1283 relative to mRNA-1273 ranged from 3.14 to 14.11, while the BCR for mRNA-1283 relative to BNT162b2 ranged from 4.81 to 20.91.

**Table 6.** Cost-effectiveness results compared to originally licensed mRNA COVID-19 vaccines.

| Vaccination strategy | Total Costs | Total QALYs Lost | Δ Costs | Δ QALYs Gained* | ICER (Δ Cost per QALY Gained) |
| --- | --- | --- | --- | --- | --- |
| **mRNA-1283 compared to mRNA-1273** |  |  |  |  |  |
| **Base-Case: Annual dosing for 12-64 High-Risk, ≥65 All** |  |  |  |  |  |
| mRNA-1283 | \$67,360,095,079 | 846,729 | -- | -- | Reference |
| mRNA-1273 | \$67,522,322,919 | 890,423 | \$162,227,840 | -43,694 | mRNA-1283 Dominates |
| **Scenario Analysis: Annual dosing for 12-64 High-Risk, Semi-annual dosing for ≥65 All** |  |  |  |  |  |
| mRNA-1273 | \$68,270,475,283 | 888,344 | -- | -- | Reference |
| mRNA-1283 | \$68,295,993,219 | 844,691 | \$25,517,936 | 43,653 | \$585 |
| **mRNA-1283 compared to BNT162b2** |  |  |  |  |  |
| **Base-Case: Annual dosing for 12-64 High-Risk, ≥65 All** |  |  |  |  |  |
| mRNA-1283 | \$67,360,095,079 | 846,729 | -- | -- | Reference |
| BNT162b2 | \$68,598,502,341 | 903,582 | \$1,238,407,263 | -56,854 | mRNA-1283 Dominates |
| **Scenario Analysis: Annual dosing for 12-64 High-Risk, Semi-annual dosing for ≥65 All** |  |  |  |  |  |
| mRNA-1283 | \$68,295,993,219 | 844,691 | -- | -- | Reference |
| BNT162b2 | \$69,375,942,695 | 901,511 | \$1,079,949,475 | -56,820 | mRNA-1283 Dominates |

Scenario analyses for the comparisons between mRNA-1283 and mRNA-1273 and mRNA-1283 and BNT162b2 in the target population are presented in the Technical Appendix (Supplemental Tables 32 and 33). Varying the COVID-19 incidence to the 2024-2025 season yielded an ICER of $20,397 per QALY gained for mRNA-1283 compared to mRNA-1273. Varying the rVEs for infection and hospitalization to their respective lower bounds led to ICERs of $9,907 and $77,473 per QALY gained for mRNA-1283 relative to mRNA-1273, respectively. In all other scenarios, mRNA-1283 dominated mRNA-1273, consistent with the base-case results. In the comparison against BNT162b2, varying the rVE of infection or the rVE of hospitalization to the lower bound led to an ICER of $5,793 and $80,229 per QALY gained respectively for mRNA-1283 relative to BNT162b2. In all other scenarios, consistent with the base-case analysis, mRNA-1283 dominated BNT162b2.

## Discussion

During the 2025/2026 season in the United States, we estimate that the next-generation Moderna COVID-19 mRNA-1283 vaccine would prevent, in its licensed target population (adults aged ≥65 years and individuals aged 12–64 years with underlying medical conditions), approximately 2.2 million symptomatic COVID-19 infections, 137,391 hospitalizations, and 17,960 deaths compared to no vaccination. Given this clinical impact, annual dosing with mRNA-1283 in the target population (base-case analysis) is expected to lead to a gain of 171,150 QALYs with incremental costs of $2.8 billion compared to no vaccine, yielding an ICER (cost per QALY gained) of $16,247 for mRNA-1283 compared to no vaccine. In the subgroup of all adults aged ≥65 years, mRNA-1283 was shown to dominate no vaccine. In the scenario analysis examining a second dose for those aged ≥65 years, the ICER for mRNA-1283 was estimated to be $21,459 per QALY gained relative to no vaccine.

Compared with the originally approved mRNA-1273 vaccine we estimate that mRNA-1283 would have an additional 427,337 symptomatic infections, 36,263 hospitalizations, and 4,725 deaths. Compared to BNT162b2, we estimate that mRNA-1283 would have averted an additional 606,942 symptomatic infections, 46,176 hospitalizations, and 6,023 deaths. Given these additional clinical outcomes averted, mRNA-1283 was shown to dominate both mRNA-1273 and BNT162b2 in the analysis target population.

Based on the base-case analysis, for every 1 USD of mRNA-1283 vaccine related costs, there is a return of 2.16 to 9.74 dollars in terms of costs from the societal perspective and monetized health benefit gained by preventing SARS-CoV-2 infections and related COVID-19 outcomes. The range depends on how health benefit is measured and monetized: at the low end a QALY gained is valued at $100,000 while at the high end it was $717,000. In the comparison with mRNA-1273 there is a return of 3.14 to 14.11 dollars and with BNT162b2, there is a return of 4.81 to 20.91 dollars.

These results suggest that broad vaccination of 12 to 64 high-risk and all ≥65 years, even in the post-pandemic era, against COVID-19 has substantial public health impact, is highly cost-effective and provides a strong economic return to society. This finding is not only important for increasing VCRs, but also decision making as to which vaccine will yield high economic return, an important consideration for informing optimal vaccination policy. Combined with multiple scenario and sensitivity analyses confirming the robustness of the current model, the results suggest that mRNA-1283 represents a valuable option to optimize US COVID-19 vaccination strategies—particularly for older adults (≥65 years) and individuals with high-risk medical conditions who remain most vulnerable to severe disease. Furthermore, the differences in COVID-19 outcomes prevented by mRNA-1283 compared to each mRNA-1273 and BNT162b2 underscore that the choice of each mRNA COVID-19 vaccine likely result in substantial differences in clinical impact.

These estimates are consistent with findings from a recent Canadian analysis conducted with a similar decision analytic model showing a large reduction in COVID-19 infections and severe COVID-19 cases with mRNA-1283 when compared to no vaccination and substantial additional COVID-19 events prevented when compared to current COVID-19 mRNA vaccines.^65^ Further, the study concluded that mRNA-1283 represented good value for money if priced below $230 (CAD) per dose under a range of assumptions around VE, COVID-19 incidence, and vaccine coverage. In the September 2025 ACIP meeting, the University of Michigan COVID-19 Vaccination Modeling Team presented an updated economic analysis^61^ of a generic COVID-19 mRNA booster compared to no updated mRNA booster in US adults (aged ≥18 years) stratified by risk status, finding that vaccination has a substantial impact on clinical outcomes;^61^ the presentation also noted that cost-effectiveness results for those ages ≥65 years were robust to variations in input parameters, and that vaccination can be considered cost-effective under certain conditions for those ages 18-49 and 50-64 at high risk of severe outcomes from COVID-19.^61^

The cost-effectiveness analysis presented in this manuscript has several limitations. First, the real-world effectiveness and safety of mRNA-1283 have not yet been established. The VE input was based on interim vaccine effectiveness estimates for mRNA-1273 targeting the KP.2 variant, combined with the rVE between mRNA-1283 and mRNA-1273. The rVE estimate for hospitalization was derived from a post-hoc analysis of FDA-defined severe COVID-19, which only included a total of 55 FDA-defined severe COVID-19 events with the majority of cases due to blood pressure and oxygen saturation abnormalities. Additionally, initial rVE estimates for mRNA-1283 versus BNT162b2 were based on indirect treatment comparisons^14^ and should be validated and refined with real-world head-to-head data.

The model assumed equal waning across all three vaccines, which may lead to conservative estimates for mRNA-1283, particularly in older adults, given that the Phase 3 trial findings show higher immunogenicity for mRNA-1283 than mRNA-1273 at all matched timepoints during follow-up^66^.

For model calibration, an average of 2023/2024 and 2024/2025 season CDC-reported hospitalization rates were used as the base case to address uncertainty similar to approaches for seasonal influenza vaccines; however, substantial uncertainty remains regarding future COVID-19 incidence.

It is also unclear how the ACIP recommendation for the 2025/2026 COVID-19 vaccination program will impact vaccine coverage. Finally, the model does not fully capture the impact of vaccination on downstream outcomes from COVID-19 infections, particularly related to the triggering and worsening of chronic medical conditions such as cardiovascular, chronic lung, kidney disease and diabetes.

## Conclusions

This assessment suggests that the next-generation COVID-19 mRNA-1283 vaccine could substantially reduce the clinical and economic burden of COVID-19 among high-risk populations in the US—specifically, individuals aged 12–64 years with underlying conditions and all adults aged ≥65 years. These benefits were observed both in comparison to no vaccination and to the originally approved mRNA vaccines. As such, mRNA-1283 represents a valuable option for optimizing COVID-19 vaccination policy in the US for those at highest risk of severe outcomes.

## Transparency

- **Declaration of Funding:** This study was supported by Moderna, Inc.
- **Declaration of competing interests:** MK is a shareholder in Quadrant Health Economics Inc, which was contracted by Moderna, Inc. to conduct this study. AL, KF, MW and SC are consultants to Quadrant Health Economics Inc. KJ, NV, EB and MB are employed by Moderna, Inc. and may hold stock/stock options in the company.

## Data Availability

All data produced in the present work are contained in the manuscript

## References

1. Centers for Disease Control and Prevention. Preliminary Estimates of COVID-19 Burden for 2024-2025. https://www.cdc.gov/covid/php/surveillance/burden-estimates.html. Accessed May 19, 2025.

2. MacNeil A. Evidence to Recommendations (partial) for 2025–2026 COVID-19 Vaccination. Presentation to the Advisory Committee on Immunization Practices. https://www.cdc.gov/acip/downloads/slides-2025-06-25-26/06-MacNeil-COVID-508.pdf. Accessed June 25, 2025.

3. Centers for Disease Control and Prevention. Updates to COVID-19 epidemiology. Advisory Committee on Immunization Practices. September 19, 2025. https://www.cdc.gov/acip/downloads/slides-2025-09-18-19/02-Srinivasan-covid-508.pdf. Accessed.

4. Centers for Disease Control and Prevention. COVID-19 Vaccination Coverage and Intent for Vaccination, Adults 18 Years and Older, United States. https://www.cdc.gov/covidvaxview/weekly-dashboard/adult-vaccination-coverage.html. Accessed May 19, 2025.

5. Centers for Disease Control and Prevention. Flu Vaccination Coverage, United States, 2023–24 Influenza Season. https://www.cdc.gov/fluvaxview/coverage-by-season/2023-2024.html. Accessed May 19, 2025.

6. Food and Drug Administration. MNEXSPIKE. https://www.fda.gov/vaccines-blood-biologics/mnexspike. Accessed July 7, 2025.

7. Center for Disease Control and Prevention. Vaccines & Immunizations. Adult Immunization Schedule Appendix. https://www.cdc.gov/vaccines/hcp/imz-schedules/adult-appendix.html. Accessed November 6, 2025.

8. Chalkias S, Dennis P, Petersen D, et al. Efficacy, immunogenicity, and safety of a next-generation mRNA-1283 COVID-19 vaccine compared with the mRNA-1273 vaccine: results from NextCOVE, a phase 3, randomised, observer-blind, active-controlled trial. The Lancet Infectious diseases. 2025.

9. Chheda U, Pradeepan S, Esposito E, Strezsak S, Fernandez-Delgado O, Kranz J. Factors Affecting Stability of RNA - Temperature, Length, Concentration, pH, and Buffering Species. J Pharm Sci. 2024;113(2):377–385.

10. Stewart-Jones GBE, Elbashir SM, Wu K, et al. Domain-based mRNA vaccines encoding spike protein N-terminal and receptor binding domains confer protection against SARS-CoV-2. Sci Transl Med. 2023;15(713):eadf4100.

11. Kopel H, Nguyen VH, Boileau C, et al. Comparative effectiveness of Bivalent (Original/Omicron BA.4/BA.5) mRNA COVID-19 Vaccines mRNA-1273.222 and BNT162b2 Bivalent Vaccine in Adults in the US. Vaccines. 2023.

12. Kavikondala S, Haeussler K, Wang X, et al. Comparative Effectiveness of mRNA-1273 and BNT162b2 COVID-19 Vaccines Among Older Adults: Systematic Literature Review and Meta-Analysis Using the GRADE Framework. Infect Dis Ther. 2024.

13. Wang X, Pahwa A, Bausch-Jurken MT, et al. Comparative Effectiveness of mRNA-1273 and BNT162b2 COVID-19 Vaccines Among Adults with Underlying Medical Conditions: Systematic Literature Review and Pairwise Meta-Analysis Using GRADE. Advances in Therapy. 2025;42(5):2040–2077.

14. Beck E, Georgieva M, Wang WJ, et al. Indirect comparison of the relative vaccine effectiveness of mRNA-1283 vs. BNT162b2 vaccines against symptomatic COVID-19 among US adults. Curr Med Res Opin. 2025;41(4):721–732.

15. US Department of Health and Human Services. ACIP Recommends COVID-19 Immunization Based on Individual Decision-making.. https://www.hhs.gov/press-room/acip-recommends-covid19-vaccination-individual-decision-making.html. Accessed.

16. Fust K, Kohli M, Joshi K, et al. Modeling the Anticipated Public Health Benefits of the Next-Generation COVID-19 mRNA-1283 vaccine: An Interim U.S. Population-Level Impact Assessment (pre-print). Available from: https://www.medrxiv.org/content/10.1101/2025.09.26.25336704v1. Last accessed 23 Oct 2025.

17. Prosser LA, Wallace M, Rose AM, et al. Cost-Effectiveness of 2023-2024 COVID-19 Vaccination in US Adults. JAMA Network Open. 2025;8(8):e2523688–e2523688.

18. University of Michigan COVID-19 Vaccination Modeling Team. Economic analysis of an additional dose of the 2024-2025 COVID-19 vaccine. Presentation to the Advisory Committee on Immunization Practices. Centers for Disease Control October 23, 2024.

19. Center for Disease Control and Prevention. Vaccines & Immunizations. Adult Immunization Schedule by Age (Addendum updated August 7, 2025). https://www.cdc.gov/vaccines/hcp/imz-schedules/adult-age.html. Accessed November 6, 2025.

20. Roper LE, Godfrey M, Link-Gelles R, et al. Use of Additional Doses of 2024-2025 COVID-19 Vaccine for Adults Aged. MMWR Morb Mortal Wkly Rep.73(49):1118–1123.

21. Neumann PJ, Kim DD. Cost-effectiveness Thresholds Used by Study Authors, 1990-2021. JAMA. 2023;329(15):1312–1314.

22. ASPE Office of Science and Data Policy. HHS Standard Values for Regulatory Analysis, 2025. https://aspe.hhs.gov/sites/default/files/documents/639756a60fbe7e51786bcec176ad52f1/Standard-RIA-Values-2025.pdf. Accessed.

23. United Nations Department of Economic and Social Affairs (Population Division). World Population Prospects 2024, Online Edition. https://population.un.org/wpp/downloads?folde=Standard%20Projections&group=Population. Published 2024. Accessed September 3, 2024.

24. Kompaniyets L, Agathis NT, Nelson JM, et al. Underlying Medical Conditions Associated With Severe COVID-19 Illness Among Children. JAMA Netw Open. 2021;4(6):e2111182.

25. Panagiotakopoulos L. Use of 2025–2026 COVID-19 Vaccines: Work Group Considerations. Presentation to the Advisory Commitee on Immunization Practices, April 15, 2025. https://www.cdc.gov/acip/downloads/slides-2025-04-15-16/05-Panagiotakopoulos-COVID-508.pdf. Accessed May 5, 2025.

26. Centers for Disease Control and Prevention. COVID-NET. COVID-19 hospitalization surveillance network. https://www.cdc.gov/covid/php/covid-net/index.html?CDC_AA_refVal=https://3A%2F%2Fwww.cdc.gov%2Fcoronavirus%2F2019-ncov%2Fcovidnetdashboard%2Fde%2Fpowerbi%2Fdashboard.html. Published 2024. Accessed May 6, 2024.

27. Kim DeLuca E, Gebremariam A, Rose A, Biggerstaff M, Meltzer MI, Prosser LA. Cost-effectiveness of routine annual influenza vaccination by age and risk status. Vaccine. 2023;41(29):4239–4248.

28. Centers for Disease Control and Prevention. Weekly Cumulative COVID-19 VACCINATION Coverage and Intent, Overall, by Selected Demographics and Jurisdiction, Among Adults 18 Years and Older. https://data.cdc.gov/Vaccinations/Weekly-Cumulative-COVID-19-Vaccination-Coverage-an/ksfb-ug5d/about_data. Published 2025. Accessed February 28, 2025.

29. Centers for Disease Control and Prevention. Weekly Parental Intent for Vaccination and Cumulative Percentage of Children 6 Months −17 Years Who are Up to date with the COVID-19 Vaccines by Season, United States. https://www.cdc.gov/covidvaxview/weekly-dashboard/child-coverage-vaccination.html. Published 2025. Accessed February 28, 2025.

30. County of Los Angeles Public Health. COVID-19 Vaccine: Vaccinations in LA County. http://publichealth.lacounty.gov/media/coronavirus/vaccine/vaccine-dashboard.htm. Accessed November 6, 2025.

31. Moderna Inc. mRNA-1283 P301 Clinical Study Report.

32. Wilson A, Bogdanov A, Zheng Z, et al. Evaluating the Effectiveness of 2024-2025 Seasonal mRNA-1273 Vaccination Against COVID-19-Associated Hospitalizations and Medically Attended COVID-19 among adults aged ≥18 years in the United States. *medRxiv.* 2025;2025.03.27.25324770.

33. MacNeil A. Updates to COVID-19 Vaccine Effectiveness. Presentation to the Advisory Committee on Immunization Practices. June 25. 2025. https://www.cdc.gov/acip/downloads/slides-2025-06-25-26/03-MacNeil-COVID-508.pdf. Accessed June 25, 2025.

34. Kopel H, Nguyen VH, Bogdanov A, et al. Comparative Effectiveness of the Bivalent (Original/Omicron BA.4/BA.5) mRNA COVID-19 Vaccines mRNA-1273.222 and BNT162b2 Bivalent in Adults with Underlying Medical Conditions in the United States. Vaccines (Basel). 2024;12(10).

35. Beck E, Georgieva M, Wang W, et al. An Indirect Treatment Comparison of COVID-19 Next Generation mRNA-1283 Vaccine and BNT162b2 Vaccine Against COVID-19 Symptomatic Infections in the US. Paper presented at: AMCP2025; Houston, Texas, USA.

36. Higdon MM, Baidya A, Walter KK, et al. Duration of effectiveness of vaccination against COVID-19 caused by the omicron variant. The Lancet Infectious diseases. 2022;22(8):1114–1116.

37. Andersson NW, Thiesson EM, Pihlström N, et al. Comparative effectiveness of monovalent XBB.1.5 containing covid-19 mRNA vaccines in Denmark, Finland, and Sweden: target trial emulation based on registry data. BMJ Med. 2024;3(1):e001074.

38. Hansen CH, Lassaunière R, Rasmussen M, Moustsen-Helms IR, Valentiner-Branth P. Effectiveness of the BNT162b2 and mRNA-1273 JN.1-adapted vaccines against COVID-19-associated hospitalisation and death: a Danish, nationwide, register-based, cohort study. The Lancet Infectious Diseases. 2025.

39. Zheng Z, Yousefi M, MacComrac D, et al. Effectiveness of 2023-2024 mRNA-1273 XBB.1.5 Vaccine Against Covid-19 Associated Hospitalizations and Medically Attended Covid-19 in the United States. Available at SSRN: https://ssrn.com/abstract=5258507 or 10.2139/ssrn.5258507. 2025.

40. University of Michigan COVID-19 Vaccination Modeling Team. Economic analysis of COVID-19 vaccination. https://www.cdc.gov/acip/downloads/slides-2024-06-26-28/05-COVID-Prosser-508.pdf. Published 2024. Updated June 24 2024. Accessed February 19, 2025.

41. Joshi K, Dronova M, Paterak E, et al. Clinical Impact and Cost-Effectiveness of Updated 2023/24 COVID-19 mRNA Vaccination in High-Risk Populations in the United States. Infect Dis Ther. 2025.

42. Ramzi ZS. Hospital readmissions and post-discharge all-cause mortality in COVID-19 recovered patients; A systematic review and meta-analysis. Am J Emerg Med. 2022;51:267–279.

43. Kopel H, Araujo AB, Bogdanov A, et al. Effectiveness of the 2023-2024 Omicron XBB.1.5-containing mRNA COVID-19 vaccine (mRNA-1273.815) in preventing COVID-19-related hospitalizations and medical encounters among adults in the United States: An interim analysis. *medRxiv*. 2024:2024.2004.2010.24305549.

44. Moderna data on file 2024. Optum’s de-identified Clinformatics® Data Mart Database. Analysis September 2023 to February 2024 COVID-19 medical attendances and hospitalizations.

45. Moderna. COVID-19 Hospitalization Risk by Comorbidities Profile (Risk Stacking). Moderna bench to practice website. https://dev.atlas.modernatx.com/bench2practice/Interactive-dashboard. Accessed.

46. Mercon KR, Rose AM, Cadham CJ, et al. Health Preferences in Transition: Differences from Pandemic to Post-Pandemic in Valuation of COVID-19 and RSV Illness in Children and Adults. Children (Basel). 2025;12(2).

47. PHOSP-COVID Collaborative Group. Clinical characteristics with inflammation profiling of long COVID and association with 1-year recovery following hospitalisation in the UK: a prospective observational study. Lancet Respir Med. 2022;10(8):761–775.

48. Sanders GD, Neumann PJ, Basu A, et al. Recommendations for Conduct, Methodological Practices, and Reporting of Cost-effectiveness Analyses: Second Panel on Cost-Effectiveness in Health and Medicine. JAMA. 2016;316(10):1093–1103.

49. Arias E. United States Life Tables, 2017 (National Vital Statistics Report). 2019.

50. Hanmer J, Lawrence WF, Anderson JP, Kaplan RM, Fryback DG. Report of nationally representative values for the noninstitutionalized US adult population for 7 health-related quality-of-life scores. Med Decis Making. 2006;26(4):391–400.

51. Yehoshua A, Cook AD, Di Fusco M, et al. Health outcomes and economic burden among patients with a COVID-19-associated hospitalization in the United States during the predominance of the XBB and JN.1 omicron lineages. J Med Econ. 2024;27(1):1372–1378.

52. Kapinos KA, Peters RM, Jr, Murphy RE, Hohmann SF, Podichetty A, Greenberg RS. Inpatient Costs of Treating Patients With COVID-19. JAMA Network Open. 2024;7(1):e2350145–e2350145.

53. Chambers LC, Park A, Cole M, et al. Long-term health care costs following COVID-19: implications for pandemic preparedness. Am J Manag Care. 2023;29(11):566–572.

54. Scott A, Ansari W, Khan F, et al. Substantial health and economic burden of COVID-19 during the year after acute illness among US adults at high risk of severe COVID-19. BMC Med. 2024;22(1):46.

55. Scott A, Ansari W, Chambers R, et al. Substantial health and economic burden of COVID-19 during the year after acute illness among US adults not at high risk of severe COVID-19. BMC Med. 2024;22(1):47.

56. Center for Disease Control and Prevention. Vaccines for Children Program. Current CDC Vaccine Price List. https://www.cdc.gov/vaccines-for-children/php/awardees/current-cdc-vaccine-price-list.html. Published 2025. Updated November 1, 2025. Accessed November 3, 2025.

57. Centers for Medicare & Medicaid Services. 2025 National Physician Fee Schedule Relative Value File October Release. Available from: https://www.cms.gov/medicare/payment/fee-schedules/physician/pfs-relative-value-files/rvu25d-0. Last accessed 6 Oct 2025.

58. Centers for Medicare & Medicaid Services. Vaccine Pricing. https://www.cms.gov/medicare/payment/part-b-drugs/vaccine-pricing. Accessed September 19, 2024.

59. Kwon J, Milne R, Rayner C, et al. Impact of Long COVID on productivity and informal caregiving. Eur J Health Econ. 2024;25(7):1095–1115.

60. Grosse SD, Krueger KV, Pike J. Estimated annual and lifetime labor productivity in the United States, 2016: implications for economic evaluations. J Med Econ. 2019;22(6):501–508.

61. University of Michigan COVID-19 Vaccination Modeling Team. Economic Analysis of COVID-19 Vaccination. Presentation to the Advisory Committee on Immunization Practices. September 19, 2025 2025.

62. Ortega-Sanchez IR. Economics of Respiratory Syncytial Virus Vaccination in Adults aged 50 - 59 years at Increased Risk of Severe RSV Disease. https://www.cdc.gov/acip/downloads/slides-2025-04-15-16/05-Ortega-Sanchez-Adult-RSV-508.pdf. Accessed April 16, 2025.

63. Leidner AJ, Bletnitsky, S.,. Summary of three economic analyses on the use of PCVs among 50-64 year old adults in the United States. https://www.cdc.gov/acip/downloads/slides-2024-10-23-24/03-Leidner-Pneumococcal-508.pdf. Published 2024. Accessed May 15, 2025.

64. Link-Gelles R. CDC National Center for Immunization and Respiratory Diseases. Effectiveness of COVID-19 (2023-2024 Formulation). https://www.cdc.gov/vaccines/acip/meetings/downloads/slides-2024-06-26-28/03-COVID-Link-Gelles-508.pdf. Published 2024. Updated June 27, 2024. Accessed.

65. Fust K, Kohli M, Cartier S, et al. The potential impact of the next-generation COVID-19 mRNA-1283 vaccine in Canada. J Med Econ. 2025;28(1):1440–1450.

66. Rizkalla B. Overview of Moderna’s Investigational Next Generation COVID-19 Vaccine, mRNA-1283, in Individuals. https://www.cdc.gov/acip/downloads/slides-2025-04-15-16/02-Rizkalla-COVID-508.pdf. Accessed April 15, 2025.

