## Supplemental File for "Modeling the Cost-Effectiveness of the Next-Generation COVID-19 mRNA-1283 vaccine in the United States"

- Technical Appendix –

### 1 Estimation of Incidence of Infection (No Vaccination)

As the hospitalization data from the CDC reflects a partially vaccinated population, information on vaccination coverage, VE, and the probability of hospitalization must be used as well as the target hospitalization rates. Two calibrations were completed, the first using data from September 2023 to August 2024 and the second for September 2024 to August 2025. The process is described below.

**Step 1**: Develop the age-specific targets for the monthly rate of hospitalizations by age group for a one-year period. As hospitalization rates for one full year, i.e. 12 months, are needed, for the base case, the hospitalization rates for September 2023 to August 2024 and September 2024 to August 2025 from COVID-NET^[1]^ were used.^1^ The rates per 100,000 are displayed in Table 1 and Table 2 below.

Table 1. Target hospitalization rates per 100,000 for the calculation of infection incidence in the model (September 2023 – August 2024)^1^

| **Age Group (Years)** | **Sep-23** | **Oct-23** | **Nov-23** | **Dec-23** | **Jan-24** | **Feb-24** | **Mar-24** | **Apr-24** | **May-24** | **Jun-24** | **Jul-24** | **Aug-24** |
| --- | --- | --- | --- | --- | --- | --- | --- | --- | --- | --- | --- | --- |
| 12-17 | 1.8 | 1.0 | 1.7 | 2.6 | 2.8 | 2.7 | 1.4 | 0.4 | 0.6 | 0.4 | 1.3 | 1.7 |
| 18-49 | 5.5 | 5.2 | 6.1 | 8.3 | 9 | 5.7 | 2.9 | 1.6 | 1.6 | 2.6 | 5.3 | 6.7 |
| 50-64 | 15.9 | 15.2 | 18.8 | 24 | 26.4 | 16.2 | 8.8 | 5 | 4.4 | 6.4 | 13 | 17.2 |
| ≥ 65 | 80.9 | 80.7 | 91.1 | 123.2 | 113.3 | 68.8 | 43 | 25.7 | 24.4 | 39.3 | 66.5 | 82.5 |

Table 2. Target hospitalization rates per 100,000 for the calculation of infection incidence in the model (September 2024 – August 2025)^1^

| **Age Group (Years)** | **Sep-24** | **Oct-24** | **Nov-24** | **Dec-24** | **Jan-25** | **Feb-25** | **Mar-25** | **Apr-25** | **May-25** | **Jun-25** | **Jul-25** | **Aug-25** |
| --- | --- | --- | --- | --- | --- | --- | --- | --- | --- | --- | --- | --- |
| 12-17 | 1.7 | 1.1 | 0.6 | 0.9 | 0.9 | 0.8 | 0.8 | 0.6 | 0.3 | 0.2 | 0.6 | 1.1 |
| 18-49 | 4.3 | 2.6 | 1.9 | 3.1 | 3.9 | 2.9 | 2.4 | 1.6 | 1.2 | 1.1 | 1.9 | 3.8 |
| 50-64 | 13.2 | 8.7 | 6.2 | 9.1 | 12.5 | 9.1 | 7 | 4.3 | 3 | 2.5 | 4.1 | 7 |
| ≥ 65 | 66.3 | 46.8 | 30.8 | 54.4 | 59.9 | 40.9 | 35.5 | 24.4 | 17.6 | 14.9 | 18.4 | 33.2 |

**Step 2**: Enter the age-specific probability of hospitalization and the related proportion of symptomatic cases that are seeking care. The same probabilities are used for the cost-effectiveness model of the symptomatic infection in the no vaccination arm and the incidence calculation.

**Step 3**: Enter the assumed vaccination coverage.

The monthly vaccine coverage rates for September 2023 to August 2024 (Table 3) and September 2024 to August 2025 were estimated from the vaccine coverage from the CDC VaxView database.^2^ Data on coverage of a second dose of the vaccine were also available for individuals aged 65 years and older, and began April 1, 2024 and end July 27, 2024. Figure 1 provides detailed vaccine coverage over time for the second doses.

A small proportion of people aged 65 years and older in the US were vaccinated with a second dose between April and July 2024 (See Figure 1). The model only accommodates one dose for the calculations of infection incidence, so the single dose coverage rates were increased to account for second doses in spring 2024 and spring 2025. Overall, excluding these vaccinations or including them made a small difference to the final infection incidence rates.

Figure 1. Second dose, COVID-19 vaccine coverage during season 2023-2024

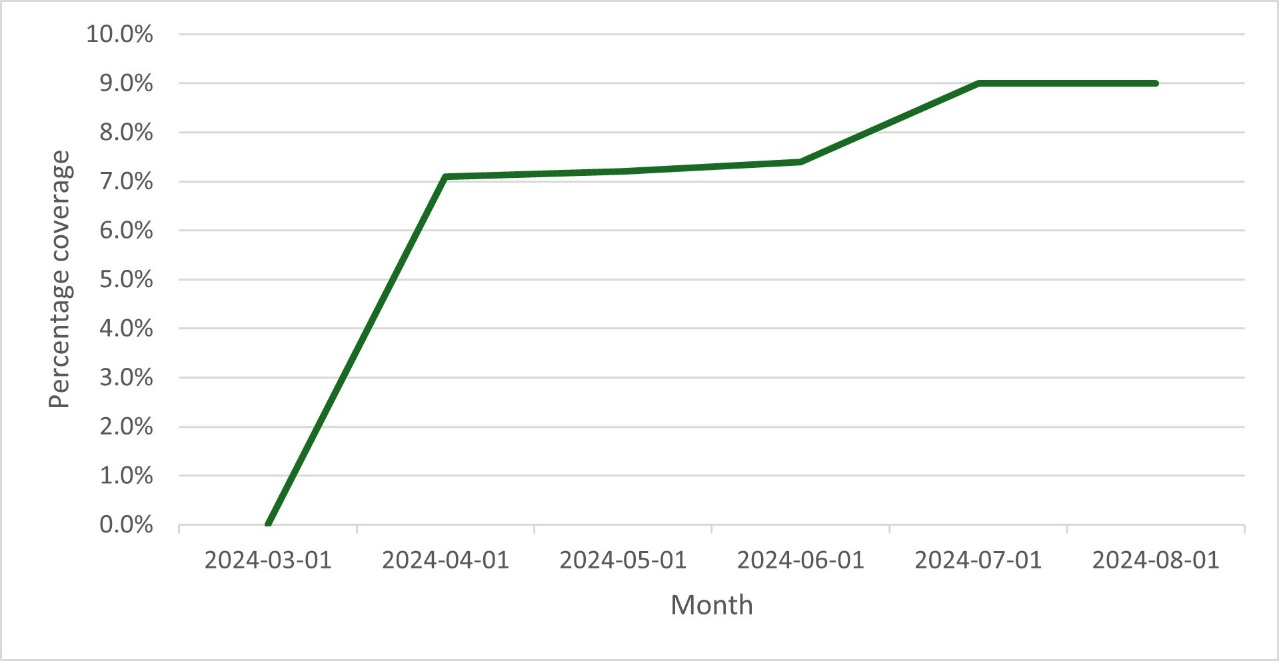

Table 3. Monthly coverage rates (September 2023 – August 2024) used in the calculation of infection incidence in the static model

| **Date** | **12-17 years** | **18-49 years** | **50-64 years** | **65+ years** |
| --- | --- | --- | --- | --- |
| 30-Sep-2023 | 0.9% | 1.4% | 3.9% | 6.1% |
| 31-Oct-2023 | 7.6% | 5.6% | 12.3% | 22.5% |
| 30-Nov-2023 | 12.3% | 9.7% | 18.0% | 28.9% |
| 31-Dec-2023 | 14.2% | 11.3% | 20.7% | 32.2% |
| 31-Jan-2024 | 15.7% | 12.9% | 22.7% | 35.1% |
| 29-Feb-2024 | 16.6% | 13.5% | 23.4% | 36.1% |
| 31-Mar-2024 | 17.2% | 13.9% | 23.7% | 37.7% |
| 30-Apr-2024 | 17.6% | 14.2% | 24.2% | 45.4% |
| 31-May-2024 | 18.1% | 14.3% | 24.3% | 45.8% |
| 30-Jun-2024 | 18.1% | 14.3% | 24.3% | 46.0% |
| 31-Jul-2024 | 18.1% | 14.3% | 24.3% | 47.6% |
| 31-Aug-2024 | 18.1% | 14.3% | 24.3% | 47.6% |

Table 4. Monthly coverage rates (September 2024 – August 2025) used in the calculation of infection incidence in the static model

| **Date** | **12-17 years** | **18-49 years** | **50-64 years** | **65+ years** |
| --- | --- | --- | --- | --- |
| 30-Sep-2023 | 4.1% | 3.3% | 7.5% | 15.8% |
| 31-Oct-2023 | 9.1% | 8.4% | 15.7% | 31.2% |
| 30-Nov-2023 | 10.8% | 10.9% | 20.3% | 38.0% |
| 31-Dec-2023 | 13.3% | 12.5% | 22.8% | 42.2% |
| 31-Jan-2024 | 14.5% | 13.6% | 24.0% | 44.0% |
| 29-Feb-2024 | 14.7% | 14.1% | 24.8% | 44.4% |
| 31-Mar-2024 | 14.7% | 14.1% | 24.8% | 44.4% |
| 30-Apr-2024 | 14.7% | 14.1% | 24.8% | 51.5% |
| 31-May-2024 | 14.7% | 14.1% | 24.8% | 51.6% |
| 30-Jun-2024 | 14.7% | 14.1% | 24.8% | 51.8% |
| 31-Jul-2024 | 14.7% | 14.1% | 24.8% | 53.4% |
| 31-Aug-2024 | 14.7% | 14.1% | 24.8% | 53.4% |

**Step 4**: Enter the assumed initial vaccine effectiveness (VE) against infection and hospitalization and the monthly linear waning rate over time. **See Section 2.2**.

In the model calculation sheets, an initial set of monthly incidence rates of symptomatic infection without seasonal vaccination is used to calculate the number of hospitalizations by month and age group. These estimated hospitalization counts are compared to the target number of hospitalizations by month and age group and the ratio of target to model estimated hospitalizations is calculated for each month and age group. These ratios are multiplied by the initial set of monthly incidence rates to determine the calibrated monthly incidence rates by age group that are required to estimate the target number of hospitalizations, given the other model inputs are held constant. These final incidence rates are then output and can be copied directly into the static CEA model. The base case incidence values are displayed in Table 5 (2023-24) and Table 6 (2024-25). The average values of 2023-24 and 2024-25, as displayed in Table 7, were used for the base case.

Table 5. 2023-24 incidence of symptomatic infection (no vaccination arm) for the static model (% infected)

| **Age group (years)** | **Sep-23** | **Oct-23** | **Nov-23** | **Dec-23** | **Jan-24** | **Feb-24** | **Mar-24** | **Apr-24** | **May-24** | **Jun-24** | **Jul-24** | **Aug-24** |
| --- | --- | --- | --- | --- | --- | --- | --- | --- | --- | --- | --- | --- |
| 12-17 | 1.0% | 0.6% | 1.0% | 1.6% | 1.7% | 1.6% | 0.8% | 0.2% | 0.4% | 0.2% | 0.8% | 1.0% |
| 18-49 | 2.0% | 1.9% | 2.3% | 3.1% | 3.4% | 2.2% | 1.1% | 0.6% | 0.6% | 1.0% | 2.0% | 2.5% |
| 50-64 | 1.4% | 1.5% | 1.9% | 2.4% | 2.7% | 1.6% | 0.9% | 0.5% | 0.4% | 0.6% | 1.3% | 1.7% |
| 65+ | 1.0% | 1.1% | 1.3% | 1.8% | 1.7% | 1.0% | 0.6% | 0.4% | 0.4% | 0.6% | 1.0% | 1.3% |

Table 6. 2024-25 incidence of symptomatic infection (no vaccination arm) for the static model (% infected)

| **Age group (years)** | **Sep-24** | **Oct-24** | **Nov-24** | **Dec-24** | **Jan-25** | **Feb-25** | **Mar-25** | **Apr-25** | **May-25** | **Jun-25** | **Jul-25** | **Aug-25** |
| --- | --- | --- | --- | --- | --- | --- | --- | --- | --- | --- | --- | --- |
| 12-17 | 0.9% | 0.6% | 0.3% | 0.5% | 0.5% | 0.5% | 0.5% | 0.3% | 0.2% | 0.1% | 0.3% | 0.6% |
| 18-49 | 1.5% | 1.0% | 0.7% | 1.2% | 1.5% | 1.1% | 0.9% | 0.6% | 0.4% | 0.4% | 0.7% | 1.4% |
| 50-64 | 1.2% | 0.8% | 0.6% | 0.9% | 1.3% | 0.9% | 0.7% | 0.4% | 0.3% | 0.2% | 0.4% | 0.7% |
| 65+ | 0.9% | 0.7% | 0.5% | 0.8% | 0.9% | 0.6% | 0.5% | 0.4% | 0.3% | 0.2% | 0.3% | 0.5% |

Table 7. Base case incidence of symptomatic infection (no vaccination arm) for the static model (% infected)

| **Age group (years)** | **Sep** | **Oct** | **Nov** | **Dec** | **Jan** | **Feb** | **Mar** | **Apr** | **May** | **Jun** | **Jul** | **Aug** |
| --- | --- | --- | --- | --- | --- | --- | --- | --- | --- | --- | --- | --- |
| 12-17 | 1.0% | 0.6% | 0.7% | 1.0% | 1.1% | 1.1% | 0.7% | 0.3% | 0.3% | 0.2% | 0.6% | 0.8% |
| 18-49 | 1.8% | 1.4% | 1.5% | 2.2% | 2.4% | 1.6% | 1.0% | 0.6% | 0.5% | 0.7% | 1.3% | 2.0% |
| 50-64 | 1.3% | 1.1% | 1.2% | 1.7% | 2.0% | 1.3% | 0.8% | 0.5% | 0.4% | 0.4% | 0.8% | 1.2% |
| 65+ | 0.9% | 0.9% | 0.9% | 1.3% | 1.3% | 0.8% | 0.6% | 0.4% | 0.3% | 0.4% | 0.6% | 0.9% |

### 2 Vaccine Effectiveness

#### 2.1 Calculation of Vaccine Effectiveness with the Model

Vaccine effectiveness declines linearly on a monthly basis within the model. A portion of the cohort can receive a new vaccine in each month of the time horizon. In order to incorporate waning of effectiveness, the effectiveness calculation is a function of the fraction of the age group vaccinated each month and vaccine effectiveness. Vaccine effectiveness is the sum of the following:

$Effectiveness={Cov}_{Cur}\times{Eff}_{Cur}+{Cov}_{Cur-1}\times{Eff}_{Cur-1}+ {Cov}_{Cur-2}\times{Eff}_{Cur-2}+\ldots+ {Cov}_{Cur-9}\times{Eff}_{Cur-9}$

Where:

Cov_cur_ = fraction of age group vaccinated in the current month

Cov_cur1_ = fraction of age group vaccinated in the month before the current month

Cov_cur2_ = fraction of the age group vaccinated two months before the current month

Eff_cur_ = effectiveness in the current month

Eff_cur1_ = effectiveness one month after initial vaccination

Eff_cur2_ = effectiveness two months after initial vaccination

#### 2.2 Inputs for the Estimation of Incidence

The 2023-2024 COVID-19 VE against infection and hospitalization for adults (aged 18 years and over) were based on a market mix of the COVID-19 mRNA vaccines mRNA-1273 and BNT162b2 administered during 2023-2024. Both VE outcomes of the Moderna XBB.1.5 vaccine were estimated based on a study by Kopel et al., (2024)^3^ that estimated the VE in those ≥18 years old receiving the vaccine between September 13, 2023 and December 15, 2023 relative to individuals that did not receive the XBB.1.5 vaccine during the same time period, independent of prior vaccination history. As the median follow-up time was 70 days post-vaccination, values for both outcomes were scaled back by 56 days to 14 days post vaccination to approximate the maximum level of protection as described in the sections below.

*Infection:*

The 2023-2024 VE for the Moderna XBB.1.5 vaccine against medically-attended SARS-CoV-2 infection at 70 days (33.1%)^3^ was used as a proxy for VE against COVID-19 symptomatic infection as this outcome was not directly measured in the Kopel et al study. As Higdon et al., (2022)^4^ approximated the aVE against infection to wane at a rate of 4.75% (95% CI 3.05%; 6.75%) monthly, the monthly waning rate of 4.75% was applied to 56 days and added to the 2023-2024 VE for the Moderna vaccine from Kopel to approximate the VE against infection at 2 weeks post-administration (41.8%). These values were used for adults aged 18 years and over.

The 2023-2024 VE for the Pfizer XBB.1.5 vaccine were estimated based on adjusted rVE values between Moderna and Pfizer vaccine values. At time of calculation, values were available for bivalent original and Omicron BA.4/5 containing mRNA-1273.222 versus BNT1162b2 , and were therefore used. rVE for outpatient visits were used as a proxy for symptomatic infection (rVE=5.1%, 95% CI 3.2-6.9%).^5^ This resulted in a 2023-2024 Pfizer XBB.1.5 VE value against infection of 38.7% in those aged 18 years and over.

The Moderna and Pfizer 2023-2024 COVID-19 VE values were then weighed by the estimated market shares of each (48% and 52%, respectively),^6^ to obtain a weighted 2023-2024 COVID-19 VE for the vaccines (40.2%).

For the age group 12 to 17 years, the 2023-2024 COVID-19 VE value was obtained from a study by Link-Gelles.^7^ The 2023-2024 COVID-19 VE for the age group of 5-17 years was 71.0% and assumed for those 12-17 years in the model. As VE was measured between 7-59 days post XBB.1.5 vaccine administration, adjustments to 2 weeks post-administration was not required.

*Hospitalization:*

The monthly waning rate of 2.46% calculated from Andersson et al., (2024)^8^ was combined with the 2023-2024 Moderna XBB.1.5 VE value against hospitalization from Kopel et al., (2024),^3^ to approximate the 2023-2024 Moderna XBB.1.5 VE against hospitalization at 14 days post-administration (64.7%).

Similar to the 2023-2024 Pfizer XBB.1.5 VE against infection, the 2023-2024 Pfizer XBB.1.5 VE against hospitalization was estimated by applying the hospitalization rVE value between the bivalent versions of mRNA-1273.222 versus BNT162b2 as a proxy (rVE=9.8%, 95% CI 2.6-16.4%).^5^ This resulted in a 2023-2024 Pfizer XBB.1.5 initial VE value against hospitalization of 60.9%.

The weighted 2023-2024 COVID-19 VE value against hospitalization using the market share values presented above is therefore 62.7%.

Table 8 summarizes the market mix VE used for calculation of the incidence of symptomatic infection without vaccination for September 2023 to August 2024.

Table 8. Market mix VE used in the calculation of infection incidence in the model (2023/24 season)

| **Age Group** | **Infection** | | **Hospitalization** | |
| --- | --- | --- | --- | --- |
|  | **Initial VE** | **Waning** | **Initial VE** | **Waning** |
| 12 to 17 years | 0.7100 | 0.0475 | 0.7100 | 0.0246 |
| 18-49 years | 0.4020 | 0.0475 | 0.6270 | 0.0246 |
| 50-64 years | 0.4020 | 0.0475 | 0.6270 | 0.0246 |
| ≥65 years | 0.4020 | 0.0475 | 0.6270 | 0.0246 |

VE: Vaccine effectiveness

The VE estimates for mRNA-1273 and BNT-162b2 for September 2024 to August 2025 (vaccines targeting KP.2). The market shares of mRNA-1273 and BNT-162b2 during this period were assumed to be the same as September 20223 to August 2024. Table 9 summarizes the market mix VE used for calculation of the incidence of symptomatic infection without vaccination for September 2024 to August 2025.

Table 9. Market mix VE used in the calculation of infection incidence in the model (2024/25 season)

| **Age Group** | **Infection** | | **Hospitalization** | |
| --- | --- | --- | --- | --- |
|  | **Initial VE** | **Waning** | **Initial VE** | **Waning** |
| 12 to 17 years | 0.624 | 0.0475 | 0.624 | 0.0246 |
| 18-49 years | 0.457 | 0.0475 | 0.544 | 0.0246 |
| 50-64 years | 0.457 | 0.0475 | 0.544 | 0.0246 |
| ≥65 years | 0.457 | 0.0475 | 0.544 | 0.0246 |

VE: Vaccine effectiveness

#### 2.3 Calculation of VE of a vaccine based on the relative vaccine effectiveness between two vaccines

When a relative vaccine effectiveness (rVE) is applied to the vaccine effectiveness (VE) value of one vaccine to estimate the VE of another, the rVE is assumed to be equivalent to the relative risk.^9^ Therefore, the VE is calculated as following: In the below calculation, the VE of mRNA-1273 is known, and the rVE of mRNA-1283 vs mRNA-1273 is applied to calculate the VE of mRNA-1283.

**Definitions**

VE_1283_ = Vaccine effectiveness of the Moderna Next Generation mRNA-1283 vaccine candidate

VE_1273_ = Vaccine effectiveness of the Moderna mRNA-1273 (Spikevax) vaccine as per Wilson et al. 2025^10^

RR _1283,1273_ =	relative risk of mRNA-1283 compared to mRNA-1273

rVE_1283,1273_ = relative vaccine effectiveness of mRNA-1283 vs mRNA-1273 as per NextCOVE Ph3 clinical trial

**Define**

Risk = 1 - VE^9^

rVE_1283,1273_ = 1 - RR _1283,1273_

**Isolate and solve for VE_1283_**

RR_1283,1273_ = Risk_1283_/Risk_1273_

RR_1283,1273_ = [1 – VE_1283_]/[1 – VE_1273_]

[1 – VE_1283_]= [1 – VE_1273_]* RR_1283,1273_

[1 – VE_1283_]= [1 – VE_1273_]*[1- rVE_1283,1273_]

VE_1283_= 1 - [1 – VE_1273_]* [1-rVE_1283,1273_]

The same approach was applied in the calibration procedure to derive the VE of BNT162b2 against infection and hospitalisation for the estimation of the market mix VE.

#### 2.4 Calculation of Incremental VE against hospitalization

The model assumes the VE against infection to be lower than the VE against hospitalization and thus applies an incremental VE against hospitalization for infections occurring in vaccinated in the vaccination arm. The hospitalization VE values are adjusted in the model to reflect the incremental protection against hospitalization above the protection against infection. In other words, VE values were adjusted to account for cases of hospitalization that are prevented due to the decrease in infections with vaccinations. This was to ensure that protection against hospitalizations is not double counted. The equations used for this are presented below.

For each age group, we define the following vaccine effectiveness variables and relationship between the variables. The superscripts and subscripts for age group are removed for clarity.

**Definitions**

${VE}_{1}$ = Vaccine effectiveness against infection

${VE}_{2}$ = ‘Total’ Vaccine effectiveness against hospitalization

${VE}_{2}^{*}$= ‘Additional’ Vaccine effectiveness against hospitalization

We assume ${VE}_{2}^{*}=0$ if there is no additional benefit against hospitalization

**Define**

$$\left[ 1-{VE}_{2} \right]= \left[ 1-{VE}_{1} \right]\times\left[ 1-{VE}_{2}^{*} \right]$$

Isolate and solve for ${VE}_{2}^{*}$

$$\left[ 1-{VE}_{2}^{*} \right]=\frac{\left[ 1-{VE}_{2} \right]}{\left[ 1-{VE}_{1} \right]}$$

$${VE}_{2}^{*}= 1- \frac{\left[ 1-{VE}_{2} \right]}{\left[ 1-{VE}_{1} \right]}$$

### 3 Cohort Size

Table 10. Cohort eligible for vaccination

| **Age Group (Years)** | **Population Size** | **Percent Eligible*** | **Eligible Population Size** | **Source** |
| --- | --- | --- | --- | --- |
| 12-17 | 26,766,952 | 25% | 6,691,738 | United Nations (2024) ^11^; Kompanieyts et al. (2021) ^12^ |
| 18-49 | 145,627,454 | 65.7% | 95,738,794 | United Nations (2024) ^11^; Panagiotakopoulos (2025) ^13^ |
| 50-64 | 63,863,350 | 81.0% | 51,729,314 |  |
| ≥65 | 59,874,349 | 100% | 59,874,349 | United Nations (2024) ^11^ |

*Reflects the proportion considered high-risk for ages 12-64 years; for ages 65+, all adults are eligible for vaccination.

*High-risk conditions for severe COVID-19 among those 12-17 years studied by Kompanieyts et al. (2021) include type 1 diabetes, cardiac and circulatory congenital anomalies, obesity, essential hypertension, epilepsy, neuropsychiatric disorders, and asthma as well as children with chronic disease. As Kompanieyts et al. (2021) studied only the prevalence of risk factors among children with a COVID-19 infection but not the prevalence among all children, we assumed a proxy estimate of 25.0% for the prevalence of these risk factors in the current analysis, based on an estimate provided in the introduction section of their study.

^†^High-risk conditions for those 18+ years reflect those included in Panagiotakopoulous et al., and include asthma, cancer (hematologic malignancies), cerebrovascular disease, chronic kidney disease (dialysis), chronic lung diseases (bronchiectasis, COPD, interstitial lung disease, pulmonary embolism, pulmonary hypertension), chronic liver diseases (cirrhosis, non-alcoholic fatty liver disease, alcoholic liver disease, autoimmune hepatitis), cystic fibrosis, diabetes mellitus (types 1 and 2), disabilities, heart conditions, HIV, mental health conditions (depression, schizophrenia spectrum disorders), neurologic conditions (dementia; Parkinson’s disease), obesity, physical inactivity, pregnancy and recent pregnancy, primary immunodeficiencies, smoking (current and former), solid organ or blood stem cell transplantation, tuberculosis, use of corticosteroids or other immunosuppressive medications

### 4 Vaccine Coverage

A figure showing the vaccine uptake by month used for the analytic time horizon, based on data from September 2024 to February 2025 from COVIDVaxView,^14,15^ is displayed below. Vaccination uptake by month for the upper and lower bound vaccination coverage rate scenarios tested in deterministic sensitivity analysis are shown in Table 11. The uptake of the second dose for the scenario where semi-annual vaccination of those 65 years and older is tested is shown in Section 1 (Figure 2).

Based on observed increases of CDC reported COVID-19 vaccination coverage rates in persons 65 years and older between season 2023/2024 and 2024/2025, two additional scenarios for 1-dose vaccination strategies were studied assuming an increase in vaccination coverage rates by 10% in the overall population, i.e. 12 to 64 years high-risk and 65 years and older, as well as 65 years and older only. The corresponding vaccination uptake is shown in Table 12.

Figure 2. First seasonal dose vaccine coverage by age used for the analytic time horizon.

Percent Coverage

|  | 12-17 years | 18-49 years | 50-64 years | 65+ years |
| --- | --- | --- | --- | --- |
| 2024-09-01 | 0% | 0% | 0% | 0% |
| 2024-09-02 | 0.141176% | 0.15% | 0.333333% | 0.683333% |
| 2024-09-03 | 0.282353% | 0.3% | 0.666667% | 1.36667% |
| 2024-09-04 | 0.423529% | 0.45% | 1% | 2.05% |
| 2024-09-05 | 0.564706% | 0.6% | 1.33333% | 2.73333% |
| 2024-09-06 | 0.705882% | 0.75% | 1.66667% | 3.41667% |
| 2024-09-07 | 0.847059% | 0.9% | 2% | 4.1% |
| 2024-09-08 | 0.988235% | 1% | 2.15714% | 4.62857% |
| 2024-09-09 | 1.12941% | 1.1% | 2.31429% | 5.15714% |
| 2024-09-10 | 1.27059% | 1.2% | 2.47143% | 5.68571% |
| 2024-09-11 | 1.41176% | 1.3% | 2.62857% | 6.21429% |
| 2024-09-12 | 1.55294% | 1.4% | 2.78571% | 6.74286% |
| 2024-09-13 | 1.69412% | 1.5% | 2.94286% | 7.27143% |
| 2024-09-14 | 1.83529% | 1.6% | 3.1% | 7.8% |
| 2024-09-15 | 1.97647% | 1.67143% | 3.38571% | 8.38571% |
| 2024-09-16 | 2.11765% | 1.74286% | 3.67143% | 8.97143% |
| 2024-09-17 | 2.25882% | 1.81429% | 3.95714% | 9.55714% |
| 2024-09-18 | 2.4% | 1.88571% | 4.24286% | 10.1429% |
| 2024-09-19 | 2.54118% | 1.95714% | 4.52857% | 10.7286% |
| 2024-09-20 | 2.68235% | 2.02857% | 4.81429% | 11.3143% |
| 2024-09-21 | 2.82353% | 2.1% | 5.1% | 11.9% |
| 2024-09-22 | 2.96471% | 2.21429% | 5.37143% | 12.3% |
| 2024-09-23 | 3.10588% | 2.32857% | 5.64286% | 12.7% |
| 2024-09-24 | 3.24706% | 2.44286% | 5.91429% | 13.1% |
| 2024-09-25 | 3.38824% | 2.55714% | 6.18571% | 13.5% |
| 2024-09-26 | 3.52941% | 2.67143% | 6.45714% | 13.9% |
| 2024-09-27 | 3.67059% | 2.78571% | 6.72857% | 14.3% |
| 2024-09-28 | 3.81176% | 2.9% | 7% | 14.7% |
| 2024-09-29 | 3.95294% | 3.11429% | 7.22857% | 15.2714% |
| 2024-09-30 | 4.09412% | 3.32857% | 7.45714% | 15.8429% |
| 2024-10-01 | 4.23529% | 3.54286% | 7.68571% | 16.4143% |
| 2024-10-02 | 4.37647% | 3.75714% | 7.91429% | 16.9857% |
| 2024-10-03 | 4.51765% | 3.97143% | 8.14286% | 17.5571% |
| 2024-10-04 | 4.65882% | 4.18571% | 8.37143% | 18.1286% |
| 2024-10-05 | 4.8% | 4.4% | 8.6% | 18.7% |
| 2024-10-06 | 4.85714% | 4.5% | 8.84286% | 19.2571% |
| 2024-10-07 | 4.91429% | 4.6% | 9.08571% | 19.8143% |
| 2024-10-08 | 4.97143% | 4.7% | 9.32857% | 20.3714% |
| 2024-10-09 | 5.02857% | 4.8% | 9.57143% | 20.9286% |
| 2024-10-10 | 5.08571% | 4.9% | 9.81429% | 21.4857% |
| 2024-10-11 | 5.14286% | 5% | 10.0571% | 22.0429% |
| 2024-10-12 | 5.2% | 5.1% | 10.3% | 22.6% |
| 2024-10-13 | 5.57143% | 5.27143% | 10.5% | 23.0857% |
| 2024-10-14 | 5.94286% | 5.44286% | 10.7% | 23.5714% |
| 2024-10-15 | 6.31429% | 5.61429% | 10.9% | 24.0571% |
| 2024-10-16 | 6.68571% | 5.78571% | 11.1% | 24.5429% |
| 2024-10-17 | 7.05714% | 5.95714% | 11.3% | 25.0286% |
| 2024-10-18 | 7.42857% | 6.12857% | 11.5% | 25.5143% |
| 2024-10-19 | 7.8% | 6.3% | 11.7% | 26% |
| 2024-10-20 | 7.82857% | 6.4% | 12.0571% | 26.2143% |
| 2024-10-21 | 7.85714% | 6.5% | 12.4143% | 26.4286% |
| 2024-10-22 | 7.88571% | 6.6% | 12.7714% | 26.6429% |
| 2024-10-23 | 7.91429% | 6.7% | 13.1286% | 26.8571% |
| 2024-10-24 | 7.94286% | 6.8% | 13.4857% | 27.0714% |
| 2024-10-25 | 7.97143% | 6.9% | 13.8429% | 27.2857% |
| 2024-10-26 | 8% | 7% | 14.2% | 27.5% |
| 2024-10-27 | 8.21429% | 7.27143% | 14.5% | 28.2429% |
| 2024-10-28 | 8.42857% | 7.54286% | 14.8% | 28.9857% |
| 2024-10-29 | 8.64286% | 7.81429% | 15.1% | 29.7286% |
| 2024-10-30 | 8.85714% | 8.08571% | 15.4% | 30.4714% |
| 2024-10-31 | 9.07143% | 8.35714% | 15.7% | 31.2143% |
| 2024-11-01 | 9.28571% | 8.62857% | 16% | 31.9571% |
| 2024-11-02 | 9.5% | 8.9% | 16.3% | 32.7% |
| 2024-11-03 | 9.62857% | 8.94286% | 16.4571% | 33.0286% |
| 2024-11-04 | 9.75714% | 8.98571% | 16.6143% | 33.3571% |
| 2024-11-05 | 9.88571% | 9.02857% | 16.7714% | 33.6857% |
| 2024-11-06 | 10.0143% | 9.07143% | 16.9286% | 34.0143% |
| 2024-11-07 | 10.1429% | 9.11429% | 17.0857% | 34.3429% |
| 2024-11-08 | 10.2714% | 9.15714% | 17.2429% | 34.6714% |
| 2024-11-09 | 10.4% | 9.2% | 17.4% | 35% |
| 2024-11-10 | 10.4% | 9.27143% | 17.5286% | 35.2% |
| 2024-11-11 | 10.4% | 9.34286% | 17.6571% | 35.4% |
| 2024-11-12 | 10.4% | 9.41429% | 17.7857% | 35.6% |
| 2024-11-13 | 10.4% | 9.48571% | 17.9143% | 35.8% |
| 2024-11-14 | 10.4% | 9.55714% | 18.0429% | 36% |
| 2024-11-15 | 10.4% | 9.62857% | 18.1714% | 36.2% |
| 2024-11-16 | 10.4% | 9.7% | 18.3% | 36.4% |
| 2024-11-17 | 10.4% | 9.82857% | 18.5143% | 36.6% |
| 2024-11-18 | 10.4% | 9.95714% | 18.7286% | 36.8% |
| 2024-11-19 | 10.4% | 10.0857% | 18.9429% | 37% |
| 2024-11-20 | 10.4% | 10.2143% | 19.1571% | 37.2% |
| 2024-11-21 | 10.4% | 10.3429% | 19.3714% | 37.4% |
| 2024-11-22 | 10.4% | 10.4714% | 19.5857% | 37.6% |
| 2024-11-23 | 10.4% | 10.6% | 19.8% | 37.8% |
| 2024-11-24 | 10.4571% | 10.6429% | 19.8714% | 37.8286% |
| 2024-11-25 | 10.5143% | 10.6857% | 19.9429% | 37.8571% |
| 2024-11-26 | 10.5714% | 10.7286% | 20.0143% | 37.8857% |
| 2024-11-27 | 10.6286% | 10.7714% | 20.0857% | 37.9143% |
| 2024-11-28 | 10.6857% | 10.8143% | 20.1571% | 37.9429% |
| 2024-11-29 | 10.7429% | 10.8571% | 20.2286% | 37.9714% |
| 2024-11-30 | 10.8% | 10.9% | 20.3% | 38% |
| 2024-12-01 | 11% | 11.0143% | 20.4286% | 38.3143% |
| 2024-12-02 | 11.2% | 11.1286% | 20.5571% | 38.6286% |
| 2024-12-03 | 11.4% | 11.2429% | 20.6857% | 38.9429% |
| 2024-12-04 | 11.6% | 11.3571% | 20.8143% | 39.2571% |
| 2024-12-05 | 11.8% | 11.4714% | 20.9429% | 39.5714% |
| 2024-12-06 | 12% | 11.5857% | 21.0714% | 39.8857% |
| 2024-12-07 | 12.2% | 11.7% | 21.2% | 40.2% |
| 2024-12-08 | 12.2% | 11.7143% | 21.3143% | 40.3286% |
| 2024-12-09 | 12.2% | 11.7286% | 21.4286% | 40.4571% |
| 2024-12-10 | 12.2% | 11.7429% | 21.5429% | 40.5857% |
| 2024-12-11 | 12.2% | 11.7571% | 21.6571% | 40.7143% |
| 2024-12-12 | 12.2% | 11.7714% | 21.7714% | 40.8429% |
| 2024-12-13 | 12.2% | 11.7857% | 21.8857% | 40.9714% |
| 2024-12-14 | 12.2% | 11.8% | 22% | 41.1% |
| 2024-12-15 | 12.2714% | 11.8429% | 22.0286% | 41.2% |
| 2024-12-16 | 12.3429% | 11.8857% | 22.0571% | 41.3% |
| 2024-12-17 | 12.4143% | 11.9286% | 22.0857% | 41.4% |
| 2024-12-18 | 12.4857% | 11.9714% | 22.1143% | 41.5% |
| 2024-12-19 | 12.5571% | 12.0143% | 22.1429% | 41.6% |
| 2024-12-20 | 12.6286% | 12.0571% | 22.1714% | 41.7% |
| 2024-12-21 | 12.7% | 12.1% | 22.2% | 41.8% |
| 2024-12-22 | 12.7429% | 12.1% | 22.2429% | 41.8429% |
| 2024-12-23 | 12.7857% | 12.1% | 22.2857% | 41.8857% |
| 2024-12-24 | 12.8286% | 12.1% | 22.3286% | 41.9286% |
| 2024-12-25 | 12.8714% | 12.1% | 22.3714% | 41.9714% |
| 2024-12-26 | 12.9143% | 12.1% | 22.4143% | 42.0143% |
| 2024-12-27 | 12.9571% | 12.1% | 22.4571% | 42.0571% |
| 2024-12-28 | 13% | 12.1% | 22.5% | 42.1% |
| 2024-12-29 | 13.1143% | 12.2286% | 22.5857% | 42.1286% |
| 2024-12-30 | 13.2286% | 12.3571% | 22.6714% | 42.1571% |
| 2024-12-31 | 13.3429% | 12.4857% | 22.7571% | 42.1857% |
| 2025-01-01 | 13.4571% | 12.6143% | 22.8429% | 42.2143% |
| 2025-01-02 | 13.5714% | 12.7429% | 22.9286% | 42.2429% |
| 2025-01-03 | 13.6857% | 12.8714% | 23.0143% | 42.2714% |
| 2025-01-04 | 13.8% | 13% | 23.1% | 42.3% |
| 2025-01-05 | 13.8143% | 13% | 23.1143% | 42.4286% |
| 2025-01-06 | 13.8286% | 13% | 23.1286% | 42.5571% |
| 2025-01-07 | 13.8429% | 13% | 23.1429% | 42.6857% |
| 2025-01-08 | 13.8571% | 13% | 23.1571% | 42.8143% |
| 2025-01-09 | 13.8714% | 13% | 23.1714% | 42.9429% |
| 2025-01-10 | 13.8857% | 13% | 23.1857% | 43.0714% |
| 2025-01-11 | 13.9% | 13% | 23.2% | 43.2% |
| 2025-01-12 | 13.9143% | 13.0286% | 23.2286% | 43.2% |
| 2025-01-13 | 13.9286% | 13.0571% | 23.2571% | 43.2% |
| 2025-01-14 | 13.9429% | 13.0857% | 23.2857% | 43.2% |
| 2025-01-15 | 13.9571% | 13.1143% | 23.3143% | 43.2% |
| 2025-01-16 | 13.9714% | 13.1429% | 23.3429% | 43.2% |
| 2025-01-17 | 13.9857% | 13.1714% | 23.3714% | 43.2% |
| 2025-01-18 | 14% | 13.2% | 23.4% | 43.2% |
| 2025-01-19 | 14.0286% | 13.2% | 23.4% | 43.2% |
| 2025-01-20 | 14.0571% | 13.2% | 23.4% | 43.2% |
| 2025-01-21 | 14.0857% | 13.2% | 23.4% | 43.2% |
| 2025-01-22 | 14.1143% | 13.2% | 23.4% | 43.2% |
| 2025-01-23 | 14.1429% | 13.2% | 23.4% | 43.2% |
| 2025-01-24 | 14.1714% | 13.2% | 23.4% | 43.2% |
| 2025-01-25 | 14.2% | 13.2% | 23.4% | 43.2% |
| 2025-01-26 | 14.2429% | 13.2714% | 23.5% | 43.3286% |
| 2025-01-27 | 14.2857% | 13.3429% | 23.6% | 43.4571% |
| 2025-01-28 | 14.3286% | 13.4143% | 23.7% | 43.5857% |
| 2025-01-29 | 14.3714% | 13.4857% | 23.8% | 43.7143% |
| 2025-01-30 | 14.4143% | 13.5571% | 23.9% | 43.8429% |
| 2025-01-31 | 14.4571% | 13.6286% | 24% | 43.9714% |
| 2025-02-01 | 14.5% | 13.7% | 24.1% | 44.1% |
| 2025-02-02 | 14.5% | 13.7571% | 24.1% | 44.1143% |
| 2025-02-03 | 14.5% | 13.8143% | 24.1% | 44.1286% |
| 2025-02-04 | 14.5% | 13.8714% | 24.1% | 44.1429% |
| 2025-02-05 | 14.5% | 13.9286% | 24.1% | 44.1571% |
| 2025-02-06 | 14.5% | 13.9857% | 24.1% | 44.1714% |
| 2025-02-07 | 14.5% | 14.0429% | 24.1% | 44.1857% |
| 2025-02-08 | 14.5% | 14.1% | 24.1% | 44.2% |
| 2025-02-09 | 14.5286% | 14.1% | 24.2% | 44.2286% |
| 2025-02-10 | 14.5571% | 14.1% | 24.3% | 44.2571% |
| 2025-02-11 | 14.5857% | 14.1% | 24.4% | 44.2857% |
| 2025-02-12 | 14.6143% | 14.1% | 24.5% | 44.3143% |
| 2025-02-13 | 14.6429% | 14.1% | 24.6% | 44.3429% |
| 2025-02-14 | 14.6714% | 14.1% | 24.7% | 44.3714% |
| 2025-02-15 | 14.7% | 14.1% | 24.8% | 44.4% |

Table 11. Vaccine coverage rates for deterministic sensitivity analyses^16-18^

|  | **Upper Bound** | | | | **Lower Bound** | | | |
| --- | --- | --- | --- | --- | --- | --- | --- | --- |
| **Age group** | **12-17 years** | **18-49 years** | **50-64 years** | **65-100 years** | **12-17 years** | **18-49 years** | **50-64 years** | **65-100 years** |
| Annual | 22.0% | 21.1% | 37.2% | 66.5% | 9.8% | 9.4% | 16.6% | 29.7% |
| Sept | 6.1% | 4.9% | 11.2% | 23.7% | 2.7% | 2.2% | 5.0% | 10.6% |
| Oct | 13.6% | 12.6% | 23.5% | 46.7% | 6.1% | 5.6% | 10.5% | 20.8% |
| Nov | 16.2% | 16.3% | 30.4% | 56.9% | 7.2% | 7.3% | 13.6% | 25.4% |
| Dec | 19.9% | 18.7% | 34.2% | 63.2% | 8.9% | 8.3% | 15.2% | 28.2% |
| Jan | 21.7% | 20.4% | 36.0% | 65.9% | 9.7% | 9.1% | 16.0% | 29.4% |
| Feb | 22.0% | 21.1% | 37.2% | 66.5% | 9.8% | 9.4% | 16.6% | 29.7% |
| March | 22.0% | 21.1% | 37.2% | 66.5% | 9.8% | 9.4% | 16.6% | 29.7% |
| April | 22.0% | 21.1% | 37.2% | 66.5% | 9.8% | 9.4% | 16.6% | 29.7% |
| May | 22.0% | 21.1% | 37.2% | 66.5% | 9.8% | 9.4% | 16.6% | 29.7% |
| June | 22.0% | 21.1% | 37.2% | 66.5% | 9.8% | 9.4% | 16.6% | 29.7% |
| July | 22.0% | 21.1% | 37.2% | 66.5% | 9.8% | 9.4% | 16.6% | 29.7% |
| August | 22.0% | 21.1% | 37.2% | 66.5% | 9.8% | 9.4% | 16.6% | 29.7% |

Table 12. Vaccine coverage rates for additional vaccine coverage rate scenarios

|  | **10% increase overall population (12 to 64 high-risk and 65 plus) (1-dose)** | | | | **10% increase 65 plus only (1-dose)** | | | |
| --- | --- | --- | --- | --- | --- | --- | --- | --- |
| **Age group** | **12-17 years** | **18-49 years** | **50-64 years** | **65-100 years** | **12-17 years** | **18-49 years** | **50-64 years** | **65-100 years** |
| Annual | 16.2% | 15.5% | 27.3% | 48.8% | 14.7% | 14.1% | 24.8% | 48.8% |
| Sept | 4.5% | 3.6% | 8.3% | 17.4% | 4.1% | 3.3% | 7.5% | 17.4% |
| Oct | 10.0% | 9.2% | 17.3% | 34.3% | 9.1% | 8.4% | 15.7% | 34.3% |
| Nov | 11.9% | 12.0% | 22.3% | 41.8% | 10.8% | 10.9% | 20.3% | 41.8% |
| Dec | 14.6% | 13.8% | 25.1% | 46.4% | 13.3% | 12.5% | 22.8% | 46.4% |
| Jan | 16.0% | 15.0% | 26.4% | 48.4% | 14.5% | 13.6% | 24.0% | 48.4% |
| Feb | 16.2% | 15.5% | 27.3% | 48.8% | 14.7% | 14.1% | 24.8% | 48.8% |
| March | 16.2% | 15.5% | 27.3% | 48.8% | 14.7% | 14.1% | 24.8% | 48.8% |
| April | 16.2% | 15.5% | 27.3% | 48.8% | 14.7% | 14.1% | 24.8% | 48.8% |
| May | 16.2% | 15.5% | 27.3% | 48.8% | 14.7% | 14.1% | 24.8% | 48.8% |
| June | 16.2% | 15.5% | 27.3% | 48.8% | 14.7% | 14.1% | 24.8% | 48.8% |
| July | 16.2% | 15.5% | 27.3% | 48.8% | 14.7% | 14.1% | 24.8% | 48.8% |
| August | 16.2% | 15.5% | 27.3% | 48.8% | 14.7% | 14.1% | 24.8% | 48.8% |

### 5 Probability of Hospitalization and Outpatient Care

#### 5.1 Derivation of Probabilities (General Population)

The model requires the probability of hospitalization in those who do not receive annual vaccination (i.e. independent of prior vaccination history) and have a symptomatic infection.

The only data that is available, comes from healthcare databases and reflects the probability of hospitalization given medically attended SARS-CoV-2 infections. It was therefore necessary to transform the data so that the denominator was all symptomatic infections and not just symptomatic, medically attended infections.

Two sources providing the number of hospitalizations among medically-attended SARS-CoV-2 infections were available. Both studies examined individuals who received the XBB.1.5 COVID-19 vaccine compared to those that did not. Data used to estimate the probability of hospitalization were based on those that did not receive an XBB.1.5 vaccine. Kopel et al., (2024)^19^ provided data on adults aged 18 years and over with COVID-19 as any diagnosis for hospitalization. The Optum’s de-identified Clinformatics® Data Mart Database^20^ also had data on any diagnosis for reason of hospitalization. A database analysis was conducted based on Optum’s de-identified Clinformatics® Data Mart Database July 2024 release (extract June 2024, with claims through May 2024). The analysis began with all patients in the Optum’s de-identified Clinformatics® Data Mart Database, and inclusion criteria were (1) at least one day of enrolment between September 12, 2023 to February 29, 2024, and (2) patients with a COVID diagnosis as the primary diagnosis between September 12, 2023 and February 29, 2024. Patients with a missing age on the index date, defined as the date of first COVID diagnosis (in primary position) were excluded. COVID-19 diagnosis was identified using ICD-10-DX codes U071 COVID-19 or J1282 Pneumonia due to COVID. To identify ICU stays, additionally the following revenue codes were applied: 0200, 0201, 0202, 0203, 0204, 0206, 0207, 0208, 0209, 0210, 0211, 0212, 0213, 0214, 0219 OR ICU_IND = 'Y'. To identify inpatient stays with invasive mechanical ventilation, HCPCS/CPT* codes E0472 and K0534 as well as ICD-10-PCS codes 5A1935Z, 5A1945Z, and 5A1955Z were applied.

Because Kopel et al.^19^ is also used for the mRNA-1273 vaccine effectiveness data, it was selected as the primary source of data for hospitalization rates, with the Optum’s de-identified Clinformatics® Data Mart Database analysis used to estimate those under the age of 18 years. A number of steps were taken in order to derive the input for static model.

**Step 1:** The number of COVID-19 related hospitalizations were divided by the number of medically-attended COVID-19 cases.

Data were available for the three older age groups for Kopel et al., (2024) (i.e. for ages 18 years and above), and for all age groups from the Optum’s de-identified Clinformatics® Data Mart Database. As Kopel et al. did not include patients under the age of 18 years, the proportion hospitalized in this age group was estimated by taking the ratio of those aged 5-17 years to those aged 18-49 years from the Optum’s de-identified Clinformatics® Data Mart Database. This ratio was applied to the proportion hospitalized in those 18-49 years from Kopel. Values from step 1 are provided in Table 13.

Table 13. Proportion hospitalized in those medically attended

| **Age group** | **Proportion hospitalized** |
| --- | --- |
| 12-17 years | 3.69% |
| 18-49 years | 2.99% |
| 50-64 years | 4.46% |
| 65+ years | 13.49% |

**Step 2:** Calculate the proportion of COVID-19 related hospitalizations in symptomatic COVID-19 cases.

In order to estimate the proportion of all infections that were hospitalized, it was next necessary to transform the proportions in Table 13 to be the proportion hospitalized amongst all symptomatic infections. Age-specific data were not available on the proportion of individuals with symptomatic COVID-19 that seek medical attention. To obtain age-specific proportions of individuals who seek medical attention for their COVID-19 symptomatic illness, the CDC reported data on the proportion of individuals, by age group, with symptomatic influenza, that sought medical attention for their illness^21^ were considered as a starting point. In order to fit a separate dynamic transmission model (DTM) to the US population, these initial values were tested and subsequently varied during the calibration procedure of the DTM to achieve a better calibration fit. Particularly, the probability of seeking care was decreased in younger age groups to achieve a better calibration model fit of the SEIR model. The final values used for the base case calibration of the DTM SEIR model are displayed in Table 14. The values calculated in Step 1 were multiplied by the proportion of symptomatic individuals seeking medical attention in each corresponding age group to obtain the probability of hospitalization per symptomatic case (Table 15).

Table 14. Proportion of those with symptomatic COVID-19 seeking medical attention

| **Age group** | **Starting Point (CDC Influenza Estimates)**^21^ | **Final Proportions for Base Case** |
| --- | --- | --- |
| 0-17 years | 52.3% | 5.0% |
| 18-49 years | 37.6% | 9.5% |
| 50-64 years | 44.1% | 25.3% |
| ≥65 years | 65.1% | 61.5% |

CDC: Centers for Disease Control and Prevention

Table 15. Hospitalization probabilities given symptomatic infection

| **Age Group** | **Probability** |
| --- | --- |
| 0-17 years | 0.18% |
| 18-49 years | 0.28% |
| 50-64 years | 1.13% |
| 65+ years | 8.29% |

The final probabilities of hospitalization given symptomatic infection and proportion who do not seek care are summarized in the Table 16. The proportion seeking outpatient care is calculated as the remainder of 1 minus the probability of hospitalization and the proportion not seeking care. For ages 12 to 64 years, these probabilities were modified to reflect the fact that those considered high-risk have a higher probability of receiving hospitalization and outpatient care compared to the general population. The methods to make these adjustments are described in the next two sections below.

Table 16. General probability of hospitalization and proportion seeking care.

| **Age Group (Years)** | **Calculated: Hospitalization Probability in Those with Symptomatic Infection** | **Proportion Not Seeking Care** | **Proportion Seeking Outpatient Care** |
| --- | --- | --- | --- |
| 12 to 17 | 0.18% | 94.99% | 4.83% |
| 18-49 | 0.28% | 90.53% | 9.19% |
| 50-64 | 1.13% | 74.68% | 24.19% |
| ≥65 | 8.29% | 38.50% | 53.20% |

#### 5.2 Derivation Of Risk Ratios for High-Risk People (Outpatient and Hospital Care)

At the ACIP April 15/16^th^ 2025 meeting, results of an analysis were presented which estimated the prevalence of US adults having at least one CDC defined medical condition putting them at high risk of severe COVID-19^22^ to be 74%^13^. Within the same meeting, data on increased risk for COVID-19 hospitalization of US adults with underlying medical conditions were presented^23^. The presented adjusted rate ratios (RR) for COVID-19–associated hospitalizations among community-dwelling adults ages 18 years and older stratified by underlying condition and age showed an increased risk of COVID-19 patients having any underlying CDC-defined condition for COVID-19 hospitalization compared to the general population. For some conditions and ages, the adjusted RR was almost 10.

To parameterize the static health economic model for high-risk patients, however, these adjusted rate ratios could not be applied as the underlying the health economic model considers a probability of COVID-19 hospitalization given symptomatic COVID-19 infection derived for the general population. Instead of using these presented rate ratios, we followed the approach of Joshi et al. 2025^24^ and applied risk ratios of increased risk of COVID-19 outpatient attendance and COVID-19 hospitalization for those having a medical condition to the underlying probabilities of COVID-19 outpatient attendance and COVID-19 hospitalization given symptomatic infection which were applied in our age-based COVID-19 vaccination model.

The risk ratios for the high-risk population having at least one CDC defined high-risk medical condition were derived as follows:

1. Derivation of hazard ratios (HR) of increased risk of COVID-19 outpatient attendance and hospitalizations derived from Moderna real-world observational electronic health records and medical claims database analyses^25^
2. Transformation of the HRs into risk ratios (RR) using an optimal minimax transformation of hazard ratio (HR) using the formula described by VanderWeele, 2020^26^

##### 5.2.1 Step 1: Derivation of Hazard Ratios

The hazard ratios for increased risk of COVID-19 related outpatient attendance and hospitalization of those having chronic conditions or high-risk condition for severe COVID-19 (i.e., chronic kidney disease, immunocompromising conditions, cardiovascular disease, chronic lung disease and diabetes) were derived in an analysis using a widely used primary care electronic medical record (EMR) platforms in the US (i.e., the Veradigm EMR dataset, which includes the Allscripts Tier 1, Allscripts Tier 2, and Practice Fusion EMR) integrated with pharmacy and medical claims data (i.e., the Komodo dataset) with an observation period of March 15^th^ to December 15^th^, 2020. N=15,127,054 individuals were included in the analysis. Of these, n=9,418,745 individuals did not have any high-risk conditions (as defined above), n=3,604,418 had exactly one of these conditions, and n=2,103,891 had two or more of these conditions.

The estimated, adjusted HRs for COVID-19 related outpatient attendance and hospitalizations of those having exactly one of these conditions versus having none of these conditions were 1.175 (95% CI: 1.672-1.734) and 1.703 (95% CI: 1.672-1.734), respectively.

Despite these estimates being derived in the early phase of the COVID-19 pandemic (wild type SARS-Cov-2 circulating), more recent Omicron related evidence (such as data presented at the ACIP April 2025 meeting^23^ and the Moderna Bench to Practice webpage^27,28^) suggests an increased risk of patients with underlying medical conditions for severe COVID-19 by means of increased hospitalization rate ratios of these patients when compared to the general population or patients without those underlying conditions. Accordingly, we consider these estimates applicable for the analysis of mRNA-1283 COVID-19 vaccination in these high-risk patients.

##### 5.2.2 Step 2: Transformation of the HRs into RRs

The resulting risk ratios were estimated to be 1.17 (95% CI: 1,15, 1.78) for COVID-19 related outpatient visits and 1.69 (95% CI: 1.66, 1.72) for COVID-19 related hospitalizations.

#### 5.3 Application of Relative Risks to Derive High-Risk Inputs

Based on the derived risk ratios for increased risk of COVID-19 outpatient attendance and hospitalization of patients having a medical condition versus persons not having a medical condition, age-specific hospitalization probabilities given symptomatic infections in not at-high risk (i.e., those not having a medical condition) and high-risk population (i.e., those having a medical condition) were estimated as follows:

We define:

- H - age-specific hospitalization probability
- P – proportion
- _total – average population
- _non - not at-high risk population
- _high - high risk population

(a) H_total = (P_high * H_high) + (P_non * H_non)

It is shown that H_high = RR * H_non, hence the equation (a) could be rewritten as:

(b) H_total = (P_high * H_non * RR) + (P_non * H_non)

(c) H_total = H_non * (P_high * RR + P_non)

(d) H_non = (H_total) / (P_high * RR + P_non)

(e) H_high = RR * H_non

### 6 Mortality and Readmission

Mortality is assumed to affect hospitalized patients only (patients receiving no formal care and outpatient care are not subject to risk of death from COVID-19 infection). The age-specific probabilities of in-hospital mortality were adjusted for the high-risk analysis using a similar approach to estimating the probability of hospitalization given symptomatic infection. Joshi et al. (2025)^24^ adapted a static Markov model for high-risk adults to estimate the clinical and economic impact of vaccination strategies; high-risk adults were defined as those having been previously diagnosed with an immunocompromising condition, chronic lung disease, chronic kidney disease, cardiovascular disease, and diabetes mellitus. Joshi et al. estimate that approximately 29.3 million US adults have diabetes, and the CDC^29^ estimates that the prevalence of diabetes ranges from 9.4% to 13.1%. Joshi et al. estimated in-hospital mortality for patients with cardiovascular disease, diabetes mellitus, and immunocompromising conditions by applying relative risks to the COVID-19 general population mortality risk, resulting in relative risks of 1.23 for diabetes, 1.62 for cardiovascular disease, and 1.74 for immunocompromising conditions. Based on Joshi et al.^24^, it was conservatively assumed that the relative risk for in-hospital mortality for high-risk patients relative to non-high-risk patients was 1.23^30^ for all locations of care (i.e., no ICU or ventilator, ICU only, or ICU with ventilator), based on patients with diabetes.

The hospital readmission rate following discharge for COVID-19 and post-discharge mortality rate were obtained from a meta-analysis by Ramzi et al.^31^ Given that most hospital readmissions and post-discharge mortality occurred within the first 30 days post-discharge, the 30-day readmission rate estimated at 8.97%% (95% CI 8.37%-11.24%%) and the 30-day post-discharge mortality rate of 7.87% (95% CI 2.78%-12,96%%) were used in model analyses.^31^ These estimates were lower but comparable to the 1 year readmission and post-discharge rates estimates specific to the USA (10.0% and 8.1%, respectively).^31^ Further, the majority of COVID-19 hospitalizations in the US during 2023-2024 had an underlying medical condition and/or involved patients ≥65 years of age.^32^ Patients with underlying medical conditions and/or older age are at higher risk for all-cause readmission following hospitalization due to respiratory infections such as seasonal influenza^33,34^ or COVID-19.^31,34^ Finally, these estimates align with US quality metrics and are comparable with Omicron-specific estimates.^34,35^

The meta-analysis did not differentiate by age or in-hospital location of care and therefore readmission rates and post-discharge mortality were assumed to be the same for all ages and for the general ward, ICU only, and ICU with mechanical ventilation.

Estimates of in-hospital mortality for the general population, stratified by age and in-hospital location of care, were obtained from the University of Michigan COVID-19 Vaccination Modeling Team and are based on COVID-NET surveillance data (March 2022 – October 2023). ^36^ Age-specific probabilities of in-hospital mortality were adjusted for the high-risk analysis using a similar approach to estimating the probability of hospitalization given symptomatic infection. Based on Joshi et al. (2025) ^24^, it was conservatively assumed that the relative risk for in-hospital mortality for high-risk patients relative to non-high-risk patients was 1.23, based on patients with diabetes.

Patients who survive the initial hospitalization are subject to risk of hospital readmission, and all patients who survive, including those who were readmitted, are subject to risk of post-discharge mortality. Unlike influenza surveillance systems, where death certificate data with pneumonia or influenza, other respiratory and circulatory causes, or other non-respiratory, non-circulatory causes of death are used to estimate deaths that occur outside the hospital (i.e., following discharge and capturing deaths related to readmission), similar surveillance systems for COVID-19 have not yet been developed. Accordingly, separate estimates for hospital readmission and post-discharge mortality related to COVID-19 are used in model analyses. The hospital readmission rate following discharge for COVID-19 and post-discharge mortality rate for the general population were obtained from a meta-analysis by Ramzi et al.^31^ Given that most hospital readmissions and post-discharge mortality occurred within the first 30 days post-discharge, the 30-day readmission (8.97%) and post-discharge mortality rates (7.87%) were used for the general population. Hospital readmission rates for high-risk patients were estimated using a similar approach as for the percentage of patients requiring hospitalization and the in-hospital mortality estimates. The RR for readmission for high-risk patients (RR=1.27) was obtained from Joshi et al. (2025) ^24^ based on the underlying risk estimate of Verna et al. (2021) ^30^ for patients with diabetes. The readmission rate for high-risk patients was calculated to be 9.49%; a weighted average of the high-risk (9.49%) and general population (8.97%) estimates of 9.35% was used for all locations of care in model analyses. Post-discharge mortality was assumed to be the same for the high-risk population as the general population.

### 7 Long COVID

Data from the CDC based on the US Census Bureau Household Pulse Survey4 were used to estimate the prevalence of long COVID for patients receiving either outpatient or hospital-based care. Estimates are based on the average percentages of adults who report currently experiencing long COVID among those who ever had COVID based on data from August 20-September 16, 2024.^37^ As data were limited to adults, children were conservatively assumed to not experience long COVID. General population estimates are applied to high-risk populations.

### 8 Adverse Events

It was assumed that patients receiving no vaccine would not experience adverse events. Grade 3 and 4 Local and Systemic adverse event (AE) rates for those ≥12 years for mRNA-1283 and mRNA-1273 were estimated from Moderna clinical trial data (NextCOVE)^38^. AE rates for BNT162b2 were assumed to be equivalent to mRNA-1273; however, no grade 4 AE for BNT162b2 was assumed. All vaccines are also associated with a risk of myocarditis/pericarditis^39^ and anaphylaxis. ^40^

Table 17. Vaccine-related adverse event rates

| **Probability** | **mRNA-1283** | **mRNA-1273** | **BNT162b2** | **Source** |
| --- | --- | --- | --- | --- |
| Grade 3 Local | 1.61% | 1.17% | 1.17% | NextCOVE trial data |
| Grade 4 Local | 0% | 0% | 0% |  |
| Grade 3 Systemic | 7.16% | 5.77% | 5.77% |  |
| Grade 4 Systemic | 0% | 0.02% | 0% |  |
| Anaphylaxis | 0.0005% | 0.0005% | 0.0005% | Klein et al. (2021) ^40^ |
| Myocarditis/Pericarditis^‡^ | 0.0008% | 0.0008% | 0.0008% | FDA Letter^39^ |

‡Myocarditis/pericarditis applies to ages 12-64 only; AE rates for BNT162b2 assumed equal to mRNA-1273; no grade 4 systemic AE was assumed for BNT162b2

### 9 Infection-Related Myocarditis

All patients with COVID-19 infection are subject to risk of infection-induced myocarditis, which is applied as a toll. The baseline rate of myocarditis in patients without COVID-19 infection and the increased risk due to COVID-19 infection was obtained from the CDC. Boehmer et al. (2021)^5^ conducted a cohort study on patients with at least one hospital-based encounter (either outpatient or inpatient) during March 2020-January 2021. The risk of myocarditis in patients with COVID-19 were compared to patients without COVID-19. The authors calculated the adjusted myocarditis risk difference, by age, between patients with and without COVID-19. These values were used to estimate the excess risk of myocarditis due to COVID-19. Where age groups reported in the CDC report did not align with age groups used in the model, a weighted average of the risks from the CDC reported age groups was used.

Table 18. Risk of myocarditis following SARS-CoV-2 infection

| Probability | Base (Range) | Source |
| --- | --- | --- |
| 12-17 years | 0.1220% | Boehmer (2021) ^41^ |
| 18-49 years | 0.0793% |  |
| 50-64 years | 0.1370% |  |
| 65+ years | 0.1800% |  |

**Assumes 50% female for all ages

### 11 Cost Inputs

Table 19. Vaccine costs

| **Vaccine Costs** | **mRNA-1283** | **BNT162b2** | **mRNA-1273** | **Source** |
| --- | --- | --- | --- | --- |
| Unit Cost | $177.12 | $147.69 | $141.80 | CDC vaccine list price^42^ |
| Administration | $20.05 | $20.05 | $20.05 | CMS 2025 Physician’s Fee Schedule^43^ |
| **Total** | **$197.17** | **$167.74** | **$161.85** |  |

CDC, Center for Disease Control; CMS, Centers for Medicare & Medicaid Services.

Table 20. Adverse event costs

| **Adverse Event** | **Cost** | **Source** |
| --- | --- | --- |
| Grade 3 Local* | $0.65 | NextCOVE clinical trial data^38,44^  CMS 2025 Physician’s Fee Schedule^43^  Drugs.com^45^  Rousculp MD, et al. 2024^46^  Walmart.com^47^ |
| Grade 3 Systemic* | $0.65 | NextCOVE clinical trial data^38,44^  CMS 2025 Physician’s Fee Schedule^43^Drugs.com^45^  Rousculp MD, et al. 2024^46^  Walmart.com^47^ |
| Grade 4 Local | $3,810 | CMS 2025 Physician’s Fee Schedule^43^  CMS Quarterly Addenda Updates^48^  Prosser et al. 2019^49^ |
| Grade 4 Systemic | $3,810 | CMS 2025 Physician’s Fee Schedule^43^  CMS Quarterly Addenda Updates^48^  Prosser et al. 2019^49^ |
| Anaphylaxis | $7,262 | CDC MMWR (2021)^50^  HCUPnet Hospital Inpatient National Statistics^51^  CMS 2025 Physician’s Fee Schedule^43^  CMS Quarterly Addenda Updates^48^  Tutle (2020)^52^ |
| Myocarditis/Pericarditis^‡^ | $9,039 | HCUPnet Hospital Inpatient National Statistics^53^  HCUPnet Average Hospital Costs per ED Visit^54^  CMS 2025 Physician’s Fee Schedule^43^ |

CDC, Center for Disease Control; CMS, Centers for Medicare and Medicaid Services; MMWR, Mortality and Morbidity weekly report.

*Based on NextCOVE clinical trial data^38,44^ on medical attendance of Grade 3 local and systemic reactogenicity events among both mRNA-1283 and mRNA-1273 (due to low numbers both vaccines were considered), it was assumed that 0.93% of grade 3 local and systemic AEs would require an outpatient visit and prescription pain medication. The cost of an outpatient visit ($54.99) was estimated from 2025 CMS data. The cost of prescription pain medications (30 oral tablets of 300 mg acetaminophen/30 mg codeine) was $11.60.^45^ Rousculp et al. (2024)^46^ estimated that 22.7% of individuals required over the counter medication use for the 6 days post COVID-19 vaccination. The cost of 6 pills of acetaminophen was estimated at $0.12.^47^

‡Applied only to those 18-49

Table 21. Hospitalization costs by location of care

| **Initial Hospitalization Costs** | **General Population Cost*** | **High-Risk Cost*** | **Final Cost**** | **Source** |
| --- | --- | --- | --- | --- |
| No ICU or Ventilator | $13,351 | $13,585 | $13,537 | Yehoshua A, et al. 2024^55^; Kapinos et al.^56^ |
| ICU only | $22,140 | $22,527 | $22,449 |  |
| ICU with Ventilator | $50,707 | $51,594 | $51,415 |  |

ICU, Intensive care unit.

* General population cost: This cost was used for the ≥65 years population; High-risk cost: General population cost multiplied by 1.07 based on a RR from Kapinos et al.^56^ used for the 12- 64 years high-risk population.

** Final costs: As the model does not allow for age-specific costs, a weighted average was calculated using the population size shown in Table 10.

Table 22. Outpatient and hospital recovery costs

| **Outpatient Costs** | **Average Number of Visits**  **(Per Patient)** | **Cost** | **Source** |
| --- | --- | --- | --- |
| Outpatient visit cost | 1.0 | $191 | Optum Database Analysis (v09062024)^57^ |
| Emergency department visit cost | 1.0 | $495 |  |
| Weighted average | | $685 |  |

The cost of outpatient care was estimated from Optum’s de-identified Clinformatics® Data Mart Database^20^ analysis, reflecting a cohort of patients with a claim for COVID-19 as the primary diagnosis between September 12, 2023 and February 29, 2024 (more detailed description of the database analysis is provided in Section 5.1). Costs of outpatient care for high-risk patients were assumed to be equal to the general population. Hospitalization cost estimates from Yehoshua et al.^55^ reflect the hospitalization length of stay only and therefore do not include any recovery or post-discharge costs; accordingly, hospitalization recovery costs were assumed to be equal to the cost of outpatient care in model analyses for all patients.

Table 23. Post-Infection costs by initial treatment location

| **Treatment location** | **Cost:**  **All patients** | **Cost:**  **High Risk Patients** | **Weighted Average Cost** | **Source** |
| --- | --- | --- | --- | --- |
| Outpatient Care | $843.60 | $2,971.87 | $2,542 | All patients:  Chambers et al. 2023^58^  High Risk patients:  Scott et al. 2024^59^ |
| Hospitalized | $1,085.98 | $16,272.25 | $13,202 |  |

For hospitalized patients, a weighted average of the costs for patients with and without ICU admission was estimated based on the percentage of high-risk versus general population patients included in the model and used in model analyses.

Table 24. Infection-related Myocarditis costs

|  | **Cost** | **Source** |
| --- | --- | --- |
| Infection-related myocarditis | $33,889 | HCUPnet Hospital Inpatient National Statistics. 2018^53^  CMS 2025 Physician’s Fee Schedule^43^ |

### 12 QALY Inputs

Table 25. Adverse event QALY losses

| **Adverse Event** | **QALY loss** | **Source** |
| --- | --- | --- |
| Grade 3 Local | 0.0004 | Walter EB, et al. (2024)^60^ |
| Grade 4 Local* | 0.0019 | Assumption based on Prosser et al. (2019)^49^ |
| Grade 3 Systemic | 0.0004 | Walter EB, et al. (2024)^60^ |
| Grade 4 Systemic* | 0.0019 | Assumption based on Prosser et al. (2019)^49^ |
| Anaphylaxis* | 0.0019 | Assumption based on Prosser et al. (2019)^49^ |
| Myocarditis/Pericarditis*^‡^ | 0.0006 | Assumption based on Prosser et al. (2019)^49^ |

LOS, length of stay; QALD, quality-adjusted life days; QALY, quality-adjusted life year.

*0.7 QALD lost due to severe AE of 3 days.

‡Median LOS due to vaccine induced myocarditis/pericarditis is 1 day (0.7 QALD/3).

Table 26. QALY lost due to infection

| **Location of Treatment** | | **QALY loss** | **Source** |
| --- | --- | --- | --- |
| **<18 years of age** | | | |
| No Formal Care | | 0.0102 | University of Michigan COVID-19 Vaccination Modeling Team^36^; Assumption (equal to outpatient care estimate) |
| Outpatient Care | | 0.0102 | Mercon et al., (2025),^61^ University of Michigan COVID-19 Vaccination Modeling Team^36^ |
| Hospitalized: | |  |  |
| No ICU or Ventilator | | 0.0320 | Mercon et al., (2025),^61^ University of Michigan COVID-19 Vaccination Modeling Team^36^ |
| ICU only | | 0.1545 |  |
| Ventilator | | 0.1545 |  |
| Readmission: | |  |  |
| No ICU or Ventilator | | 0.0320 | Assumption (equal to QALY loss of initial hospitalization) |
| ICU only | | 0.1545 |  |
| Ventilator | | 0.1545 |  |
| **18+ years of age** | | | |
| No Formal Care | | 0.0046 | University of Michigan COVID-19 Vaccination Modeling Team^36^; Assumption (equal to outpatient care estimate) |
| Outpatient Care | | 0.0046 | University of Michigan COVID-19 Vaccination Modeling Team^36^ |
| Hospitalized: | |  |  |
| No ICU or Ventilator | | 0.0174 | University of Michigan COVID-19 Vaccination Modeling Team^36^ |
| ICU only | | 0.0394 |  |
| Ventilator | | 0.0394 |  |
| Readmission: | |  |  |
| No ICU or Ventilator | | 0.0174 | Assumption (equal to QALY loss of initial hospitalization) |
| ICU only | | 0.0394 |  |
| Ventilator | | 0.0394 |  |
| **All Ages** | | | |
| No Formal Care | 0 | | Assumption |
| Outpatient Care | 0.0231 | | Sandmann et al. (2022)^62^, adjusted for outpatient care QALY loss |
| Hospitalized | 0.1026 | | PHOSP (2022)^63^ |
| Infection-related myocarditis | 0.0019 | | Assumption based on Prosser et al. (2019)^49*^ |

ICU, intensive care unit; QALY, quality-adjusted life year.

*0.7 quality-adjusted life-days (QALD) lost due to severe adverse event of 3 days.

Table 27. Baseline utility by age group

| **Age group** | **Baseline utility data for population** | **Source** |
| --- | --- | --- |
| 12-17 years | 0.9205 | Hanmer et al. (2006)^64^ |
| 18-49 years | 0.9055 |  |
| 50-64 years | 0.8662 |  |
| 65-100 years | 0.8092 |  |

### 13 Opportunity Costs

COIVD-19 hospitalizations place a high burden on the healthcare system, especially in winter.^65^ Therefore, the additional benefit of reducing COVID-19 hospitalisations in terms of reducing burden on the healthcare system was assessed by calculating the opportunity cost of bed days required for other hospitalisations using a similar approach to Brassel et al. (2023)^66^ and Sandmann et al. (2018).^67^ The opportunity cost per COVID-19 hospitalisation is the sum of the hospital cost for COVID-19 and the net monetary benefit forgone from the elective hospitalisation. The analysis assumes that hospital beds can be made available to treat other patients not hospitalised with COVID-19 infection. Other aspects of healthcare system strengthening and reducing crowded hospitals, especially during the winter months, has not been considered in the analysis.

Leuchter et al. (2025)^65^ reported that the hospital bed occupancy post COVID-19 pandemic (May 2023-April 2024) was 11% percentage points higher compared to pre-pandemic levels (2009-2019). This was primarily due to a reduction in staffed hospital beds capacity.

During the winter season, hospitals typically experience an elevated number of hospital admissions due to an increase in respiratory illnesses, leading to strains in healthcare capacity and resulting in hospitals experiencing crowded conditions. Crowded hospitals have previously been associated with adverse outcomes for patients including excess death, delays in treatment and elective procedures, increased medical errors, lower quality of care, longer inpatient stays, increased rates of healthcare-associated infections, and poorer outcomes.^68^ The CDC is monitoring respiratory illnesses impact on the healthcare system by means of tracking hospital bed occupancy as well as trying to mitigate strain by recommending vaccination for the prevention of respiratory illness associated hospitalizations in order to reduce crowding and strain in hospitals during the winter season.^69^

To recognize the impact of vaccination and prevention of COVID-19 hospitalizations on healthcare systems resilience and the reduction in the likelihood of hospitals experiencing high occupancy levels and crowded conditions, we apply the opportunity cost approach established by Sandmann et al. (2018)^67,70^ and used by others.^66,71^ In traditional cost-effectiveness analyses, costs and health burden associated with hospitalizations considered are a sum of small marginal changes/cost savings and thus can be assumed to not capture peak load opportunity cost which is a large non-marginal change in the system when hospitals are exceeding maximum capacity.^72^

A key consideration of the opportunity cost approach is that during levels of high occupancy in the hospital (i.e. hospitals experience crowding), the demand for healthcare is higher than what hospitals can offer, and due to the urgency and need to treat non-elective hospitalizations, elective hospital stays are cancelled which is associated with opportunity costs (i.e. the cost foregone for the next best option).^67^ The opportunity cost of COVID-19 hospitalizations in the model considered both economic (i.e., costs) and health (e.g., quality-adjusted life-years [QALY]) outcomes to ensure compatibility with CEA^72^ using the following formula established by Sandmann et al. (2018)^67^

$$Opportunity Costs= {Cost}_{COVID19}+\left( \frac{{LOS}_{COVID19}}{{LOS}_{Elective}} \right)x \left( {Benefit}_{Elective}*WTP-{Cost}_{Elective} \right)$$

Where:

${LOS}_{COVID19}$: Length of hospital stay of a COVID-19 hospitalization

${LOS}_{Elective}$: Length of hospital stay of an elective, non-COVID-19 treatment ${Benefit}_{Elective}$: Benefit (in QALYs) for an elective, non-COVID-19 treatment

$WTP$: Willingness to pay (assumed as per Neumann et al. at USD 100,000 per QALY)^73^

${Cost}_{COVID19}$: Average cost of COVID-19 hospitalization

${Cost}_{Elective}$: Average cost of elective care treatment

The opportunity cost is only assumed for COVID-19 hospitalizations but not for COVID-19 vaccine administration because approximately 90% of all COVID-19 vaccinations during Fall 2024 to February 22^nd^, 2025 were administered in the pharmacy setting^74^ for which opportunity cost do not apply. Additionally, it is assumed that the administration cost of vaccination is higher than the average actual cost incurred by physicians based on the time spent administering the vaccine.

The percentage of hospitalizations to include was based on the mean weekly hospital occupancy post COVID-19 pandemic (May 2023-April 2024). Using data from Leuchter et al., (2025),^65^ it was estimated that 23% of US hospitals experience on average more than 80% occupancy.^75^ It is likely a conservative assumption that 23% of COVID-19 hospitalizations occur in crowded settings (i.e. with >80% occupancy), as Leuchter reported annual mean occupancy rates and COVID-19 experiencing multiple peaks per year.

The cost and LOS of a delayed/cancelled elective inpatient stay was conservatively approximated based on non-respiratory/non-sepsis hospitalizations in the US. These were estimated using 2021 HCUPnet data, with costs inflated to 2025 USD. The length of stay of a COVID-19 hospitalization was estimated to be 8 days based on the COVID-19 Hospital data from the National Hospital Care Survey (Data from September 7, 2022 to April 12, 2023).^76^

QALYs gained from treatment of a non-infection condition was estimated to be 0.239 QALYs.^70^ This QALY estimate was based on the mean (discounted) QALYs gained from hospital treatment for non-gastroenteritis cases with chronic conditions. The estimate however considers QALY gains foregone from delayed and cancelled (i.e., no improvement of health related quality of life) hospitalisations of hospital treatments of chronic conditions but not from acute life threatening conditions^70^.

All values are summarized in Table 28.

Table 28. Opportunity cost inputs

| **Variable** | **Value** | **Source** |
| --- | --- | --- |
| Percentage of hospitalizations included | 23% | Leuchter et al. (2025)^65^ |
| Hospitalization length of stay (days) |  |  |
| COVID-19 related | 8 | National Hospital Care Survey^76^ |
| Elective | 4.65 | HCUPnet^77^ |
| Cost per hospitalization |  |  |
| COVID-19 | $0 | Assumption |
| Elective | $17,015 | HCUPnet^77^ |
| QALYs gained per treatment of a non-infection condition | 0.239 | Sandmann et al. (2018)^70^ |

HCUP: Healthcare Cost and Utilization Project; QALYs: Quality-adjusted life-year

### 14 Productivity (Patient)

Table 29. Productivity loss inputs

| **Model Parameter** | **Value** | **Data Source** |
| --- | --- | --- |
| Percentage of adults in US labor force |  |  |
| 18-49 years* | 61.5% | US Bureau of Labor Statistics^78^ |
| 50-64 years* | 55.9% | US Bureau of Labor Statistics^78^ |
| ≥65 years | 18.6% | US Bureau of Labor Statistics^78^ |
| Daily wage | $292 | US Bureau of Labor Statistics^79^ |
| Time loss (days) due to: |  |  |
| Vaccination | 0.043 | Prosser et al. (2019)^49^, CDC^74^ |
| Grade 3 Local | 0.19 | Rousculp et al. (2024)^46^ |
| Grade 3 Systemic | 0.19 | Rousculp et al. (2024)^46^ |
| Grade 4 Local** | 1.39 | Prosser et al. (2019)^49^ |
| Grade 4 Systemic** | 1.39 | Prosser et al. (2019)^49^ |
| Anaphylaxis | 2.25 | HCUPnet^51^ |
| Myocarditis/Pericarditis | 1.00 | Shimabukuro (2022)^80^ |
| Infection-related myocarditis | 3.0 | HCUPNet^53^ |
| Short-term infection period: |  |  |
| No formal care | 0.0 | Assumption |
| Outpatient care only | 6.7 | Bonner et al. (2024)^81^ |
| Hospitalization: |  |  |
| Symptoms prior to hospitalization | 6.7 | Assumption (equal to outpatient care) |
| In-hospital | 5.71 |  |
| Recovery | 8.8 | Chopra et al. (2021)^82^; Yang et al. (2024)^83^ |
| Readmission | 5.71 | Assumption (equal to original length of stay) |
| Long COVID |  |  |
| Sought outpatient care only for acute infection | 14.58 | Bartsch et al. (2025)^84^ |
| Hospitalized during acute infection | 14.58 | Bartsch et al. (2025)^84^ |

*Based on the Minnesota Department of Health Diabetes unit, 19% of patients with diabetes report being unable to work; this estimate was used to reduce the general population labor force participation rates for those at high-risk (ages 18-49 and 50-64).

**Assumed 50% of patients visit the ER and 50% require hospitalizations.

### 15 Lost Productivity (Caregiver)

Table 30. Caregiver productivity loss inputs

| **Model Parameter** | **Value** | **Data Source** |
| --- | --- | --- |
| Percentage of adults in US labor force | 75.9% | US Bureau of Labor Statistics^78^ |
| Daily wage | $292 | US Bureau of Labor Statistics^79^ |
| Time loss (days) due to: |  |  |
| Vaccination | 0.083 | Prosser et al. (2019)^49^, CDC^74^ |
| Grade 3 Local | 0.19 | Rousculp et al. (2024)^46^ |
| Grade 3 Systemic | 0.19 | Rousculp et al. (2024)^46^ |
| Grade 4 Local** | 1.39 | Prosser et al. (2019)^49^ |
| Grade 4 Systemic** | 1.39 | Prosser et al. (2019)^49^ |
| Anaphylaxis | 2.25 | HCUPnet^51^ |
| Myocarditis/Pericarditis | 1.00 | Shimabukuro (2022)^80^ |
| Infection-related myocarditis | 3.0 | HCUPNet^53^ |
| Short-term infection period: |  |  |
| No formal care | 0.80 | Assumed 50% of outpatient care |
| Outpatient care only | 1.59 | Ortega-Sanchez^85^, Optum database^20^ |
| Hospitalization: |  |  |
| Symptoms prior to hospitalization | 0 | Assumption |
| In-hospital | 7.71 | Hutton (ACIP June 2023)^86^ |
| Recovery | 0 | Assumption |
| Readmission | 7.71 | Assumption (equal to original length of stay) |
| Long COVID |  |  |
| Sought outpatient care only | 14.58 | Bartsch et al. (2025)^84^ |
| Hospitalized | 14.58 | Bartsch et al. (2025)^84^ |

**Assumed 50% of patients visit the ER and 50% require hospitalizations.

### 16 Productivity Loss due to Premature Mortality

From the societal perspective, excluding premature mortality lost productivity may result in an underestimation of the cost-effectiveness of the vaccine. A literature review of cost-effectiveness analyses of vaccines found that 23% of the 88 identified studies consider productivity losses due to pre-mature mortality.^87^ Among North American studies (18 with 16 being US specific), 33% of the studies identified considered pre-mature mortality losses. Furthermore, pre-mature mortality associated productivity losses were considered in infectious disease burden estimation in the US with authors stating that the pre-mature productivity losses may also provide a basis for cost-effectiveness analysis.^87-89^

Therefore, in the base-case analysis, the productivity losses due to COVID-19 premature mortality were included, using a human capital approach based on age-stratified market productivity loss estimates from Grosse et al. (2019).^90^ The United States Bureau of Economic Analysis Implicit Price Deflator for Gross Domestic Product was applied to adjust values from 2016 USD to 2025 USD.^91^ Non-market productivity losses were also included in scenario analyses. The adjusted values, displayed in Table 31, were linearly interpolated, to calculate values by age at one year increments.

Table 31. Market productivity loss due to premature mortality, by age

| **Model Parameter** | **Base-Case Values (Market)** | **Non-Market Values** | **Scenario (Market & Non-Market)** | **Data Source** |
| --- | --- | --- | --- | --- |
| Age |  |  |  |  |
| 0 | $1,216,853 | $695,395 | $1,912,248 | Grosse et al. (2019); US Bureau of Economic Analysis^90,91^ |
| 18 | $1,743,539 | $971,041 | $2,714,579 |  |
| 21 | $1,823,988 | $997,942 | $2,821,930 |  |
| 30 | $1,838,984 | $973,530 | $2,812,514 |  |
| 40 | $1,533,850 | $769,772 | $2,303,622 |  |
| 50 | $1,015,287 | $569,700 | $1,584,987 |  |
| 60 | $446,391 | $440,411 | $886,802 |  |
| 70 | $103,199 | $284,757 | $387,956 |  |
| 80 | $21,249 | $117,612 | $138,861 |  |
| 90 | $3,827 | $17,741 | $21,568 |  |
| 100 | $0 | $0 | $0 |  |

### 18 Details on Scenario Analyses

#### 18.1 Inputs for VE Scenario Analyses

Table 32. mRNA-1283 initial VE inputs used for scenario analyses

| **Scenario** | **12-17 years** | | **18-49 years** | | **50-64 years** | | **65+ years** | |
| --- | --- | --- | --- | --- | --- | --- | --- | --- |
|  | **Infection** | **Hospitalization** | **Infection** | **Hospitalization** | **Infection** | **Hospitalization** | **Infection** | **Hospitalization** |
| ICATT | 68.5% | 76.7% | 64.0% | 69.4% | 64.0% | 69.4% | 62.8% | 73.5% |
| rVE hospitalisation = rVE infection | 68.5% | 68.5% | 56.6% | 58.6% | 56.6% | 58.6% | 60.7% | 63.0% |

### 19 Clinical Results for Sub-Group Analyses and Comparators

Table 33. Cases Averted (mRNA-1283 relative to No Vaccine): Sub-group analysis

| **Strategy** | **Health outcomes** | | | | |
| --- | --- | --- | --- | --- | --- |
|  | **Cases** | **Outpatient** | **Long COVID** | **Hosp.** | **Deaths** |
| **Base-Case for the 12-64 High-Risk Subgroup: Annual dosing for 12-64 High-Risk** | | | | | |
| **No vaccine** | 23,447,882 | 3,244,410 | 302,181 | 135,769 | 14,923 |
| **mRNA-1283** | 22,193,971 | 3,050,836 | 283,349 | 121,702 | 13,330 |
| **Outcomes Averted by mRNA-1283 (%)** | 1,253,911 (5.3%) | 193,574 (6.0%) | 18,832 (6.2%) | 14,067 (10.4%) | 1,592 (10.7%) |
| **Base-Case for the ≥65 Subgroup: Annual dosing for ≥65 All** | | | | | |
| **No vaccine** | 5,648,149 | 3,004,934 | 275,174 | 468,490 | 62,176 |
| **mRNA-1283** | 4,685,645 | 2,536,350 | 228,747 | 345,167 | 45,809 |
| **Outcomes Averted by mRNA-1283 (%)** | 962,504 (17.0%) | 468,584 (15.6%) | 46,427 (16.9%) | 123,324 (26.3%) | 16,367 (26.3%) |

Table 34. Cases Averted (mRNA-1283 relative to mRNA-1273)

| **Strategy** | **Symptomatic Infections** | **Outpatient** | **Long COVID** | **Hospitalizations** | **Death** |
| --- | --- | --- | --- | --- | --- |
| 12-64 High-Risk, 65+ All (1-Dose) | 427,337 | 99,984 | 11,118 | 36,263 | 4,725 |
| 65+ All Subgroup  (1-Dose) | 145,660 | 57,794 | 6,886 | 31,782 | 4,218 |
| 12-64 High-Risk (1-Dose), 65+ All (2-Doses) | 427,338 | 99,985 | 11,118 | 36,263 | 4,725 |

Table 35. Cases Averted (mRNA-1283 relative to BNT162b2)

| **Strategy** | **Symptomatic Infections** | **Outpatient** | **Long COVID** | **Hospitalizations** | **Death** |
| --- | --- | --- | --- | --- | --- |
| 12-64 High-Risk, 65+ All (1-Dose) | 606,942 | 176,813 | 18,112 | 46,176 | 6,023 |
| 65+ All Subgroup  (1-Dose) | 272,265 | 126,684 | 13,070 | 40,750 | 5,408 |
| 12-64 High-Risk (1-Dose), 65+ All (2-Doses) | 607,467 | 177,136 | 18,138 | 46,176 | 6,023 |

The corresponding NNV results for the comparison between mRNA-1283 and mRNA-1273 and the comparison between mRNA-1283 and BNT162b2 are stated in Section 22 of the appendix.

### 20 Base Case Economic analysis: Sub-group Analyses

Table 36. Cost-effectiveness Analysis Results: Sub-Group Analyses (mRNA-1283 relative to No Vaccine)

| **Vaccination strategy** | **Total Costs** | **Total QALYs Lost** | **Δ Costs** | **Δ QALYs Gained*** | **ICER (Δ Cost per QALY Gained)** |
| --- | --- | --- | --- | --- | --- |
| **Base-Case for the 12-64 High-Risk Subgroup: Annual dosing for 12-64 High-Risk** | | | | | |
| No vaccine | $35,250,542,160 | 471,978 | -- | -- | Reference |
| mRNA-1283 | $38,122,218,533 | 433,447 | $2,871,676,373 | 38,531 | $74,530 |
| **Base-Case for the ≥65 Subgroup: Annual dosing for ≥65 All** | | | | | |
| mRNA-1283 | $29,237,876,546 | 413,282 | -- | -- | Reference |
| No vaccine | $29,328,941,999 | 545,901 | $91,065,453 | -132,619 | mRNA-1283 Dominates |

Abbreviations: Δ, difference; ICER, incremental cost-effectiveness ratio; QALY, quality-adjusted life-year

*Difference in QALYs gained is equivalent to negative one times the difference in total QALYs lost.

Table 37. Cost-effectiveness Analysis Results: Sub-Group Analyses (mRNA-1283 relative to mRNA-1273)

| **Vaccination strategy** | **Total Costs** | **Total QALYs Lost** | **Δ Costs** | **Δ QALYs Gained*** | **ICER (Δ Cost per QALY Gained)** |
| --- | --- | --- | --- | --- | --- |
| **Base-Case for the 12-64 High-Risk Subgroup: Annual dosing for 12-64 High-Risk** | | | | | |
| mRNA-1283 | $37,960,695,571 | 444,817 | -- | -- | Reference |
| mRNA-1273 | $38,122,218,533 | 433,447 | $161,522,962 | 11,370 | $14,206 |
| **Base-Case for the ≥65 Subgroup: Annual dosing for ≥65 All** | | | | | |
| mRNA-1283 | $29,237,876,546 | 413,282 | -- | -- | Reference |
| mRNA-1273 | $29,561,627,348 | 445,606 | $323,750,803 | -32,324 | mRNA-1283 Dominates |

Abbreviations: Δ, difference; ICER, incremental cost-effectiveness ratio; QALY, quality-adjusted life-year

*Difference in QALYs gained is equivalent to negative one times the difference in total QALYs lost.

Table 38. Cost-effectiveness Analysis Results: Sub-Group Analyses (mRNA-1283 relative to BNT162b2)

| **Vaccination strategy** | **Total Costs** | **Total QALYs Lost** | **Δ Costs** | **Δ QALYs Gained*** | **ICER (Δ Cost per QALY Gained)** |
| --- | --- | --- | --- | --- | --- |
| **Base-Case for the 12-64 High-Risk Subgroup: Annual dosing for 12-64 High-Risk** | | | | | |
| mRNA-1283 | $38,122,218,533 | 433,447 | -- | -- | Reference |
| BNT162b2 | $38,265,936,483 | 447,190 | $143,717,950 | -13,743 | mRNA-1283 Dominates |
| **Base-Case for the ≥65 Subgroup: Annual dosing for ≥65 All** | | | | | |
| mRNA-1283 | $29,237,876,546 | 413,282 | -- | -- | Reference |
| BNT162b2 | $30,332,565,858 | 456,393 | $1,094,689,312 | -43,111 | mRNA-1283 Dominates |

Abbreviations: Δ, difference; ICER, incremental cost-effectiveness ratio; QALY, quality-adjusted life-year

*Difference in QALYs gained is equivalent to negative one times the difference in total QALYs lost.

### 21 Deterministic Sensitivity Analysis Results

Figure 3. Tornado diagram for clinical outcomes (mRNA-1283 relative to no vaccination)

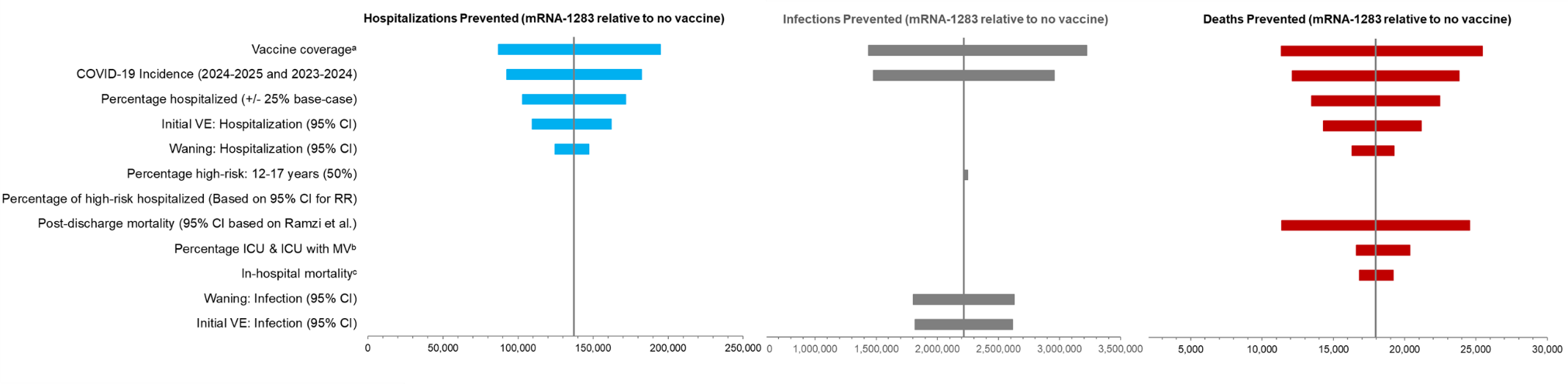

CI: confidence interval; ICU: intensive care unit; RR: relative risk; VE: vaccine efficacy.

a. Lower - narrow VCR and Higher-VCR among high-risk 2023/24. Both decrease the ICER so only the Lower bound is visible.

b. Minimum and maximum based on COVID-NET data.

c. Lower and upper bounds based on UM-CDC data.

Table 39. Clinical Results of Deterministic Sensitivity Analysis Results (mRNA-1283 relative to no vaccination)

| **Model Parameter** | **Scenario** | **Symptomatic infections averted** | | **Hospitalisations averted** | | **Deaths averted** | |
| --- | --- | --- | --- | --- | --- | --- | --- |
|  |  | **Number** | **% Change from Base** | **Number** | **% Change from Base** | **Number** | **% Change from Base** |
| Base Case | | 2,216,414 | - | 137,391 | - | 17,960 | - |
| Vaccine coverage | Upper bound based on ratio of high-risk to all adults in 2023-2024 | 3,222,065 | 45.37% | 194,752 | 41.75% | 25,436 | 41.63% |
| COVID-19 Incidence | 2023-2024 Incidence (upper bound) | 2,956,544 | 33.39% | 182,169 | 32.59% | 23,807 | 32.56% |
| Percentage hospitalized | Upper Bounds  (+25% of base-case) | 2,216,207 | -0.01% | 171,717 | 24.98% | 22,447 | 24.98% |
| Initial VE (Hospitalization) | Upper bound based on 95% CI  12-17: 86.5%  18-49: 82.2%  50-64: 82.2%  65+: 84.6% | 2,216,328 | 0.00% | 162,083 | 17.97% | 21,177 | 17.92% |
| Waning (Hospitalization) | Lower bound: 1.37% | 2,216,393 | 0.00% | 147,344 | 7.24% | 19,259 | 7.23% |
| Percentage high-risk (12-17) | 50% (Assumption) | 2,249,909 | 1.51% | 137,513 | 0.09% | 17,970 | 0.06% |
| Percentage of high-risk hospitalized | Upper bound based on 95% CI for RR (RR=1.72)  12-17: 0.27%  18-49: 0.33%  50-64: 1.23%  ≥65: 8.29% | 2,216,414 | 0.00% | 137,448 | 0.04% | 17,966 | 0.04% |
| Post-discharge mortality | Lower Bound: 2.78% (95% CI lower bound based on Ramzi et al.) | 2,216,723 | 0.01% | 137,415 | 0.02% | 11,363 | -36.73% |
| Percentage ICU & ICU with MV | Lower bounds (minimum based on COVID-NET data) | 2,216,476 | 0.00% | 137,396 | 0.00% | 16,612 | -7.50% |
| In-hospital mortality | Lower bounds  (Based on UM-CDC data) | 2,216,468 | 0.00% | 137,395 | 0.00% | 16,811 | -6.39% |
| Initial VE (infection) | Lower bound based on 95% CI  12-17: 63.1%  18-49: 49.1%  50-64: 49.1%  65+: 54.4% | 1,816,300 | -18.05% | 137,391 | 0.00% | 17,960 | 0.00% |
| Initial VE (infection) | Upper bound based on 95% CI  12-17: 73.2%  18-49: 63.0%  50-64: 63.0%  65+: 68.4% | 2,615,444 | 18.00% | 137,391 | 0.00% | 17,960 | 0.00% |
| Waning (Infection) | Lower bound: 3.05% | 2,630,285 | 18.67% | 137,391 | 0.00% | 17,960 | 0.00% |
| Waning (Infection) | Upper bound: 6.75% | 1,802,450 | -18.68% | 137,391 | 0.00% | 17,960 | 0.00% |
| In-hospital mortality | Upper bounds  (Based on UM-CDC data) | 2,216,357 | 0.00% | 137,386 | 0.00% | 19,201 | 6.91% |
| Percentage ICU & ICU with MV | Upper bounds (maximum based on COVID-NET data) | 2,216,305 | 0.00% | 137,381 | -0.01% | 20,379 | 13.47% |
| Post-discharge mortality | Upper Bound: 12.96% (95% CI upper bound based on Ramzi et al.) | 2,216,106 | -0.01% | 137,366 | -0.02% | 24,553 | 36.72% |
| Percentage of high-risk hospitalized | Lower bound based on 95% CI for RR (RR=1.66)  12-17: 0.26%  18-49: 0.33%  50-64: 1.22%  ≥65: 8.29% | 2,216,415 | 0.00% | 137,361 | -0.02% | 17,956 | -0.02% |
| Waning (hospitalization) | Upper bound: 3.87% | 2,216,442 | 0.00% | 124,598 | -9.31% | 16,290 | -9.30% |
| Initial VE (Hospitalization) | Lower bound based on 95% CI  12-17: 62.4%  18-49: 50.5%  50-64: 50.5%  65+: 57.2% | 2,216,512 | 0.00% | 109,403 | -20.37% | 14,304 | -20.35% |
| Percentage hospitalized | Lower Bounds  (-25% of base-case) | 2,216,622 | 0.01% | 103,056 | -24.99% | 13,471 | -24.99% |
| COVID-19 Incidence | 2024-2025 Incidence (lower bound) | 1,476,113 | -33.40% | 92,598 | -32.60% | 12,111 | -32.57% |
| Vaccine coverage | Lower bound based on narrower vaccine recommendations | 1,436,861 | -35.17% | 86,923 | -36.73% | 11,353 | -36.79% |

CI: confidence interval; ICU: intensive care unit; MV: mechanical ventilation; RR: relative risk; VE: vaccine efficacy.

Table 40. Economic Results of Deterministic Sensitivity Analysis Results (mRNA-1283 relative to no vaccination)

| **Model Parameter** | **Scenario** | **ICER** | **% Change from Base** |
| --- | --- | --- | --- |
| **Base Case** | | **$16,247** | **-** |
| COVID-19 Incidence | 2023-2024 Incidence (upper bound) | 1283 dominates No Vaccine | N/A |
| COVID-19 Incidence | 2024-2025 Incidence (lower bound) | $51,113 | 214.61% |
| Percentage hospitalized | Lower Bounds (-25% of base-case) | $31,826 | 95.89% |
| Post-discharge mortality | Lower Bound: 2.78% (95% CI lower bound based on Ramzi et al.) | $29,794 | 83.39% |
| Initial VE (Hospitalization) | Lower bound based on 95% CI  12-17: 62.4%  18-49: 50.5%  50-64: 50.5%  65+: 57.2% | $28,493 | 75.38% |
| Vaccine administration cost | $40 based on CMS vaccine pricing payment allowance | $23,155 | 42.52% |
| Waning (hospitalization) | Upper bound: 3.87% | $21,294 | 31.06% |
| Post-Infection Costs | Lower bound (-25% of base-case) | $21,008 | 29.31% |
| Waning (Infection) | Upper bound: 6.75% | $20,355 | 25.29% |
| Initial VE (infection) | Lower bound based on 95% CI  12-17: 63.1%  18-49: 49.1%  50-64: 49.1%  65+: 54.4% | $20,015 | 23.19% |
| Hospitalization Costs | Lower bound (-25% of base-case) | $19,906 | 22.53% |
| Percentage ICU & ICU with MV | Lower bounds (minimum based on COVID-NET data) | $18,781 | 15.60% |
| Percentage no formal care | Upper Bounds (calculated using RR=1.16 for high-risk outpatient care; ≥65 years +25% of base-case) | $18,478 | 13.73% |
| In-hospital mortality | Lower bound (Based on UM-CDC data) | $18,025 | 10.94% |
| Premature mortality productivity losses | Lower bound (-10% of base-case) | $17,539 | 7.96% |
| Percentage high-risk (12-17) | 50% (Assumption) | $17,239 | 6.11% |
| Post-infection QALY loss | Lower bounds (outpatient based on 95% CI; hospitalization -25% of base-case value) | $17,087 | 5.17% |
| Acute Phase QALYs Lost | Lower bound based on low bounds of ranges from UM-CDC and Mercon (2025) | $17,043 | 4.90% |
| No Formal Care QALY Loss | Assume 50% of outpatient values | $16,571 | 2.00% |
| Long COVID Time Loss | Lower bound of 8.11 days from Bartsch | $16,462 | 1.32% |
| Outpatient Care Cost | Lower Bound (Equal to UM-CDC values) of $476.92 | $16,404 | 0.97% |
| Percentage with Long COVID | Lower bounds (95% CI lower bounds) | $16,370 | 0.76% |
| Hospital readmission | Assume same as general population: 8.97% | $16,299 | 0.32% |
| Percentage of high-risk hospitalized | Lower bound based on 95% CI for RR (RR=1.66)  12-17: 0.26%  18-49: 0.33%  50-64: 1.22%  ≥65: 8.29% | $16,271 | 0.15% |
| Caregiver QALY Loss (<18) | Excluded from acute phase QALY loss estimates for those <18 years | $16,261 | 0.09% |
| Hospital readmission | Assume 9.66% for high risk patients (9.47% overall) | $16,230 | -0.10% |
| Percentage of high-risk hospitalized | Upper bound based on 95% CI for RR (RR=1.72)  12-17: 0.27%  18-49: 0.33%  50-64: 1.23%  ≥65: 8.29.% | $16,198 | -0.30% |
| Percentage with Long COVID | Upper bounds (95% CI upper bounds) | $16,092 | -0.95% |
| Long COVID Time Loss | Upper bound of 21.05 days from Bartsch | $16,031 | -1.32% |
| Hospital Recovery Time Loss | 19.3 days based on Chopra et al. | $15,720 | -3.24% |
| Acute Phase QALYs Lost | Upper bound based on upper bounds of ranges from UM-CDC and Mercon (2025) | $15,516 | -4.50% |
| Post-infection QALY loss | Lower bounds (outpatient based on 95% CI; hospitalization +25% of base-case value) | $15,438 | -4.98% |
| Premature mortality productivity losses | Upper bound (+10% of base-case) | $14,954 | -7.96% |
| In-hospital mortality | Upper bounds (Based on UM-CDC data) | $14,646 | -9.85% |
| Percentage no formal care | Lower Bounds (calculated using RR=1.78 for high-risk outpatient care; ≥65 years -25% of base-case) | $13,615 | -16.20% |
| Vaccine coverage | Lower bound based on UK data following narrowing of vaccine recommendations | $12,912 | -20.52% |
| Vaccine coverage | Upper bound based on ratio of high-risk to all adults in 2023-2024 | $12,908 | -20.55% |
| Waning (hospitalization) | Lower bound: 1.37% | $12,841 | -20.96% |
| Percentage ICU & ICU with MV | Upper bounds (maximum based on COVID-NET data) | $12,787 | -21.30% |
| Hospitalization Costs | Upper bound (+25% of base-case) | $12,587 | -22.53% |
| Waning (Infection) | Lower bound: 3.05% | $12,468 | -23.26% |
| Initial VE (infection) | Upper bound based on 95% CI  12-17: 73.2%  18-49: 63.0%  50-64: 63.0%  65+: 68.4% | $12,387 | -23.76% |
| Post-Infection Costs | Upper bound (+25% of base-case) | $11,485 | -29.31% |
| Post-discharge mortality | Upper Bound: 12.96% (95% CI upper bound based on Ramzi et al.) | $8,784 | -45.93% |
| Initial VE (Hospitalization) | Upper bound based on 95% CI  12-17: 86.5%  18-49: 82.2%  50-64: 82.2%  65+: 84.6% | $8,301 | -48.91% |
| Percentage hospitalized | Upper Bounds (+25% of base-case) | $6,100 | -62.45% |

CI: confidence interval; ICU: intensive care unit; MV: mechanical ventilation; QALY: quality-adjusted life year; RR: relative risk; VE: vaccine efficacy.

### 22 Scenario Analysis Results

#### 22.1 mRNA-1283 relative to no vaccination

Table 41. Clinical Scenario Analysis Results (mRNA-1283 relative to no vaccination)

| **Model Parameter** | **Range/Data Selections** | **Symptomatic Infections Averted** | | **Hospitalizations Averted** | | **Deaths Averted** | |
| --- | --- | --- | --- | --- | --- | --- | --- |
|  |  | **Number** | **% Change*** | **Number** | **% Change*** | **Number** | **% Change*** |
| Base-Case | | 2,216,414 | - | 137,391 | - | 17,960 | - |
| Vaccine Efficacy | rVE against hospitalization = rVE against infection | 2,216,495 | 0.00% | 114,359 | -16.76% | 14,953 | -16.74% |
| Vaccine coverage rates | Increase 10% (Add to September; across all ages) | 3,223,501 | 45.44% | 174,189 | 26.78% | 22,665 | 26.20% |
| Vaccine coverage rates | Increase 5% (Add to September; across all ages) | 2,719,957 | 22.72% | 155,790 | 13.39% | 20,312 | 13.10% |
| Percentage hospitalized (high-risk) | Increased RR of hospitalization (RR=1.45) for high-risk | 2,216,417 | 0.00% | 137,071 | -0.23% | 17,924 | -0.20% |
| Inpatient mortality (high-risk) | Increased mortality (RR=1.62) for high-risk | 2,216,417 | 0.00% | 137,391 | 0.00% | 17,915 | -0.25% |
| Vaccine coverage rates | Increase coverage by 5% in September for ages 65+ only | 2,324,324 | 4.87% | 151,875 | 10.54% | 19,882 | 10.70% |
| Vaccine coverage rates | Increase coverage by 10% in September for ages 65+ only | 2,432,234 | 9.74% | 166,358 | 21.08% | 21,804 | 21.41% |
| Hospital readmission | Assume same as general population: 8.97% | 2,216,414 | 0.00% | 137,391 | 0.00% | 17,960 | 0.00% |
| Hospital readmission | Assume 9.66% for high risk patients (9.47% overall) | 2,216,414 | 0.00% | 137,391 | 0.00% | 17,960 | 0.00% |
| Indirect benefit | Based on CDC Right Study | 2,937,958 | 32.55% | 137,391 | 0.00% | 17,960 | 0.00% |

RR: relative risk; rVE: relative vaccine effectiveness

*From Base-Case Results

#### 22.2 mRNA-1283 relative to mRNA-1273

Table 42. Clinical Scenario Analysis Results (mRNA-1283 relative to mRNA-1273)

| **Model Parameter** | **Range/Data Selections** | **Symptomatic Infections Averted** | | **Hospitalizations Averted** | | **Deaths Averted** | |
| --- | --- | --- | --- | --- | --- | --- | --- |
|  |  | **Number** | **% Change*** | **Number** | **% Change*** | **Number** | **% Change*** |
| Strategy: 12-64 High-Risk, 65+ All (1-Dose) | | | | | | | |
| Base-Case | | 427,337 | - | 36,263 | - | 4,725 | - |
| COVID-19 Incidence | 2023-2024 Incidence (upper bound) | 573,109 | 34.1% | 48,281 | 33.1% | 6,290 | 33.1% |
| COVID-19 Incidence | 2024-2025 Incidence (lower bound) | 281,530 | -34.1% | 24,243 | -33.1% | 3,160 | -33.1% |
| Initial VE (infection) | Based on lower 95% CI of rVE  12-17: 73.2%  18-49: 63.0%  50-64: 63.0%  65+: 68.4% | 826,367 | 93.4% | 36,263 | 0.0% | 4,725 | 0.0% |
| Initial VE (infection) | Based on upper 95% CI of rVE  12-17: 63.1%  18-49: 49.1%  50-64: 49.1%  65+: 54.4% | 27,223 | -93.6% | 36,263 | 0.0% | 4,725 | 0.0% |
| Initial VE (Hospitalization) | Based on lower 95% CI of rVE  12-17: 86.5%  18-49: 82.2%  50-64: 82.2%  65+: 84.6% | 427,251 | 0.0% | 60,956 | 68.1% | 7,943 | 68.1% |
| Initial VE (Hospitalization) | Based on upper 95% CI of rVE  12-17: 62.4%  18-49: 50.5%  50-64: 50.5%  65+: 57.2% | 427,435 | 0.0% | 8,275 | -77.2% | 1,070 | -77.4% |
| Vaccine coverage | Upper bound based on ratio of high-risk to all adults in 2023-2024 | 627,965 | 46.9% | 51,670 | 42.5% | 6,727 | 42.4% |
| Vaccine coverage | Lower bound based on narrower vaccine recommendations | 280,011 | -34.5% | 23,061 | -36.4% | 3,002 | -36.5% |
| Strategy: 65+ All Subgroup (1-Dose) | | | | | | | |
| Base-Case | | 145,660 | - | 31,782 | - | 4,218 | - |
| COVID-19 Incidence | 2023-2024 Incidence (upper bound) | 193,506 | 32.8% | 42,229 | 32.9% | 5,604 | 32.9% |
| COVID-19 Incidence | 2024-2025 Incidence (lower bound) | 97,784 | -32.9% | 21,332 | -32.9% | 2,831 | -32.9% |
| Initial VE (infection) | Based on lower 95% CI of rVE  12-17: 73.2%  18-49: 63.0%  50-64: 63.0%  65+: 68.4% | 326,704 | 124.3% | 31,782 | 0.0% | 4,218 | 0.0% |
| Initial VE (infection) | Based on upper 95% CI of rVE  12-17: 63.1%  18-49: 49.1%  50-64: 49.1%  65+: 54.4% | -2,468 | -101.7% | 31,782 | 0.0% | 4,218 | 0.0% |
| Initial VE (Hospitalization) | Based on lower 95% CI of rVE  12-17: 86.5%  18-49: 82.2%  50-64: 82.2%  65+: 84.6% | 145,587 | -0.1% | 53,427 | 68.1% | 7,091 | 68.1% |
| Initial VE (Hospitalization) | Based on upper 95% CI of rVE  12-17: 62.4%  18-49: 50.5%  50-64: 50.5%  65+: 57.2% | 145,744 | 0.1% | 6,824 | -78.5% | 906 | -78.5% |
| Vaccine coverage | Upper bound based on ratio of high-risk to all adults in 2023-2024 | 206,024 | 41.4% | 44,954 | 41.4% | 5,966 | 41.4% |
| Vaccine coverage | Lower bound based on narrower vaccine recommendations | 91,967 | -36.9% | 20,067 | -36.9% | 2,663 | -36.9% |

CI: Confidence interval; VE: Vaccine efficacy.

Table 43. Economic Scenario Analysis Results (mRNA-1283 relative to mRNA-1273)

| **Model Parameter** | **Range/Data Selections** | **ICER** | **% Change*** |
| --- | --- | --- | --- |
| Strategy: 12-64 High-Risk, 65+ All (1-Dose) | | | |
| Base-Case | | 1283 dominates 1273 | - |
| COVID-19 Incidence | 2023-2024 Incidence (upper bound) | 1283 dominates 1273 | N/A |
| COVID-19 Incidence | 2024-2025 Incidence (lower bound) | $20,397 | N/A |
| Initial VE (infection) | Based on lower 95% CI of rVE  12-17: 73.2%  18-49: 63.0%  50-64: 63.0%  65+: 68.4% | 1283 dominates 1273 | N/A |
| Initial VE (infection) | Based on upper 95% CI of rVE  12-17: 63.1%  18-49: 49.1%  50-64: 49.1%  65+: 54.4% | $9,907 | N/A |
| Initial VE (Hospitalization) | Based on lower 95% CI of rVE  12-17: 86.5%  18-49: 82.2%  50-64: 82.2%  65+: 84.6% | 1283 dominates 1273 | N/A |
| Initial VE (Hospitalization) | Based on upper 95% CI of rVE  12-17: 62.4%  18-49: 50.5%  50-64: 50.5%  65+: 57.2% | $77,473 | N/A |
| Vaccine coverage | Upper bound based on ratio of high-risk to all adults in 2023-2024 | 1283 dominates 1273 | N/A |
| Vaccine coverage | Lower bound based on narrower vaccine recommendations | 1283 dominates 1273 | N/A |
| Strategy: 65+ All Subgroup (1-Dose) | | | |
| Base-Case | | 1283 dominates 1273 | - |
| COVID-19 Incidence | 2023-2024 Incidence (upper bound) | 1283 dominates 1273 | N/A |
| COVID-19 Incidence | 2024-2025 Incidence (lower bound) | $6,891 | N/A |
| Initial VE (infection) | Based on lower 95% CI of rVE  12-17: 73.2%  18-49: 63.0%  50-64: 63.0%  65+: 68.4% | 1283 dominates 1273 | N/A |
| Initial VE (infection) | Based on upper 95% CI of rVE  12-17: 63.1%  18-49: 49.1%  50-64: 49.1%  65+: 54.4% | $693 | N/A |
| Initial VE (Hospitalization) | Based on lower 95% CI of rVE  12-17: 86.5%  18-49: 82.2%  50-64: 82.2%  65+: 84.6% | 1283 dominates 1273 | N/A |
| Initial VE (Hospitalization) | Based on upper 95% CI of rVE  12-17: 62.4%  18-49: 50.5%  50-64: 50.5%  65+: 57.2% | $60,605 | N/A |
| Vaccine coverage | Upper bound based on ratio of high-risk to all adults in 2023-2024 | 1283 dominates 1273 | N/A |
| Vaccine coverage | Lower bound based on narrower vaccine recommendations | 1283 dominates 1273 | N/A |

CI: Confidence interval; N/A: Not Applicable; VE: Vaccine efficacy.

#### 22.3 mRNA-1283 relative to BNT162b2

Table 44. Clinical Scenario Analysis Results (mRNA-1283 relative to BNT162b2)

| **Model Parameter** | **Range/Data Selections** | **Symptomatic Infections Averted** | | **Hospitalizations Averted** | | **Deaths Averted** | |
| --- | --- | --- | --- | --- | --- | --- | --- |
|  |  | **Number** | **% Change*** | **Number** | **% Change*** | **Number** | **% Change*** |
| Strategy: 12-64 High-Risk, 65+ All (1-Dose) | | | | | | | |
| Base-Case | | 606,942 | - | 46,176 | - | 6,023 | - |
| COVID-19 Incidence | 2023-2024 Incidence (upper bound) | 812,347 | 33.8% | 61,473 | 33.1% | 8,016 | 33.1% |
| COVID-19 Incidence | 2024-2025 Incidence (lower bound) | 401,489 | -33.9% | 30,876 | -33.1% | 4,029 | -33.1% |
| Initial VE (infection) | Based on lower 95% CI of rVE  12-17: 62.4%  18-49: 54.4%  50-64: 54.4%  65+: 59.2% | 113,438 | -81.3% | 31,531 | -31.7% | 4,115 | -31.7% |
| Initial VE (infection) | Based on upper 95% CI of rVE  12-17: 62.4%  18-49: 37.2%  50-64: 37.2%  65+: 36.5% | 1,134,436 | 86.9% | 46,483 | 0.7% | 6,058 | 0.6% |
| Initial VE (Hospitalization) | Based on lower 95% CI of rVE  12-17: 62.4%  18-49: 68.3%  50-64: 68.3%  65+: 72.6% | 607,122 | 0.0% | 1,954 | -95.8% | 253 | -95.8% |
| Initial VE (Hospitalization) | Based on upper 95% CI of rVE  12-17: 62.4%  18-49: 5.1%  50-64: 5.1%  65+: 17.9% | 606,915 | 0.0% | 53,194 | 15.2% | 6,954 | 15.5% |
| Vaccine coverage | Upper bound based on ratio of high-risk to all adults in 2023-2024 | 886,211 | 46.0% | 65,771 | 42.4% | 8,571 | 42.3% |
| Vaccine coverage | Lower bound based on narrower vaccine recommendations | 395,227 | -34.9% | 29,354 | -36.4% | 3,825 | -36.5% |
| Strategy: 65+ All Subgroup (1-Dose) | | | | | | | |
| Base-Case | | 272,265 | - | 40,750 | - | 5,408 | - |
| COVID-19 Incidence | 2023-2024 Incidence (upper bound) | 361,426 | 32.7% | 54,145 | 32.9% | 7,186 | 32.9% |
| COVID-19 Incidence | 2024-2025 Incidence (lower bound) | 183,062 | -32.8% | 27,351 | -32.9% | 3,630 | -32.9% |
| Initial VE (infection) | Based on lower 95% CI of rVE  12-17: 62.4%  18-49: 54.4%  50-64: 54.4%  65+: 59.2% | 36,302 | -86.7% | 27,976 | -31.3% | 3,713 | -31.3% |
| Initial VE (infection) | Based on upper 95% CI of rVE  12-17: 62.4%  18-49: 37.2%  50-64: 37.2%  65+: 36.5% | 531,094 | 95.1% | 40,750 | 0.0% | 5,408 | 0.0% |
| Initial VE (Hospitalization) | Based on lower 95% CI of rVE  12-17: 62.4%  18-49: 68.3%  50-64: 68.3%  65+: 72.6% | 272,418 | 0.1% | 1,686 | -95.9% | 224 | -95.9% |
| Initial VE (Hospitalization) | Based on upper 95% CI of rVE  12-17: 62.4%  18-49: 5.1%  50-64: 5.1%  65+: 17.9% | 272,237 | 0.0% | 47,768 | 17.2% | 6,340 | 17.2% |
| Vaccine coverage | Upper bound based on ratio of high-risk to all adults in 2023-2024 | 384,873 | 41.4% | 57,639 | 41.4% | 7,650 | 41.4% |
| Vaccine coverage | Lower bound based on narrower vaccine recommendations | 171,800 | -36.9% | 25,730 | -36.9% | 3,415 | -36.9% |

CI: Confidence interval; VE: Vaccine efficacy.

Table 45. Economic Scenario Analysis Results (mRNA-1283 relative to BNT162b2)

| **Model Parameter** | **Range/Data Selections** | **ICER** | **% Change*** |
| --- | --- | --- | --- |
| Strategy: 12-64 High-Risk, 65+ All (1-Dose) | | | |
| Base-Case | | 1283 dominates BNT162b2 | - |
| COVID-19 Incidence | 2023-2024 Incidence (upper bound) | 1283 dominates BNT162b2 | N/A |
| COVID-19 Incidence | 2024-2025 Incidence (lower bound) | 1283 dominates BNT162b2 | N/A |
| Initial VE (infection) | Based on lower 95% CI of rVE  12-17: 62.4%  18-49: 54.4%  50-64: 54.4%  65+: 59.2% | $5,793 | N/A |
| Initial VE (infection) | Based on upper 95% CI of rVE  12-17: 62.4%  18-49: 37.2%  50-64: 37.2%  65+: 36.5% | 1283 dominates BNT162b2 | N/A |
| Initial VE (Hospitalization) | Based on lower 95% CI of rVE  12-17: 62.4%  18-49: 68.3%  50-64: 68.3%  65+: 72.6% | $80,229 | N/A |
| Initial VE (Hospitalization) | Based on upper 95% CI of rVE  12-17: 62.4%  18-49: 5.1%  50-64: 5.1%  65+: 17.9% | 1283 dominates BNT162b2 | N/A |
| Vaccine coverage | Upper bound based on ratio of high-risk to all adults in 2023-2024 | 1283 dominates BNT162b2 | N/A |
| Vaccine coverage | Lower bound based on narrower vaccine recommendations | 1283 dominates BNT162b2 | N/A |
| Strategy: 65+ All Subgroup (1-Dose) | | | |
| Base-Case | | 1283 dominates BNT162b2 | - |
| COVID-19 Incidence | 2023-2024 Incidence (upper bound) | 1283 dominates BNT162b2 | N/A |
| COVID-19 Incidence | 2024-2025 Incidence (lower bound) | 1283 dominates BNT162b2 | N/A |
| Initial VE (infection) | Based on lower 95% CI of rVE  12-17: 62.4%  18-49: 54.4%  50-64: 54.4%  65+: 59.2% | 1283 dominates BNT162b2 | N/A |
| Initial VE (infection) | Based on upper 95% CI of rVE  12-17: 62.4%  18-49: 37.2%  50-64: 37.2%  65+: 36.5% | 1283 dominates BNT162b2 | N/A |
| Initial VE (Hospitalization) | Based on lower 95% CI of rVE  12-17: 62.4%  18-49: 68.3%  50-64: 68.3%  65+: 72.6% | $39,733 | N/A |
| Initial VE (Hospitalization) | Based on upper 95% CI of rVE  12-17: 62.4%  18-49: 5.1%  50-64: 5.1%  65+: 17.9% | 1283 dominates BNT162b2 | N/A |
| Vaccine coverage | Upper bound based on ratio of high-risk to all adults in 2023-2024 | 1283 dominates BNT162b2 | N/A |
| Vaccine coverage | Lower bound based on narrower vaccine recommendations | 1283 dominates BNT162b2 | N/A |

CI: Confidence interval; N/A: Not Applicable; VE: Vaccine efficacy.

### 23 Antimicrobial Resistance

Antimicrobial resistance (AMR) is a global health threat where microorganisms, such as bacteria, viruses, fungi, and parasites, become resistant to the drugs designed to treat them. Antibiotics can save lives, but any time they are used they can contribute to resistance.

Antibiotic prescriptions received in the outpatient setting for COVID-19 patients has been reported in multiple studies.^92,93^ Table 46 displays the proportion of COVID-19 patients in the outpatient setting estimated to receive antibiotic prescriptions.

Table 46. Proportion of COVID-19 outpatients receiving antibiotic prescriptions

| **Age group** | **Proportion prescribed antibiotics** | **Source** |
| --- | --- | --- |
| 12-17 years | 5.40% | Extrapolated from Figure 2 in Wittman et al. 2023^92^ |
| 18-49 years | 11.51% | Extrapolated from Figure 2 in Wittman et al. 2023^92^ for age groups 18-24, 25-44, and 45-64, and then weighted by US population size. |
| 50-64 years | 16.00% | Extrapolated from Figure 2 in Wittman et al. 2023^92^ |
| 65+ years | 29.90% | Based on data from Tsay et al. 2022^93^. Weighted average based on US population size for age groups. |

The number of antibiotic prescriptions in the outpatient setting that could be avoided with use of mRNA-1283 compared to no vaccine is shown in Table 47.

Table 47. Number of antibiotic prescriptions in the outpatient setting avoided

| **Age groups** | **Number of Prescriptions** | | **Prescriptions avoided** |
| --- | --- | --- | --- |
|  | **No Vaccine** | **1283** |  |
| 12-17 years | 1,608 | 1,512 | 96 |
| 18-49 years | 181,721 | 174,089 | 7,632 |
| 50-64 years | 261,821 | 241,740 | 20,081 |
| 65-100 years | 898,344 | 758,258 | 140,086 |
| Total | 1,343,494 | 1,175,599 | 167,896 |

### 24 Number Needed to Vaccinate

Table 48. Number needed to vaccinate (mRNA-1283 relative to no vaccination)

| Strategy | Symptomatic Infections | Outpatient | Long COVID | Hospitalizations | Deaths |
| --- | --- | --- | --- | --- | --- |
| 12-64 High-Risk, 65+ All (1-Dose) | 27 | 90 | 908 | 432 | 3,301 |
| 65+ All Subgroup (1-Dose) | 33 | 68 | 689 | 259 | 1,953 |

Table 49. Second Dose: Number needed to vaccinate (mRNA-1283 relative to no vaccination)

| Strategy | Symptomatic Infections | Outpatient | Long COVID | Hospitalizations | Deaths |
| --- | --- | --- | --- | --- | --- |
| 12-64 High-Risk, 65+ All (1-Dose): Cases averted | 2,216,414 | 662,158 | 65,259 | 137,391 | 17,960 |
| 12-64 High-Risk, 65+ All (2-Dose): Cases averted | 2,254,353 | 683,857 | 67,124 | 139,023 | 18,176 |
| Additional cases avereted with 2nd dose | 37,939 | 21,699 | 1,865 | 1,632 | 217 |
| Number of 2nd dose vaccinations | 5,388,691 | | | | |
| NNV with second dose* | 142 | 248 | 2,890 | 3,301 | 24,874 |

*To avert one additional outcome compared to 1-dose strategy.

Table 50. Difference in Number Needed to Vaccinate Between mRNA-1283 and mRNA-1273 Compared to No Vaccine*

| Strategy/Comparison | Symptomatic Infections | Outpatient | Long COVID | Hospitalizations | Deaths |
| --- | --- | --- | --- | --- | --- |
| Base-Case: 12-64 High-Risk, 65+ All (1-Dose) | | | | | |
| mRNA-1283 vs. No vaccine | 27 | 90 | 908 | 432 | 3,301 |
| mRNA-1273 vs. No vaccine | 33 | 105 | 1,095 | 586 | 4,480 |
| mRNA-1283 vs. mRNA-1273 | 6 | 16 | 187 | 155 | 1,179 |
| Base-Case: 65+ All Subgroup (1-Dose) | | | | | |
| mRNA-1283 vs. No vaccine | 33 | 68 | 689 | 259 | 1,953 |
| mRNA-1273 vs. No vaccine | 39 | 78 | 809 | 349 | 2,632 |
| **mRNA-1283 vs. mRNA-1273** | 6 | 10 | 120 | 90 | 678 |

* Calculated as (NNV mRNA-1273 compared to No Vaccine) - (NNV mRNA-1283 compared to No Vaccine).

Table 51. Difference in Number Needed to Vaccinate Between mRNA-1283 and BNT162b2 Compared to No Vaccine*

| Strategy/Comparison | Symptomatic Infections | Outpatient | Long COVID | Hospitalizations | Deaths |
| --- | --- | --- | --- | --- | --- |
| Base-Case: 12-64 High-Risk, 65+ All (1-Dose) | | | | | |
| mRNA-1283 vs. No vaccine | 27 | 90 | 908 | 432 | 3,301 |
| BNT162b2 vs. No vaccine | 37 | 122 | 1,257 | 650 | 4,967 |
| mRNA-1283 vs. BNT162b2 | 10 | 33 | 349 | 218 | 1,666 |
| Base-Case: 65+ All Subgroup (1-Dose) | | | | | |
| mRNA-1283 vs. No vaccine | 33 | 68 | 689 | 259 | 1,953 |
| BNT162b2 vs. No vaccine | 46 | 94 | 959 | 387 | 2,918 |
| **mRNA-1283 vs. BNT162b2** | 13 | 25 | 270 | 128 | 964 |

* Calculated as (NNV BNT162b2 compared to No Vaccine) - (NNV mRNA-1283 compared to No Vaccine).

When considering additional symptomatic infections, outpatient episodes, long COVID cases, hospitalizations and death prevented by mRNA-1283 over mRNA-1273, the corresponding incremental NNVs were estimated at 6, 16, 187, 155, and 1,179 [Base-Case: 12-64 High-Risk, 65+ All (1-Dose)]. For hospitalisations this means that 155 more persons would need to be vaccinated with mRNA-1273 to prevent a hospitalization when compared with mRNA-1283.

Again, when considering additional symptomatic infections, outpatient episodes, long COVID cases, hospitalizations and death prevented by mRNA-1283 over BNT162b2, the corresponding incremental NNVs were estimated at 10, 33, 349, 218, and 1,666 [Base-Case: 12-64 High-Risk, 65+ All (1-Dose)]. For hospitalisations this means that 218 more persons would need to be vaccinated with BNT162b2 to prevent a hospitalization when compared with mRNA-1283.

### 25 Cost-Benefit Ratio

Table 52. Cost-Benefit Ratio Societal Perspective (mRNA-1283 relative to no vaccination)

| **Strategy** | **QALYs Monetized (Value=$100k)** | **QALYs**  **Monetized (Value=$150k)** | **LYs**  **Monetized (VSLY $604k)** | **QALYs**  **Monetized (VQALY $717k)** |
| --- | --- | --- | --- | --- |
| 12-64 High-Risk, 65+ All (1-Dose) | 2.16 | 2.84 | 8.91 | 9.74 |
| 65+ All Subgroup (1-Dose) | 3.04 | 4.03 | 13.29 | 14.05 |
| 12-64 High-Risk Subgroup (1-Dose) | 1.17 | 1.50 | 3.98 | 4.85 |
| 12-64 High-Risk (1-Dose), 65+ All (2-Doses) | 2.01 | 2.64 | 8.27 | 9.05 |

LY: life year; QALYs: quality-adjusted life years; VSLY: Value of a Statistical Life Year; VQALY: Value per Quality-Adjusted Life Year.

Table 53. Cost-Benefit Ratio (mRNA-1283 relative to mRNA-1273)

| **Strategy** | **Monetized QALYs  (Value=$100k)** | **Monetized QALYs (Value=$150k)** | **Monetized**  **LYs (VSLY $604k)** | **Monetized**  **QALYs (VQALY $717k)** |
| --- | --- | --- | --- | --- |
| **Societal Perspective** | | | | |
| 12-64 High-Risk, 65+ All (1-Dose) | 3.14 | 4.13 | 13.88 | 14.11 |
| 65+ All Subgroup (1-Dose) | 4.12 | 5.49 | 19.49 | 19.21 |
| 12-64 High-Risk Subgroup (1-Dose) | 1.99 | 2.55 | 7.39 | 8.18 |
| 12-64 High-Risk (1-Dose), 65+ All (2-Doses) | 2.88 | 3.79 | 12.73 | 12.93 |

LY: life year; QALYs: quality-adjusted life years; VSLY: Value of a Statistical Life Year; VQALY: Value per Quality-Adjusted Life Year.

Table 54. Cost-Benefit Ratio (mRNA-1283 relative to BNT162b2)

| **Strategy** | **QALYs Monetized (Value=$100k)** | **QALYs**  **Monetized (Value=$150k)** | **LYs**  **Monetized (VSLY $604k)** | **QALYs**  **Monetized (VQALY $717k)** |
| --- | --- | --- | --- | --- |
| **Societal Perspective** | | | | |
| 12-64 High-Risk, 65+ All (1-Dose) | 4.81 | 6.31 | 20.72 | 20.91 |
| 65+ All Subgroup (1-Dose) | 6.56 | 8.67 | 29.60 | 29.29 |
| 12-64 High-Risk Subgroup (1-Dose) | 2.80 | 3.58 | 10.48 | 11.22 |
| 12-64 High-Risk (1-Dose), 65+ All (2-Doses) | 4.42 | 5.79 | 19.01 | 19.18 |

LY: life year; QALYs: quality-adjusted life years; VSLY: Value of a Statistical Life Year; VQALY: Value per Quality-Adjusted Life Year.

### 26 Cheers Checklist

|  | **Item** | **Guidance for Reporting** | **Reported in section** |
| --- | --- | --- | --- |
| **TITLE** | | |  |
| Title | 1 | Identify the study as an economic evaluation and specify the interventions being compared. | Title Page |
| **ABSTRACT** | | |  |
| Abstract | 2 | Provide a structured summary that highlights context, key methods, results and alternative analyses. | Abstract |
| **INTRODUCTION** | | |  |
| Background and objectives | 3 | Give the context for the study, the study question and its practical relevance for decision making in policy or practice. | Introduction |
| **METHODS** | | |  |
| Health economic  analysis plan | 4 | Indicate whether a health economic analysis plan was developed and  where available. | Analysis plan not developed |
| Study population | 5 | Describe characteristics of the study population (such as age range, demographics, socioeconomic, or clinical characteristics). | Introduction; Methods Overview |
| Setting and location | 6 | Provide relevant contextual information that may influence findings. | Overview |
| Comparators | 7 | Describe the interventions or strategies being compared and why chosen. | Overview |
| Perspective | 8 | State the perspective(s) adopted by the study and why chosen. | Overview |
| Time horizon | 9 | State the time horizon for the study and why appropriate. | Overview |
| Discount rate | 10 | Report the discount rate(s) and reason chosen. | Quality of Life |
| Selection of outcomes | 11 | Describe what outcomes were used as the measure(s) of benefit(s) and harm(s). | Overview; Model Structure |
| Measurement of outcomes | 12 | Describe how outcomes used to capture benefit(s) and harm(s) were measured. | Vaccine Coverage; Vaccine Effectiveness; Probabilities for the COVID-19 Consequences Tree |
| Valuation of outcomes | 13 | Describe the population and methods used to measure and value outcomes. | Target Population |
| Measurement and valuation of resources  and costs | 14 | Describe how costs were valued. | Healthcare costs; Lost productivity |
| Currency, price date, and conversion | 15 | Report the dates of the estimated resource quantities and unit costs, plus the currency and year of conversion. | Healthcare costs |
| Rationale and  description of model | 16 | If modelling is used, describe in detail and why used. Report if the model  is publicly available and where it can be accessed. | Overview; Model Structure |
| Analytics and assumptions | 17 | Describe any methods for analysing or statistically transforming data, any extrapolation methods, and approaches for validating any model used. | Methods |
| Characterizing heterogeneity | 18 | Describe any methods used for estimating how the results of the study vary for sub-groups. | Methods Overview |
| Characterizing  distributional effects | 19 | Describe how impacts are distributed across different individuals  or adjustments made to reflect priority populations. | Not applicable |
| Characterizing uncertainty | 20 | Describe methods to characterize any sources of uncertainty in the analysis. | Sensitivity Analyses; Scenario Analyses |
| Approach to engagement with patients and others affected by the study | 21 | Describe any approaches to engage patients or service recipients, the general public, communities, or stakeholders (e.g., clinicians or payers) in the design of the study. | Not applicable |
| **RESULTS** | | |  |
| Study parameters | 22 | Report all analytic inputs (e.g., values, ranges, references) including uncertainty or distributional assumptions. | Included in Methods/Tech appendix & DSA/Scenario Results |
| Summary of main results | 23 | Report the mean values for the main categories of costs and outcomes of interest and summarise them in the most appropriate overall measure. | Results |
| Effect of uncertainty | 24 | Describe how uncertainty about analytic judgments, inputs, or projections  affect findings. Report the effect of choice of discount rate and time horizon, if applicable. | Deterministic Sensitivity Analyses; Scenario Analyses |
| Effect of engagement with patients and others affected by the study | 25 | Report on any difference patient/service recipient, general public, community, or stakeholder involvement made to the approach or findings of the study | Not Applicable |
| **DISCUSSION** | | |  |
| Study findings, limitations, generalizability, and current knowledge | 26 | Report key findings, limitations, ethical or equity considerations not captured, and how these could impact patients, policy, or practice. | Discussion |
| **OTHER RELEVANT INFORMATION** | | | |
| Source of funding | 27 | Describe how the study was funded and any role of the funder in the identification, design, conduct, and reporting of the analysis | Transparency |
| Conflicts of interest | 28 | Report authors conflicts of interest according to journal or  International Committee of Medical Journal Editors requirements. | Transparency |

### 27 References

1.	Centers for Disease Control and Prevention. COVID-NET Monthly rates of laboratory confirmed COVID-19 hospitalizations. Accessed September 9, 2025. <https://www.cdc.gov/covid/php/covid-net/index.html>

2.	Centers for Disease Control and Prevention. COVIDVaxView: COVID-19 Vaccination Coverage and Vaccine Confidence Among Adults. Accessed May 9, 2025. <https://www.cdc.gov/covidvaxview/interactive/adults.html>

3.	Kopel H, Araujo AB, Bogdanov A, et al. Effectiveness of the 2023-2024 Omicron XBB.1.5-containing mRNA COVID-19 Vaccine (mRNA-1273.815) in Preventing COVID-19-related Hospitalizations and Medical Encounters Among Adults in the United States. *Open Forum Infect Dis*. Dec 2024;11(12):ofae695. doi:10.1093/ofid/ofae695

4.	Higdon MM, Baidya A, Walter KK, et al. Duration of effectiveness of vaccination against COVID-19 caused by the omicron variant. *The Lancet Infectious diseases*. Aug 2022;22(8):1114-1116. doi:10.1016/S1473-3099(22)00409-1

5.	Kopel H, Nguyen VH, Boileau C, et al. Comparative effectiveness of Bivalent (Original/Omicron BA.4/BA.5) mRNA COVID-19 Vaccines mRNA-1273.222 and BNT162b2 Bivalent Vaccine in Adults in the US. *Vaccines*. 2023;

6.	Moderna annual report 2023. <https://s29.q4cdn.com/435878511/files/doc_financials/2023/ar/MRNA008_Moderna_2023-Digital-Annual-Report_Bookmarked-1.pdf>

7.	Link-Gelles R. CDC National Center for Immunization and Respiratory Diseases. Effectiveness of COVID-19 (2023-2024 Formulation). Updated June 27, 2024. <https://www.cdc.gov/vaccines/acip/meetings/downloads/slides-2024-06-26-28/03-COVID-Link-Gelles-508.pdf>

8.	Andersson NW, Thiesson EM, Pihlström N, et al. Comparative effectiveness of monovalent XBB.1.5 containing covid-19 mRNA vaccines in Denmark, Finland, and Sweden: target trial emulation based on registry data. *BMJ Med*. 2024;3(1):e001074. doi:10.1136/bmjmed-2024-001074

9.	Habibzadeh F, Habibzadeh P, Yadollahie M. On Measuring Vaccine Effectiveness with Observational Study Designs. *Acta Med Acad*. Aug 2022;51(2):134-146. doi:10.5644/ama2006-124.383

10.	Wilson A, Bogdanov A, Zheng Z, et al. Evaluating the Effectiveness of 2024-2025 Seasonal mRNA-1273 Vaccination Against COVID-19-Associated Hospitalizations and Medically Attended COVID-19 among adults aged ≥18 years in the United States. *medRxiv*. 2025;2025.03.27.25324770

11.	United Nations Department of Economic and Social Affairs (Population Division). World Population Prospects 2024, Online Edition. Accessed September 3, 2024. <https://population.un.org/wpp/downloads?folder=Standard%20Projections&group=Population>

12.	Kompaniyets L, Agathis NT, Nelson JM, et al. Underlying Medical Conditions Associated With Severe COVID-19 Illness Among Children. *JAMA Netw Open*. Jun 1 2021;4(6):e2111182. doi:10.1001/jamanetworkopen.2021.11182

13.	Panagiotakopoulos L. Use of 2025–2026 COVID-19 Vaccines: Work Group Considerations. Presentation to the Advisory Commitee on Immunization Practices, April 15, 2025. Accessed May 5, 2025. <https://www.cdc.gov/acip/downloads/slides-2025-04-15-16/05-Panagiotakopoulos-COVID-508.pdf>

14.	Centers for Disease Control and Prevention. Weekly Cumulative COVID-19 VACCINATION Coverage and Intent, Overall, by Selected Demographics and Jurisdiction, Among Adults 18 Years and Older. Accessed February 28, 2025. <https://data.cdc.gov/Vaccinations/Weekly-Cumulative-COVID-19-Vaccination-Coverage-an/ksfb-ug5d/about_data>

15.	Centers for Disease Control and Prevention. Weekly Parental Intent for Vaccination and Cumulative Percentage of Children 6 Months -17 Years Who are Up to date with the COVID-19 Vaccines by Season, United States. Accessed February 28, 2025. <https://www.cdc.gov/covidvaxview/weekly-dashboard/child-coverage-vaccination.html>

16.	UK Health Security Agency. Almost 80% of eligible over-75s receive spring booster. Accessed May 9, 2025. <https://www.gov.uk/government/news/almost-80-of-eligible-over-75s-receive-spring-booster>

17.	UK Health Security Agency. COVID-19 vaccine surveillance report: Week 23. Accessed May 9, 2025. <https://assets.publishing.service.gov.uk/media/649473a19e7a8b0013932a44/vaccine-surveillance-report-2023-week-23.pdf>

18.	Centers for Disease Control and Prevention. COVID-19 vaccination coverage and vaccine confidence among adults. Accessed July 4 2024. <https://www.cdc.gov/vaccines/imz-managers/coverage/covidvaxview/interactive/adults.html>

19.	Kopel H, Araujo AB, Bogdanov A, et al. Effectiveness of the 2023-2024 Omicron XBB.1.5-containing mRNA COVID-19 vaccine (mRNA-1273.815) in preventing COVID-19-related hospitalizations and medical encounters among adults in the United States: An interim analysis. *medRxiv*. 2024:2024.04.10.24305549. doi:10.1101/2024.04.10.24305549

20.	Moderna data on file 2024. Optum’s de-identified Clinformatics® Data Mart Database. Analysis September 2023 to February 2024 COVID-19 medical attendances and hospitalizations.

21.	Centers for Disease Control and Prevention. Flu burden prevented from vaccination 2022-2023 flu season. Updated December 13, 2023. Accessed June 5, 2024.

22.	Centers for Disease Control and Prevention. Underlying Conditions and the Higher Risk for Severe COVID-19. Accessed May 12, 2025. <https://www.cdc.gov/covid/hcp/clinical-care/underlying-conditions.html>

23.	Havers FP. COVID-19–Associated Hospitalizations — COVID-NET, April 2025 Update. Presentation to the Advisory Committee on Immunization Practices, April 12, 2025. Accessed May 9, 2025. <https://www.cdc.gov/acip/downloads/slides-2025-04-15-16/03-Havers-COVID-508.pdf>

24.	Joshi K, Dronova M, Paterak E, et al. Clinical Impact and Cost-Effectiveness of Updated 2023/24 COVID-19 mRNA Vaccination in High-Risk Populations in the United States. *Infect Dis Ther*. Apr 15 2025;doi:10.1007/s40121-025-01128-z

25.	Moderna. Data on File. Disease burden in patients with Medical conditions. Moderna Bench to Practice. 2024.

26.	VanderWeele TJ. Optimal approximate conversions of odds ratios and hazard ratios to risk ratios. *Biometrics*. Sep 2020;76(3):746-752. doi:10.1111/biom.13197

27.	Moderna Bench to Practice. Respiratory Dashboard. Accessed May 9, 2025. <https://atlas.modernatx.com/bench2practice/Interactive-dashboard>

28.	Mansi JA, Hensler HR, Dawson R, Tuckson R, Wolynn T. Navigating the Evolving Landscape of COVID-19: Strategies to Increase Vaccine Confidence and Improve Vaccination Rates in the United States. *Vaccines (Basel)*. Sep 19 2024;12(9)doi:10.3390/vaccines12091072

29.	Benavidez GA, Zahnd WE, Hung P, Eberth JM. Chronic Disease Prevalence in the US: Sociodemographic and Geographic Variations by Zip Code Tabulation Area. *Prev Chronic Dis*. Feb 29 2024;21:E14. doi:10.5888/pcd21.230267

30.	Palaiodimos L, Chamorro-Pareja N, Karamanis D, et al. Diabetes is associated with increased risk for in-hospital mortality in patients with COVID-19: a systematic review and meta-analysis comprising 18,506 patients. *Hormones (Athens)*. Jun 2021;20(2):305-314. doi:10.1007/s42000-020-00246-2

31.	Ramzi ZS. Hospital readmissions and post-discharge all-cause mortality in COVID-19 recovered patients; A systematic review and meta-analysis. *Am J Emerg Med*. Jan 2022;51:267-279. doi:10.1016/j.ajem.2021.10.059

32.	Taylor CA, Patel K, Pham H, et al. COVID-19-Associated Hospitalizations Among U.S. Adults Aged ≥18 Years - COVID-NET, 12 States, October 2023-April 2024. *MMWR Morbidity and mortality weekly report*. Oct 3 2024;73(39):869-875. doi:10.15585/mmwr.mm7339a2

33.	Yandrapalli S, Aronow WS, Frishman WH. Readmissions in adult patients following hospitalization for influenza: a nationwide cohort study. *Ann Transl Med*. Aug 2018;6(16):318. doi:10.21037/atm.2018.07.18

34.	Xie Y, Choi T, Al-Aly Z. Long-term outcomes following hospital admission for COVID-19 versus seasonal influenza: a cohort study. *The Lancet Infectious diseases*. Mar 2024;24(3):239-255. doi:10.1016/s1473-3099(23)00684-9

35.	Oseran AS, Song Y, Xu J, et al. Long term risk of death and readmission after hospital admission with covid-19 among older adults: retrospective cohort study. *Bmj*. Aug 9 2023;382:e076222. doi:10.1136/bmj-2023-076222

36.	University of Michigan COVID-19 Vaccination Modeling Team. Economic analysis of COVID-19 vaccination. Updated June 24. Accessed February 19, 2025. <https://www.cdc.gov/acip/downloads/slides-2024-06-26-28/05-COVID-Prosser-508.pdf>

37.	Centers for Disease Control and Prevention. National Center for Health Statistics. Long COVID: Household Pulse Survey. Accessed November 30, 2024. <https://www.cdc.gov/nchs/covid19/pulse/long-covid.htm>.

38.	Chalkias S, Dennis P, Petersen D, et al. Efficacy, immunogenicity, and safety of a next-generation mRNA-1283 COVID-19 vaccine compared with the mRNA-1273 vaccine: results from NextCOVE, a phase 3, randomised, observer-blind, active-controlled trial. *The Lancet Infectious diseases*. 2025;

39.	FDA U.S. Food & Drug Administration. Letters for myocarditis.

40.	Klein N. Rapid cycle analysis to monitor the safety of COVID-19 vaccines in near real-time within the vaccine safety datalink: Myocarditis and anaphylaxis. Presentation to the Advisory Committee on Immunization Practices. August 30, 2021. <https://www.cdc.gov/vaccines/acip/meetings/downloads/slides-2021-08-30/04-COVID-Klein-508.pdf>

41.	Boehmer TK, Kompaniyets L, Lavery AM, et al. Association Between COVID-19 and Myocarditis Using Hospital-Based Administrative Data - United States, March 2020-January 2021. *MMWR Morbidity and mortality weekly report*. Sep 3 2021;70(35):1228-1232. doi:10.15585/mmwr.mm7035e5

42.	Center for Disease Control and Prevention. Vaccines for Children Program. Current CDC Vaccine Price List. Updated November 1, 2025. Accessed November 3, 2025. <https://www.cdc.gov/vaccines-for-children/php/awardees/current-cdc-vaccine-price-list.html>

43.	Centers for Medicare & Medicaid Services. 2025 National Physician Fee Schedule Relative Value File October Release. Available from: <https://www.cms.gov/medicare/payment/fee-schedules/physician/pfs-relative-value-files/rvu25d-0>. Last accessed 6 Oct 2025.

44.	Moderna I. NextCOVE Phase 3 Clinical Trial. Clinical Study Report (CSR).

45.	Drugs.com. Acetaminophen/codeine Prices, Coupons, Copay Cards & Patient Assistance. . Accessed August 1, 2024. <https://www.drugs.com/price-guide/acetaminophen-codeine>

46.	Rousculp MD, Hollis K, Ziemiecki R, et al. Burden and Impact of Reactogenicity among Adults Receiving COVID-19 Vaccines in the United States and Canada: Results from a Prospective Observational Study. *Vaccines (Basel)*. Jan 13 2024;12(1)doi:10.3390/vaccines12010083

47.	Walmart. Equate extra strength acetaminophen pain reliever/fever reducer caplets, 500 mg, 100 count. Accessed March 5, 2025. <https://www.walmart.com/ip/Equate-Extra-Strength-Acetaminophen-Pain-Reliever-Caplets-500-mg-100-Count/334176131?classType=VARIANT&athbdg=L1600&from=/search>

48.	Centers for Medicare & Medicaid Services. Quarterly Addenda Updates (October 2025). Available from: <https://www.cms.gov/medicare/payment/prospective-payment-systems/hospital-outpatient-pps/quarterly-addenda-updates>. Last accessed 6 October 2025.

49.	Prosser LA, Harpaz R, Rose AM, et al. A Cost-Effectiveness Analysis of Vaccination for Prevention of Herpes Zoster and Related Complications: Input for National Recommendations. *Ann Intern Med*. Mar 19 2019;170(6):380-388. doi:10.7326/M18-2347

50.	Centers for Disease Control and Prevention. Allergic Reactions Including Anaphylaxis After Receipt of the First Dose of Moderna COVID-19 Vaccine - United States, December 21, 2020 - January 10, 2021. *MMWR Morbidity and mortality weekly report*. January 29, 2021 2021;70(4):125-129.

51.	HCUPnet - Hospital Inpatient National Statistics. 2018 National Diagnoses - Clinical Classification Software Refined (CCSR), Principal Diagnosis: INJ031 Allergic Reactions. . Accessed October 13, 2021. <https://hcupnet.ahrq.gov/#setup>.

52.	Tutle KL WP. Capturing anaphylaxis through medical records. Are ICD and CPT codes sufficient. *Ann Allergy Asthma Immunol*. 2020;124:150-55.

53.	HCUPnet - Hospital Inpatient National Statistics. 2018 National Diagnoses - Clinical Classification Software Refined (CCSR), Principal Diagnosis: CIR005 Myocarditis and Cardiomyopathy. . Accessed October 13, 2021. <https://hcupnet.ahrq.gov/#setup>.

54.	HCUPnet. Average Hospital Costs per ED Visit, United States, 2019 to 2019 Diagnoses--Clinical Classification Software Refined (CCSR), Principal/First-listed - Selected Categories Treat-and-release ED visits (95% confidence intervals). . Accessed November, 2022. <https://datatools.ahrq.gov/hcupnet>

55.	Yehoshua A, Cook AD, Di Fusco M, et al. Health outcomes and economic burden among patients with a COVID-19-associated hospitalization in the United States during the predominance of the XBB and JN.1 omicron lineages. *J Med Econ*. Jan-Dec 2024;27(1):1372-1378. doi:10.1080/13696998.2024.2416873

56.	Kapinos KA, Peters RM, Jr, Murphy RE, Hohmann SF, Podichetty A, Greenberg RS. Inpatient Costs of Treating Patients With COVID-19. *JAMA Network Open*. 2024;7(1):e2350145-e2350145. doi:10.1001/jamanetworkopen.2023.50145

57.	Moderna. Optum Database Analyses (v09062024). Data on File.

58.	Chambers LC, Park A, Cole M, et al. Long-term health care costs following COVID-19: implications for pandemic preparedness. *Am J Manag Care*. Nov 2023;29(11):566-572. doi:10.37765/ajmc.2023.89452

59.	Scott A, Ansari W, Khan F, et al. Substantial health and economic burden of COVID-19 during the year after acute illness among US adults at high risk of severe COVID-19. *BMC Med*. Feb 1 2024;22(1):46. doi:10.1186/s12916-023-03234-6

60.	Walter EB, Schlaudecker EP, Talaat KR, et al. Safety of Simultaneous vs Sequential mRNA COVID-19 and Inactivated Influenza Vaccines: A Randomized Clinical Trial. *JAMA Netw Open*. Nov 4 2024;7(11):e2443166. doi:10.1001/jamanetworkopen.2024.43166

61.	Mercon KR, Rose AM, Cadham CJ, et al. Health Preferences in Transition: Differences from Pandemic to Post-Pandemic in Valuation of COVID-19 and RSV Illness in Children and Adults. *Children (Basel)*. Jan 31 2025;12(2)doi:10.3390/children12020181

62.	Sandmann FG, Tessier E, Lacy J, et al. Long-Term Health-Related Quality of Life in Non-Hospitalized Coronavirus Disease 2019 (COVID-19) Cases With Confirmed Severe Acute Respiratory Syndrome Coronavirus 2 (SARS-CoV-2) Infection in England: Longitudinal Analysis and Cross-Sectional Comparison With Controls. *Clin Infect Dis*. Aug 24 2022;75(1):e962-e973. doi:10.1093/cid/ciac151

63.	PHOSP-COVID Collaborative Group. Clinical characteristics with inflammation profiling of long COVID and association with 1-year recovery following hospitalisation in the UK: a prospective observational study. *Lancet Respir Med*. Aug 2022;10(8):761-775. doi:10.1016/s2213-2600(22)00127-8

64.	Hanmer J, Lawrence WF, Anderson JP, Kaplan RM, Fryback DG. Report of nationally representative values for the noninstitutionalized US adult population for 7 health-related quality-of-life scores. *Med Decis Making*. Jul-Aug 2006;26(4):391-400. doi:10.1177/0272989x06290497

65.	Leuchter RK, Delarmente BA, Vangala S, Tsugawa Y, Sarkisian CA. Health Care Staffing Shortages and Potential National Hospital Bed Shortage. *JAMA Network Open*. 2025;8(2):e2460645-e2460645. doi:10.1001/jamanetworkopen.2024.60645

66.	Brassel S, Neri M, Schirrmacher H, Steuten L. The Value of Vaccines in Maintaining Health System Capacity in England. *Value Health*. Jul 2023;26(7):1067-1072. doi:10.1016/j.jval.2022.06.018

67.	Sandmann FG, Robotham JV, Deeny SR, Edmunds WJ, Jit M. Estimating the opportunity costs of bed-days. *Health Econ*. Mar 2018;27(3):592-605. doi:10.1002/hec.3613

68.	Centers for Disease Control and Prevention. National Center for Immunization and Respirator Diseases. CDC Tracks Hospital Capacity as Respirator Diseases Continue to Spread. Updated January 12, 2024. Accessed March 28, 2025. <https://www.cdc.gov/ncird/whats-new/track-hospital-capacity.html>

69.	Centers for Disease Control and Prevention. Urgent Need to Increase Immunization Coverage for Influenza, COVID-19, and RSV and Use of Authorized/Approved Therapeutics in the Setting of Increased Respiratory Disease Actvity During the 2023-2024 Winter Season. Updated December 14, 2023. Accessed March 28, 2025. <https://www.cdc.gov/han/2023/han00503.html>

70.	Sandmann FG, Shallcross L, Adams N, et al. Estimating the Hospital Burden of Norovirus-Associated Gastroenteritis in England and Its Opportunity Costs for Nonadmitted Patients. *Clinical Infectious Diseases*. 2018;67(5):693-700. doi:10.1093/cid/ciy167

71.	Kohli M, Maschio M, Lee A, et al. The potential clinical impact and cost-effectiveness of the updated COVID-19 mRNA Autumn 2024 vaccines in the United Kingdom. *J Med Econ*. Jan-Dec 2024;27(1):1359-1372. doi:10.1080/13696998.2024.2413288

72.	Biundo E, Dronova M, Chicoye A, et al. Capturing the Value of Vaccination within Health Technology Assessment and Health Economics—Practical Considerations for Expanding Valuation by Including Key Concepts. *Vaccines*. 2024;12(7):773.

73.	Neumann PJ, Cohen JT, Weinstein MC. Updating cost-effectiveness--the curious resilience of the $50,000-per-QALY threshold. *N Engl J Med*. Aug 28 2014;371(9):796-7. doi:10.1056/NEJMp1405158

74.	Centers for Disease Control and Prevention. COVIDVaxView COVID-19 Vaccinations Administered in Pharmacies and Medical Offices, Adults 18 Years and Olcer, United States. Updated March 26, 2025. Accessed March 27, 2025. <https://www.cdc.gov/covidvaxview/weekly-dashboard/vaccinations-administered-pharmacies-medical.html>

75.	French G, Hulse M, Nguyen D, et al. Impact of hospital strain on excess deaths during the COVID-19 pandemic-United States, july 2020-july 2021. *Am J Transplant*. Feb 2022;22(2):654-657. doi:10.1111/ajt.16645

76.	Centers for Disease Control and Prevention. COVID-19 Data from Selected Hospitals. National Center for Health Statistics. Accessed May 24, 2024. <https://www.cdc.gov/nchs/covid19/nhcs.htm>

77.	Agency for Healthcare Research and Quality. Healthcare Cost and Utilization Project (HCUPnet). Accessed February 24, 2025. <https://datatools.ahrq.gov/hcupnet/>

78.	United States Bureau of Labor Statistics. Labor Force Statistics from the Current Population Survey. Accessed October 6, 2025. <https://www.bls.gov/web/empsit/cpseea13.htm>

79.	United States Bureau of Labor Statistics. Employment and Earnings Table B-3: Average hourly and weekly earnings of all employees on private nonfarm payrolls by industry sector, seasonally adjusted. Accessed October 6, 2025. <https://www.bls.gov/news.release/empsit.t19.htm>

80.	Shimabukuro T. Update on myocarditis following mRNA COVID-19 vaccination. Presentation to the Vaccines and Related Biological Products Advisory Committee. Accessed June 7, 2022. <https://www.fda.gov/media/159007/download>

81.	Bonner C, Ghouralal SL. Long COVID and Chronic Conditions in the US Workforce: Prevalence, Productivity Loss, and Disability. *J Occup Environ Med*. Mar 1 2024;66(3):e80-e86. doi:10.1097/JOM.0000000000003026

82.	Chopra V, Flanders SA, O'Malley M, Malani AN, Prescott HC. Sixty-Day Outcomes Among Patients Hospitalized With COVID-19. *Ann Intern Med*. Apr 2021;174(4):576-578. doi:10.7326/M20-5661

83.	Yang J, Rai KK, Seif M, et al. COVID-19-Related Work Absenteeism and Associated Lost Productivity Cost in Germany: A Population-Based Study. *J Occup Environ Med*. Jun 1 2024;66(6):514-522. doi:10.1097/JOM.0000000000003093

84.	Bartsch SM, Chin KL, Strych U, et al. The Current and Future Burden of Long COVID in the United States (U.S.). *J Infect Dis*. Jan 22 2025;doi:10.1093/infdis/jiaf030

85.	Ortega-Sanchez IR, Molinari NA, Fairbrother G, et al. Indirect, out-of-pocket and medical costs from influenza-related illness in young children. *Vaccine*. Jun 13 2012;30(28):4175-81. doi:10.1016/j.vaccine.2012.04.057

86.	Hutton D. Economics of Pfizer maternal RSVpreF vaccine. Presented to ACIP June 21, 2023. Accessed March 6, 2025. <https://www.cdc.gov/acip/downloads/slides-2023-06-21-23/02-RSV-Mat-Ped-Hutton-508.pdf>

87.	Yuasa A, Yonemoto N, LoPresti M, Ikeda S. Productivity loss/gain in cost-effectiveness analyses for vaccines: a systematic review. *Expert Rev Pharmacoecon Outcomes Res*. Apr 2021;21(2):235-245. doi:10.1080/14737167.2021.1881484

88.	Chesson H, Spicknall IH, Kreisel KM, Gift TL. Estimates of the Lifetime Productivity Costs of Chlamydia, Gonorrhea, and Syphilis in the United States. *Sex Transm Dis*. Oct 1 2024;51(10):635-640. doi:10.1097/OLQ.0000000000001973

89.	Islam M, Chesson H, Hutchinson A, Shrestha RK, Song R, Viguerie A, Farnham PG,. Lifetime Productivity Loss Due to HIV Mortality in the United States. Accessed March 2025. <https://www.ispor.org/docs/default-source/intl2024/ispor24islamee271poster136378-pdf.pdf?sfvrsn=f6d83460_04>;

90.	Grosse SD, Krueger KV, Pike J. Estimated annual and lifetime labor productivity in the United States, 2016: implications for economic evaluations. *J Med Econ*. Jun 2019;22(6):501-508. doi:10.1080/13696998.2018.1542520

91.	United States Bureau of Economic Analysis. National Data: Table 1.1.9. Implicit Price Deflator for Gross Domestic Product. Accessed March 19, 2025. <https://apps.bea.gov>

92.	Wittman SR, Martin JM, Mehrotra A, Ray KN. Antibiotic Receipt During Outpatient Visits for COVID-19 in the US, From 2020 to 2022. *JAMA Health Forum*. Feb 3 2023;4(2):e225429. doi:10.1001/jamahealthforum.2022.5429

93.	Tsay SV, Bartoces M, Gouin K, Kabbani S, Hicks LA. Antibiotic Prescriptions Associated With COVID-19 Outpatient Visits Among Medicare Beneficiaries, April 2020 to April 2021. *JAMA*. May 24 2022;327(20):2018-2019. doi:10.1001/jama.2022.5471

1. The COVID-NET data was downloaded on September 12, 2025.
